# Ocular biomarker profiling after complement factor I gene therapy in geographic atrophy secondary to age-related macular degeneration

**DOI:** 10.1101/2024.06.12.24308796

**Authors:** Thomas M Hallam, Emanuela Gardenal, Fraser McBlane, GaEun Cho, Lucy Lee Ferraro, Eva Pekle, Darlene Lu, Kate Carney, Claire Wenden, Hannah Beadsmoore, Sergio Kaiser, Lauren Drage, Thomas Haye, Iris Kassem, Nalini Rangaswamy, Ma’en Obeidat, Cyndy Grosskreutz, Magali Saint-Geniez, David H Steel, Robert E MacLaren, FOCUS Principal Investigators, Scott Ellis, Claire L Harris, Stephen Poor, Amy V Jones

**Affiliations:** Gyroscope Therapeutics Ltd, A Novartis Company, United Kingdom; Translational Research, Ophthalmology, BioMedical Research, Novartis, United Kingdom; Pharmacokinetic Sciences, Translational Medicine, BioMedical Research, Novartis, Switzerland; Clinical Development, Global Drug Development, Novartis, United States; Clinical Development, Global Drug Development, Novartis, Switzerland; Biomarker Development, Translational Medicine, BioMedical Research, Novartis, United Kingdom; Biostatistics, Global Drug Development, Novartis, United States; Pharmacokinetic Sciences, Translational Medicine, BioMedical Research, Novartis, United Kingdom; Biomarker Development, Translational Medicine, BioMedical Research, Novartis, Switzerland; Cell and Gene Therapies, Technical Research and Development, Novartis, United Kingdom; Ophthalmology, Translational Medicine, BioMedical Research, Novartis, United States; Biomarker Development, Translational Medicine, BioMedical Research, Novartis, United States; Translational Research, Ophthalmology, BioMedical Research, Novartis, United States; Bioscience Institute, Newcastle University, Newcastle Upon Tyne, UK; Sunderland Eye Infirmary, Sunderland, United Kingdom; Nuffield Laboratory of Ophthalmology and Oxford Eye Hospital, NIHR Oxford Biomedical Research Centre, Oxford University Hospitals NHS Foundation Trust, Oxford, United Kingdom

**Keywords:** complement system, age-related macular degeneration, gene therapy, biomarker analysis, vitreous humor

## Abstract

**Objective:** Complement biomarker analysis in ocular fluid samples from subjects with geographic atrophy (GA) in a Phase I/II clinical trial of subretinal AAV2 complement factor I (*CFI*; FI) gene therapy, PPY988 (formerly GT005), to understand target pharmacokinetics/pharmacodynamics. Clinical findings were subsequently utilized to investigate the therapeutic dose in an *in vitro* complement activation assay.

**Design, setting and participants:** Biomarker data were evaluated from 28 subjects in FOCUS, a Phase I/II clinical trial evaluating the safety and efficacy of three ascending doses of PPY988.

**Main outcomes and measures:** Vitreous humor (VH), and aqueous humor (AH) from subjects before surgery and at serial timepoints (week 5 or 12, 36, 96) were evaluated for changes in levels of intact complement factors I, B and H (FI, FB, FH) components C3, C4, and C1q and breakdown products (Ba, C3a, C3b/iC3b, C4b) using validated assays and OLINK^®^ proteomics.

A modified *in vitro* assay of complement activation modelling VH complement concentrations was used to compare PPY988 potency to the approved intravitreal C3 inhibitor pegcetacoplan (Apellis) and complement Factor H (FH).

**Results:** An average 2-fold increase in VH FI was observed post-treatment at week 36 and week 96. This correlated with a marked post-treatment reduction in VH concentration of the FB breakdown product Ba and Ba:FB ratio, but minimal changes in C3a and C3b/iC3b levels. Variable concordance in complement biomarker levels in VH versus AH suggest AH is not a reliable proxy for VH for complement activation. During the experimental comparison of doses, a 2-fold increase of FI achieved in the vitreous had only a minor effect on the complement amplification loop *in vitro*, indicating limited impact [IC50: 1229nM]. Pegcetacoplan completely blocks C3a generation at concentrations much lower than the estimated trough level for monthly intravitreal injections [IC50: 2nM]. Supplementation with FH in the assay revealed similar potency to pegcetacoplan [IC50: 6nM].

**Conclusions and relevance:** PPY988 subretinal gene therapy may not have provided sufficient FI protein to meaningfully modulate complement activation to slow GA growth. Reviewing VH biomarkers is important for understanding target expression, pathway engagement, and determining optimal dose, thereby informing future clinical development.

## Introduction

Age-related macular degeneration (AMD) is a leading cause of blindness, particularly in developed countries. In the United States (US) alone, there are approximately 18 million individuals aged 40 and above with early-stage AMD, and 1.5 million with late-stage AMD, which includes geographic atrophy (GA) and/or neovascular AMD ^1^. GA is a condition characterized by the progressive loss of photoreceptors, retinal pigment epithelium (RPE), and underlying choriocapillaris, resulting in a central scotoma ^2^. Reduced vision can significantly impact activities of daily living and quality of life ^3,4^. There have been two recent Food and Drug Administration (FDA)-approved therapies that inhibit complement activity, with a modest slowing of GA growth ^5,6^. However, they require life-long monthly or bimonthly intravitreal (IVT) injections and have so far exhibited limited functional efficacy that present challenges for long term compliance ^7^. Therefore, a one-time gene therapy that provides long-lasting efficacy equivalent to or better than repeated IVT therapies is a promising avenue for exploration.

One such gene therapy, called PPY998 (formerly GT005), is a subretinal gene therapy that supplements complement factor I (*CFI*; FI) protein in the retinal layers where atrophy occurs ^8^. A Phase II study (HORIZON, NCT04566445) was initiated, however a decision to stop the PPY988 program was taken by Novartis based on a recommendation from the independent Data Monitoring Committee following an overall benefit-risk assessment of available data from the program studies. [Press release: Novartis AG; September 11th 2023, : https://www.novartis.com/news/gt005-ppy988-development-program-geographic-atrophy]. This raises important questions about whether *CFI* was sufficiently expressed after subretinal gene therapy, whether there was evidence of modulation of intraocular complement activity, or if this modulation was sufficient to potentially alter the disease course. Such information is crucial to the retina community, as there are many companies developing complement modulating therapies for AMD ^9^. Sharing the learnings from the PPY998 program would improve the risk-benefit analysis for these programs and help understand if new approaches differentiate from PPY998, justifying further clinical study of complement targets for GA.

The complement system, specifically the alternative pathway (AP), plays an important role in the development of AMD, and increasing local FI concentration could be a promising approach to regulate ocular complement activity. The AP is a part of the complement system that constantly surveys for foreign cells and debris and requires tight regulation to prevent inflammation and self-cell damage ^10^. Regulatory proteins like factor H (FH), decay-accelerating factor (DAF; CD55), membrane cofactor protein (MCP; CD46), complement receptor 1 (CR1; CD35), and FI help control the AP, with FI being the only regulatory enzyme ^11^. FI, expressed from the *CFI* gene, cleaves C3b by forming a trimolecular complex with cofactors FH for C3b, and MCP and CR1 for C3b and C4b ^12–18^. Studies have shown that carriers of rare variants (RV) in *CFI* have increased risk of developing advanced AMD and more rapid disease progression ^19–23^. *CFI* RVs can result in low levels of serum FI (Type I variants) or normal levels with functional impairment (Type II variants) ^24,25^, many examples of which were observed in Gyroscope’s SCOPE natural history study ^26^. Research has also suggested that doubling serum FI levels may help control complement overactivation driven by other AMD risk genetic factors mapping to complement genes ^27^. Based on these findings, clinical trials were conducted to explore the potential of sub-retinal delivery of AAV2-*CFI* as a gene therapy for treating dry AMD ^8^.

The FOCUS study is a Phase I/II open-label study evaluating the safety, and dose response of three doses of PPY988 AAV2-*CFI* subretinal gene therapy in GA. To assess transgene pharmacokinetics and pharmacodynamics, vitreous humor (VH) and aqueous humor (AH) samples were collected at baseline (BL) and four time points up to 96 weeks after dosing and were evaluated for exploratory complement biomarkers. This report shares findings regarding change from baseline in FI and complement protein levels and activation, over time after vitrectomy and PPY988 administration. Levels of FI achieved in VH from FOCUS subjects were then used in an *in vitro* simulated human vitreous complement activation assay, to gain insights on the therapeutic potencies of FI, C3 inhibitor pegcetacoplan (Syfovre, Apellis), and FH. We also offer critical learnings for other AMD complement gene therapy programs and a guide for future analyses of complement biomarker data in ocular samples.

## Methods

### Ethics

The FOCUS study was conducted in accordance with the tenets of the Declaration of Helsinki and all other local regulations, in accordance with the International Conference on Harmonization E6 Guidelines for Good Clinical Practice and with applicable local, state, and federal laws. Institutional review board or ethics committee approval was obtained, and the study was registered at ClinicalTrials.gov (reference: NCT03846193) and EudraCT (reference: 2017-003712-39). All subjects provided written informed consent to participate, and an independent data monitoring committee provided ongoing oversight.

### Study design

FOCUS is an open-label first in human Phase I/II multicentre clinical trial to evaluate the safety, and dose response of PPY988 (AAV2-*CFI)* ^8^, administered as a single subretinal injection in subjects with GA secondary to AMD, followed for up to 240 weeks (5 years) post-dosing. Subjects were administered one of three doses to the study eye under local anaesthesia by a qualified vitreoretinal surgeon: 2E10 viral genome (vg; low), 5eE10 vg (medium) and 2E11 vg (high). At screening, a saliva sample was taken from subjects and extracted DNA was analysed for *CFI* RV using targeted next generation sequencing ^21^. Genotyping results were not required at screening if already available through participation in a previous Gyroscope sponsored study. The term ‘rare’ was defined as variants that changed the *CFI* coding sequence, had ≤1% minor allele frequency in GnomAD non-Finnish Europeans (V2.1.1, accessed http://gnomad.broadinstitute.org), and were previously associated with FI haploinsufficiency ^11^. A detailed description of study design, route of administration, dose, number of subjects, biomarker matrices, sampling timing and methodology are provided in **Supplementary Methods and Supplementary Table 1.**

### Complement protein and total protein quantification

AH and VH samples of subjects were taken on the treated eye immediately prior to subretinal surgery at baseline (BL) and at 5 or 12 weeks, 36, and 96 weeks post-treatment, as described in detail in **Supplementary Table 1**. Samples were rapidly frozen at −80 °C and transported frozen for analysis. Prior to analysis, samples were thawed and subjected to centrifugation at 17,000g for 10 minutes at 4°C.Samples were kept on ice during sample preparation.

Complement factors C2, C4b, C5, C5a, factor D, mannose binding lectin (MBL), FI, C1q, C3b/iC3b, C3, FH, FB and C4 were quantified in AH and VH using the MILLIPLEX Human Complement Panel 1 and 2 (cat. no. HCMP1MAG-19K and HCMP2MAG-19K, EMD Millipore Corporation, Billerica, MA) following extensive quality control and validation of calibration curves. Plates were read using the Luminex MagPix and xPONENT software (Luminex Corporation, Austin, TX). C5a was omitted from further analysis as measurements fell under the lower limit for quantification.

Complement factors C3a and Ba were quantified in AH and VH using MicroVue EIA kits (cat. no. A032 and A034, QuidelOrtho Corporation, San Diego, CA) following manufacturer instructions. Plates were read using a Tecan Sunrise absorbance microplate reader using the Magellan Tracker data analysis software (Tecan Trading AG, Switzerland).

Each assay was validated and considered fit for purpose. Sample processing and analyses were conducted in accordance with Good Clinical Laboratory Practice, the analytes or measurements that did not meet acceptance criteria (coefficient of variation of 20%, or 30% when results are below the lower limit of quantification but above limit of detection) were excluded from the final analysis. The number of successfully sampled subjects for each biomarker at each time-point is reported in **Supplementary Table 2**.

Total protein was quantified in AH and VH using the Quant-iT protein assay kit (cat. no. Q33210, Invitrogen, Thermo Fisher Scientific, Waltham, MA) as per the manufacturer’s instructions. Plates were read using a Varioskan LUX fluorescent microplate reader using the SkanIt data acquisition software (Thermo Fisher Scientific, Waltham, MA).

### *In vitro* modelling the ocular complement system and potency assay

The assay was performed using reagents from the Wieslab AP assay kit (cat. no. COMPLAP330, Svar Life Science AB, Sweden) and purified human serum complement proteins (Complement Technology Inc., Texas USA). All preparation steps were performed on ice. Serial dilutions of the test analytes FI, FH and pegcetacoplan were separately prepared and added to a mix of the complement components C3 (2543.3ng/mL), FB (853.6ng/mL), FH (296.2ngmL), FI (603.8ng/mL) and Properdin (18.1ng/mL) simulating the final median concentrations measured in the VH baseline of FOCUS subjects (concentrations of C3, FB, FH, FI are from *CFI* RV negative population, Properdin concentration is from a limited number of FOCUS subjects from both populations). The simulated vitreous complement mix was transferred on the lipopolysaccharide (LPS) coated Wieslab plate on ice. FD was separately added to the mix, the plate was immediately put at 37°C and incubated for 1 hour. At the end of the incubation, the solution was transferred into a dilution plate and kept on ice, EDTA was added to each well to obtain a final 20mM concentration. Ba and C3a generated in the assay were measured by using the MicroVUE EIA kits (cat. no. A032 and A034, QuidelOrtho Corporation, San Diego, CA) following the manufacturer instructions. The ELISA plates were scanned using the Varioskan plate reader and analysed using the SkanIT software (Thermo Fisher Scientific, Waltham, MA). The *in vitro* FI potency comparability assay is described in **Supplementary Methods**. The trough level of pegcetacoplan in the VH after 27 days was calculated by halving the treatment dose of 15mg/eye 6 times (pegcetacoplan half-life 4.5 days), the number obtained was converted to µg/mL using an estimated volume of VH of 4 mL ^28^.

### Statistics

All subject sample biomarker analyses were performed in R version 4.1.0. The analysis set consisted of all subjects who received subretinal delivery of PPY998 from cohorts 1 to 4 and had at least one non-missing biomarker assessment performed at BL and either week 36 or week 96. Given sparsity of data at week 5 and week 12, the data were combined, and the average value was used in instances where subjects had non-missing data from both visits. Concentrations values above the upper limit of quantification (ULOQ) was set to ULOQ and values below the lower limit of detection (LOD) were set to LOD. Please see Supplementary Tables 3-4 for limits.

Significance testing for the change in protein level from BL to week 36 or week 96 were performed on the differences of log-transformed data with the non-parametric Wilcoxon rank-sum test. Means and standard error bars are shown in the respective figures. Correlations between VH and AH sampling, as well as between FI and other proteins, were summarized with the Spearman’s Rank correlation coefficient. As analyses were exploratory in nature and hypothesis-generating, no adjustments for multiple testing were performed.

For the *in vitro* VH modelling assay, to calculate IC50s, 4-parameter logistic curves were generated using GraphPad Prism version 10.0.0 for Windows, GraphPad Software, Boston, Massachusetts USA, www.graphpad.com.

## Results

### Biomarker detection in FOCUS subjects that received PPY988

In FOCUS Part 1 and 2 (cohorts 1 to 4), 31 subjects received one of three doses of PPY988 via the standard subretinal delivery approach (**Supplementary Table 1**). A total of 97% of the ocular samples expected under the protocol were obtained prior to early termination of the study which triggered cessation of further ocular fluid sampling. A total of 88 VH, 63 AH and 45 plasma samples from 30 subjects were obtained at various timepoints including BL, and 5 or 12, 36 and 96 weeks post-treatment (**Supplementary Methods and Supplementary Tables 1 and 2**).

In FOCUS, PPY988 was well tolerated with no serious ocular AEs, some dose-related RPE changes in the bleb area were evident and there was no clinically significant vector-related inflammation. A full account of the FOCUS study safety and clinical data, as well as full Phase II PPY988 safety, efficacy and biomarker data will be reported separately following study completion.

### Vitrectomy for subretinal AAV-*CFI* gene therapy procedure results in sustained loss of protein from the VH

To test impact of total vitrectomy performed during transvitreal subretinal delivery of PPY988 (**Figure 1A and B**) influencing global proteomic matrix changes and biomarker analyses/interpretation, total protein (TP) measurements were obtained at BL and each sampling timepoint (according to sample availability). In VH, a ∼30% reduction in TP from baseline was observed at either week 5 or 12 and week 36 (NS, P=0.162), with a significant >50% reduction identified at week 96 (P=0.033), while there were minimal changes in AH TP at week 5/12, 36 or 96 (**Figure 1C**). Because VH TP was reduced post-operatively, we performed TP normalisation to compare the results with non-normalised data interpretation for VH but not AH.

**Figure 1.**
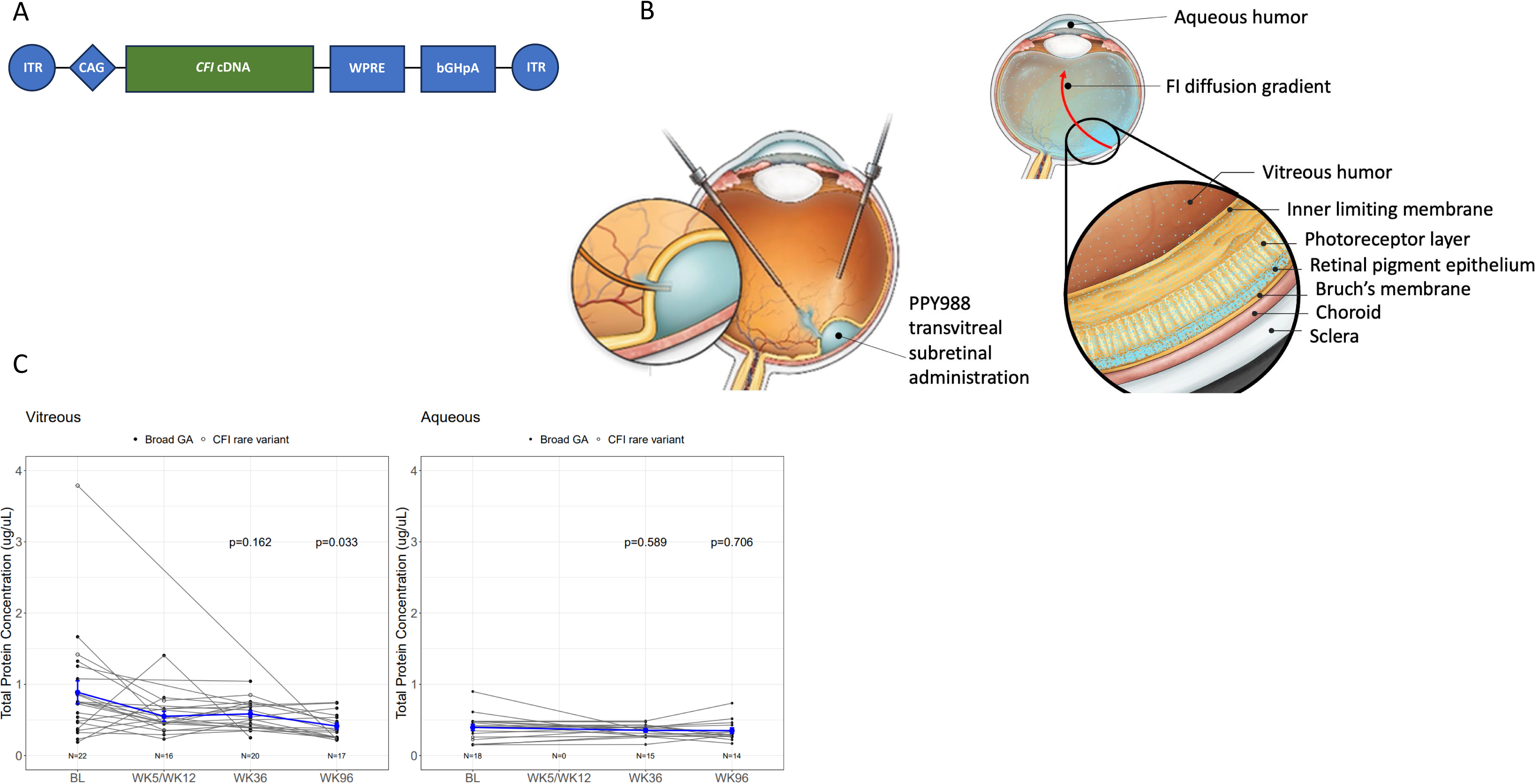
Surgical delivery of PPY988 AAV2-*CFI*. (A) PPY988 vector design. FI protein is secreted by cells targeted by AAV2 serotype and containing the cDNA sequence for wild-type *CFI* (NM_000204.4) driven by a CAG promoter ^8^. (B) Subretinal delivery of PPY988. (C) Vitrectomy during SR delivery of PPY988 results in loss of total protein (TP) in the vitreous humor. TP concentration (y-axis, log2 µg/µL) was measured at baseline and 5 or 12, 36, and 96 weeks post-surgery (y-axis). Mean +/− SE is shown by blue bars and lines whilst all subjects are shown by black lines for both vitreous (left) and aqueous (right) humor. A significant reduction in TP was observed at week 96 in vitreous humor (*P=0.033). Significance is defined as *P<0.05. BL = Baseline; WK= week.

### AAV2-*CFI* delivery results in a sustained increase in FI protein level in the VH and AH

Transgene expression was measured in VH and AH samples at weeks 5 or 12, 36, and 96 after treatment with PPY988. On average, we identified a 2-fold increase in VH FI at week 36 compared to baseline, with non-TP normalized baseline concentrations of 500 ng/mL compared to 1000 ng/mL at week 36 (non-normalised P<0.001; TP normalised increase ∼2-fold (P=0.002)) (**Figure 2A, 2B**). The increase in FI concentration was sustained, with a ∼2.5-fold increase in FI observed at week 96. (mean ∼1250 ng/mL) (non-normalised P<0.001; TP normalised increase ∼3-fold (P=0.002)). *CFI* RV (n=5) and non-*CFI* RV (‘broad’) (n=19) genotype groups demonstrate comparable magnitudes of absolute FI elevation post-treatment, with the overall *CFI* RV cohort FI starting concentration ∼50% reduced (NS). Elevated FI expression in AH samples at week 36 (P<0.001) were sustained out to week 96 (P<0.001) (**Figure 2C, 2D**). Lastly, there was a variable response to the three PPY988 doses in both VH and AH with no significant dose effect observed (Figure 2C, 2D).

**Figure 2.**
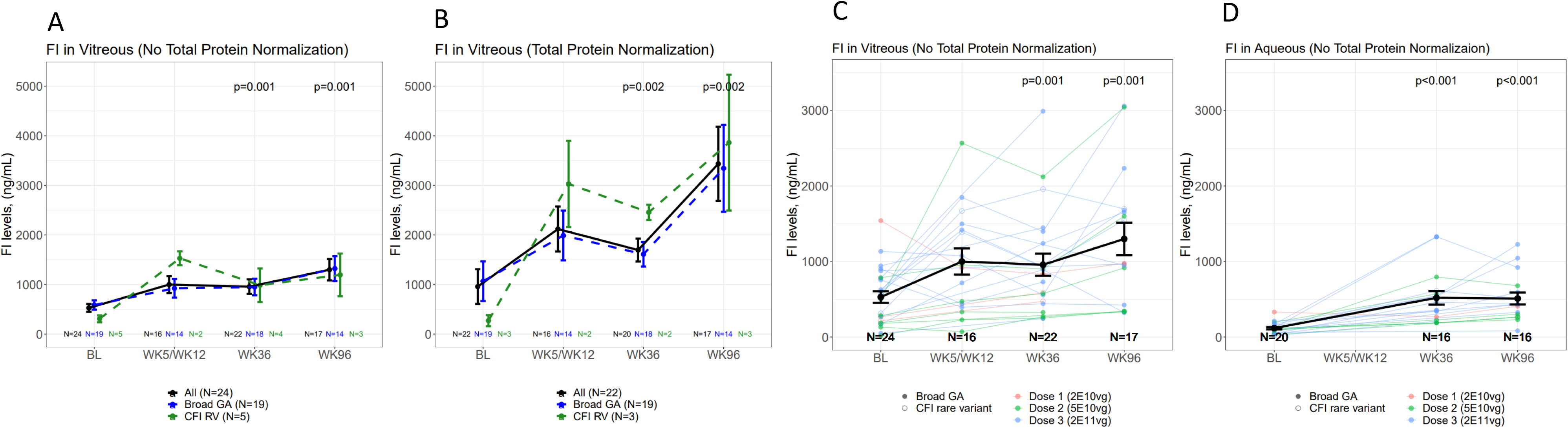
Temporal expression of factor I (FI) in vitreous humor (VH) and aqueous humor (AH) after PPY988 delivery. (A,B) FI levels (y-axis, ng/mL) in VH were significantly increased at week 36 and 96 post-surgery (x-axis) both when non-normalised (A) (P=0.001 for both timepoints) and total protein -normalised (B) (P=0.002 at both timepoints). Mean +/− SE is shown by bars for all (black lines) broad (blue lines) and CFI RV (green lines) groups. (C,D) FI level (y-axis, ng/mL) for each subject in the cohort is tracked by week (x-axis) and separated by dose (2E10 vg: orange lines; 5E10 vg: green lines; 2E11 vg: blue lines) in VH (C) and AH (with significant increase in AH FI at week 36 and 96 (both P<0.001)) (D), where mean +/− SE shown by black bars. Significance is defined as *P<0.05; **P<0.01, ***P<0.001. N numbers are shown above X-axis. BL = Baseline; WK= week.

### Increased FI expression results in a reduction in Ba and Ba:FB in the VH

To test for FI target engagement at the level of formation of the alternative pathway C3 convertase, we measured Ba and FB concentrations at each sampling week after subretinal delivery of PPY988. A reduction in Ba in the VH is indicative of a reduction in C3 convertase formation in the ocular environment. We observed that Ba levels were reduced by ∼2-fold compared to baseline at week 36 and 2.5-fold at week 96 in non-normalised data (week 36: P=0.004; week 96: P=0.003) (**Figure 3A**) and TP normalised data (week 36: P=0.002; week 96: P=0.038) (**Figure 3B**). To add confidence to this read-out by accounting for fluctuations in endogenous FB biosynthesis, ratios of intact complement proteins and their breakdown products were assessed as in previous studies ^29–31^. Ba:FB ratio changes post-treatment reflected the raw data. When reviewing biomarker levels from both non-normalized and TP normalized datasets, a significant reduction in Ba:FB was observed at week 36 (both P<0.001) and at week 96 (non-TP normalized (P=0.001); TP normalized (P=0.033)) (**Figure 3C, 3D**).

**Figure 3.**
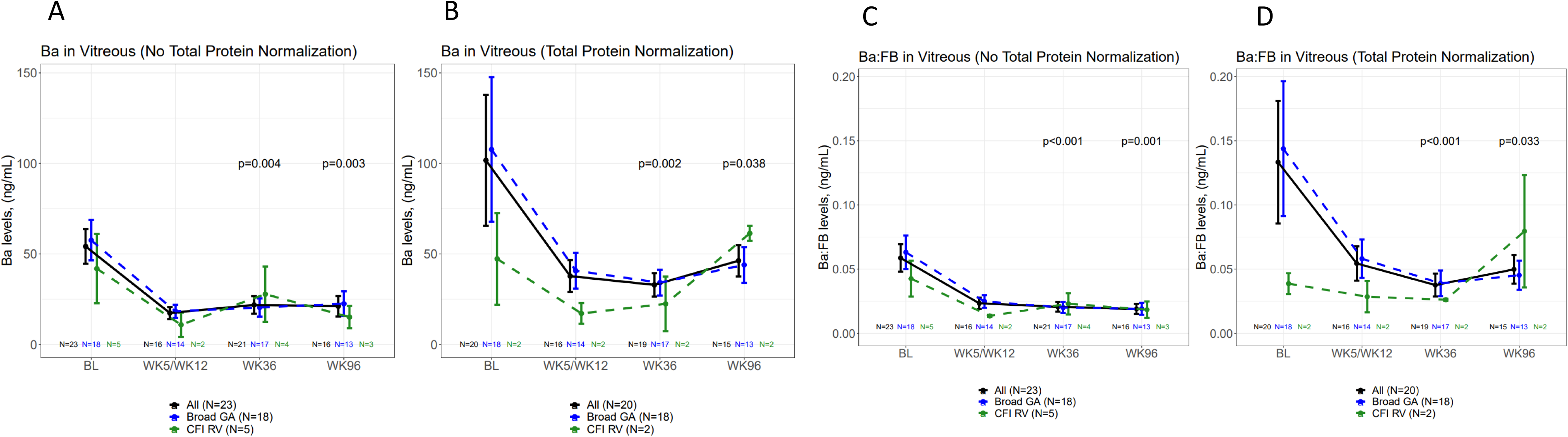
Modulation of Ba and Ba:FB by PPY988 delivery. (A,B) Ba concentrations (Y-axis, ng/mL) in vitreous humor (VH) samples by week post-surgery (X-axis). Mean +/− SE is shown by bars for all (black lines) broad (blue lines) and CFI RV (green lines) groups. Significant reductions in Ba were observed at week 36 for both non-normalised (P=0.004) (A) and total protein (TP) normalised (P=002) (B) data and week 96 for non-normalised data (P=0.003) and TP normalised (P=038). (C,D) Ratios of Ba:FB concentration (y-axis, ng/mL) were calculated for each subject at each sampling post-surgery week (x-axis). Mean +/− SE is shown by bars for all (black lines) broad (blue lines) and *CFI* RV (green lines) groups. Significant reductions were identified at week 36 for both non-normalised (P<0.001) (C) and TP normalised (P<0.001) (D) Ba:FB ratios, with a reduction at week 96 for non-normalized (P=0.001) and TP normalized (P=0.033) ratios. Significance is defined as *P<0.05; **P<0.01, ***P<0.001. N numbers are shown above X-axis. BL = Baseline; WK= week.

### Increased FI expression results in modest alterations in VH and AH complement biomarker profiles

To investigate the effect of PPY988 delivery on complement AP and classical pathway (CP) activity, a panel of biomarkers including FI, C3, FB, FH, C1q, Ba, C3a, C3b/iC3b, C4 and C4b were measured in the VH and AH (in addition to plasma) at BL, week 5 or 12, week 36 and week 96. The complete set of concentrations at BL are reported in the **Supplementary Table 3** (VH), **Supplementary Table 4** (AH), and **Supplementary Table 5** (plasma). In addition, Ba:FB, C3a:C3 and C3b/iC3b:C3 ratios were calculated at each time-point. Baseline values for each biomarker with descriptive statistics are reported in **Supplementary Table 3** (VH) and **Supplementary Table 4** (AH), with baseline and post-treatment pairwise correlations between FI and biomarkers presented in **Supplementary Figure 1** (VH) and **Supplementary Figure 2** (AH). Fold changes in complement biomarker concentration from baseline are presented for each biomarker or ratio at each post-surgery sampling week in VH non-normalised data (**Figure 4A**), VH TP normalised data (**Figure 4B**) and AH non-normalised data (**Figure 4C**). In the non-normalised VH dataset, we observed reductions in Ba and Ba:FB, with a reduction in C1q possibly indicative of reduced ocular immune cell (e.g. microglial) activation ^32^ **(Figure 4A**). We detected no modulation of VH C3a and C4b. Although levels of C3b/iC3b per unit of C3 fall substantially at week 96 in both non-normalized (P=0.002) and TP-normalized (P=0.012) analyses (**Supplementary Figure 3**), pairwise correlation analyses reveal that no biomarkers or ratios were negatively correlated with increasing FI level at week 36 or 96 post-treatment (**Supplementary Figures 1 and 2**). Minimal reduction of CP activity was indicated by C4b:C4 ratio analysis, however, this was not observed when normalizing for TP (**Supplementary Figure 4**). VH FB and FH concentrations do not fluctuate substantially from BL values after treatment with PPY988. Overall, TP normalisation did not substantially alter the interpretation of VH biomarkers with broadly similar trends observed **(Figure 4B**).

**Figure 4.**
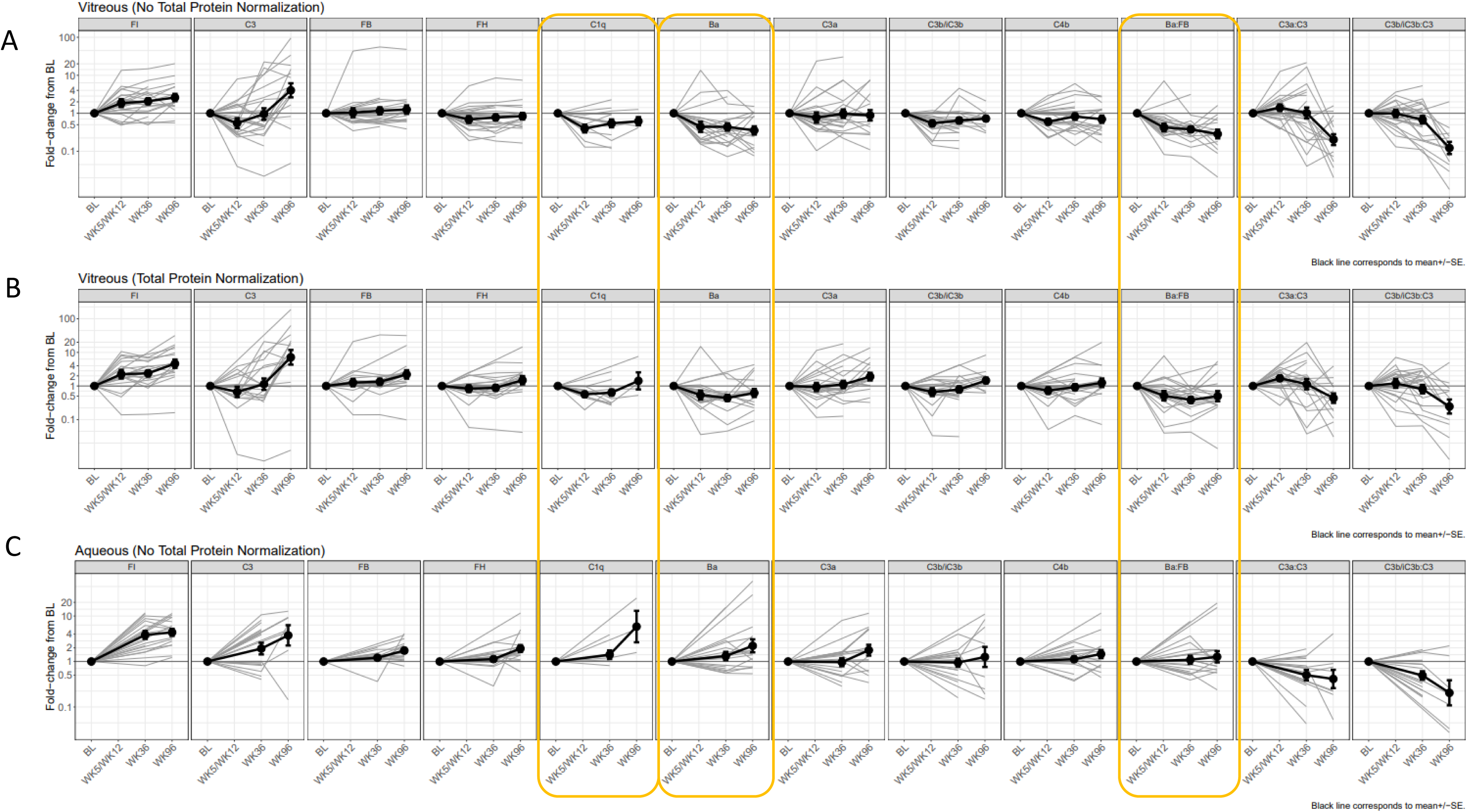
Modulation of complement AP and CP biomarkers by PPY988 delivery. Fold-changes (y-axis) in complement biomarker [FI, C3, FH, FB, C1q, Ba, C3a, C3b/iC3b, C4b, Ba:FB, C3a:C3, C3b/iC3b:C3] concentrations from baseline at each post-surgery week (VH: week 5/12, 36 and 96; AH: week 36 and 96; X-axis) are shown in VH with no normalisation (A), VH with total protein normalisation (B), and AH with no normalisation. Mean is shown by thick black lines, +/− SE is shown by bars. Individual subject changes are shown by grey lines. Yellow boxes highlight disparate trends in VH vs AH biomarkers. BL = Baseline; WK= week.

### AH sampling is not reflective of VH complement activation profile

Complement biomarker profiling in AH demonstrated increasing concentrations of a majority of biomarkers (**Figure 4C**), including for Ba, C1q, and C3a (at week 96) which was discordant with VH sampling data. Whether these components (C1q) and breakdown products (Ba, C3a) are generated locally, systemically, or diffusely from the retina or VH is unclear. A similar trend to VH was observed when considering the increase in AH C3 and associated fall in C3a:C3 and C3b/iC3b:C3 ratios at week 96 in particular. However, in the case of AH, the increase in C3 was also observed at week 36, suggesting compartmentalization of response of the AH vs the VH. In line with these data, pairwise correlations between VH and AH levels of Ba, C3a and C3b/iC3b post-treatment are modest and variable (**Supplementary Figure 5**), with only FI consistently well correlated between the two ocular compartments.

### Proteomic analysis on VH using Olink provides supportive evidence for target expression and modest modulation of the complement system

Olink proteomic analysis of the VH samples identified a 2-fold median increase in week 36 FI levels relative to baseline (**Supplementary Methods**, **Supplementary Figure 6a**). Further review of differentially expressed proteins revealed trends for elevation of additional complement components including factor B, properdin and C2 (**Supplementary Figure 6b, Supplementary Table 6).**

### VH modelling of complement activation reveals inferior potency of FI vs pegcetacoplan

By modelling the components of the VH based on median measurements of C3, FB, FH, FI (broad cohort measurements) and properdin (all measurements) at baseline in the FOCUS study, we compared the potency of FI, FH and the C3 inhibitor pegcetacoplan for their effectiveness in controlling the generation of C3a on an LPS-coated plate in a simulated ocular environment context (**Figure 5**). In this assay, C3 is directly activated by O-antigens on LPS thus triggering activation to drive the AP amplification loop. A wide range of FI, FH and pegcetacoplan concentrations were titrated through an artificial VH matrix so that comparisons could be made to clinically relevant levels of both FI and pegcetacoplan. Serum purified FI was revealed to be of similar potency to FI expressed from Good Manufacturing Practice (GMP) manufactured PPY988 (**Supplementary Figure 7**). The trough level of pegcetacoplan at 27 days was estimated using injection dose and half-life information available on the Syfovre^TM^ prescribing information datasheet. No diffusion data in human was available so this was not counted in the trough calculation. The concentration obtained was similar to that reported in the pegcetacoplan rabbit pharmacokinetics (PK) study and PK/PD prediction study published by Apellis ^33,34^. Pegcetacoplan vitreal trough level was compared to the 1^st^ and 3^rd^ quartile increase (at week 36 vs BL) in FI associated with PPY988 delivery. Pegcetacoplan exhibited greater potency than FI, with complete inhibition of the AP amplification loop and C3a generation at predicted trough level (∼59 μg/mL), whilst FI was observed to have minimal effect at levels of expression associated with PPY988 after 36 weeks. Overall curves generated by logistic regression revealed an IC50 of 1229 nM for FI, whilst FH [IC50: 6 nM] was similar in potency to pegcetacoplan [IC50: 2 nM]. The estimated vitreous trough level of pegcetacoplan was approximately 100-fold greater than that required for complete inhibition of the AP in this assay. On the other hand, FI was observed to have minimal effect on AP activation at the levels of PPY988 measured in VH after 36 weeks of treatment.

**Figure 5.**
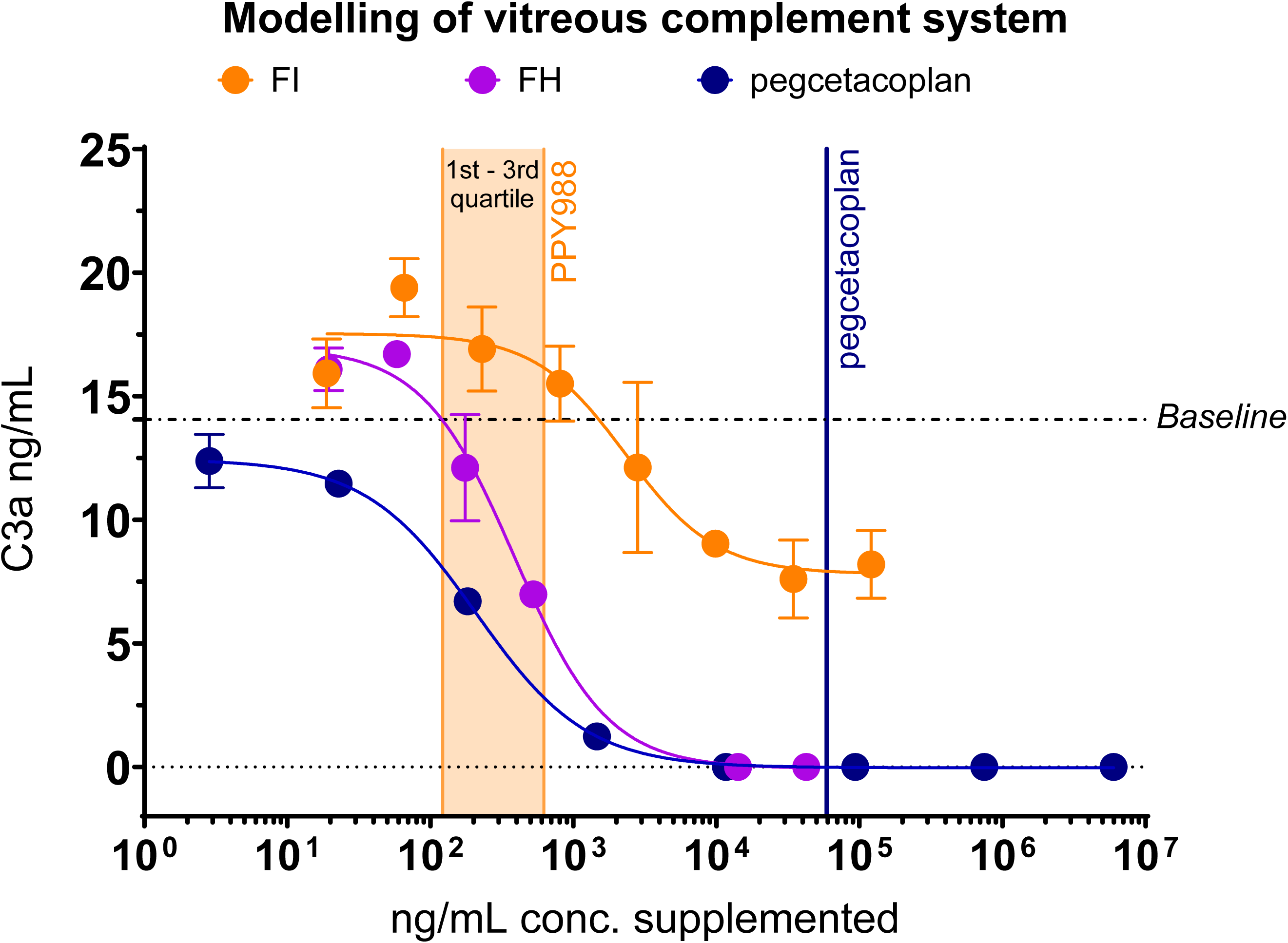
*In vitro* modelling of vitreous humor complement activation inhibition by FI vs pegcetacoplan and additional complement regulators. FI (orange), FH (purple), and pegcetacoplan (blue) were titrated (x-axis, ng/mL) through artificial vitreous complement matrix on LPS-coated Weislab^TM^ plates and C3a generation was measured by ELISA (y-axis, ng/mL). C3a is a marker of convertase activity in the complement activation cascade, the potency of complement inhibitors or regulators is reflected in lower amount of C3a generated in the assay when testing comparable concentrations. Mean is shown with +/−SD represented by bars and 4-parameter fit logistic regression curves are shown by lines. The 1^st^-3^rd^ quartile of increased FI expression from baseline after 36 weeks of PPY988 delivery is shown by orange area. The calculated trough concentration of pegcetacoplan in eyes 27 days after IVT injection is shown by the blue vertical line.

## Discussion

Gene therapy has been suggested as a promising treatment approach for slowing macular degeneration in GA secondary to AMD. This manuscript describes a comprehensive examination of the pharmacokinetic / pharmacodynamic properties of complement biomarkers in the context of the PPY988 GA subretinal gene therapy program and provides technical considerations for analyzing biomarker data from treated subjects. Despite the early termination of the clinical development program, the PPY988 study achieved notable milestones in the field, including the implementation of subretinal delivery of gene therapy for GA and the use of serial vitreous sampling. Herein, we present for the first time an analysis of VH complement biomarkers at baseline and after surgical delivery of a subretinal gene therapy. Baseline levels were similar to a previous study of VH complement ^35^ with potentially confounding loss of total protein also observed in our study, highlighting the need to analyze data in the context of changing total protein levels post-vitrectomy. Our observations reveal consistent elevation of VH FI levels over a 96-week period, accompanied by reduced detection of the FB activation product Ba and ratios of Ba:FB, consistent with sustained modulation of complement AP activity in the vitreous humor compartment. These data confirm that AAV gene therapy has a consistent and lasting effect. Furthermore, although unpowered due to low N, the effects of FI gene therapy on complement breakdown product biomarker profiles (Ba (**Figure 2A**) and C3b/iC3b (**Supplementary Figure 3A**)) appear to be comparable in both *CFI* RV positive and negative sub-populations, which is counter to the original hypothesis of a more dramatic effect in a sub-population with lower baseline FI levels ^19,20^. However, due to small sample size this finding should be interpreted with caution. Although the impact on C3 breakdown products was minimal at week 36, it became significant at week 96. Elevation of FI and modulation of the broader complement system was also observed in Olink proteomic profiling of VH samples (**Supplementary Figure 6**), supporting the complement biomarker findings in the validated panel. Elevated levels of FI were detected in both AH and VH. PPY988 efflux from the sub-retinal injection may have transduced cells in the anterior chamber of the eye resulting in an increase in AH FI concentration. However, the correlation of other complement biomarkers between VH and AH was inconsistent, in particular, Ba and C1q exhibited disparate post-treatment trends (**Figure 4**), indicating that AH alone may not be a reliable biomarker matrix for assessing the activity of complement biomarkers at the target tissues. This cautionary advice should be considered in context of previous reports, suggesting AH could be a broadly reliable proxy for VH ^36^, albeit that only full length complement proteins were studied therein. To ensure quality control, protein level normalization was added to the analysis comparing biomarker levels before and after vitrectomy. Interestingly, the overall conclusions remained consistent regardless of protein adjustment, which strengthens confidence in the study’s findings. After determining the futility of the Phase II study (HORIZON, NCT04566445), an *in vitro* assay using clinically relevant levels of complement proteins was conducted to investigate a possible biomarker-related explanation. The results showed that pegcetacoplan had higher potency than FI in reducing C3 convertase activity *in vitro* suggesting that limited *in-vitro* potency of FI may have contributed to the lack of clinical success at Phase II. Although the translatability of the VH modelling assay to GA remains to be demonstrated, FI’s lower potency suggests that further increasing FI concentration on its own by gene therapy may not achieve clinically meaningful levels of C3 inhibition. It should however be noted that intravitreal pegcetacoplan will be at its highest concentration in the mid-vitreous whereas FI will most likely be at the highest level in the subretinal space where it is being produced. Hence directly comparing vitreous levels of each compound may underestimate the activity of FI compared with pegcetacoplan at the level of the subretinal space. Notably, that FI does not reduce C3a breakdown to baseline even at supermolecular concentrations suggests cofactor limitation that might be more pronounced in VH given FH is not in excess of FI as is the case in blood ^27^. Moreover, there are additional cell-bound cofactors for FI that may be present in retinal tissue that are not present in the VH and could further alter the kinetics of the assay, such as MCP (CD46) albeit with disease related changes observed ^37^. However, the findings of elevated FI in the VH after sub-retinally delivered *CFI* gene therapy and changes in complement pathway proteins post-treatment build confidence that VH matrix is at least partially reflecting the complement system at the retinal layers and treatment site, and is a valid biomarker for PK/PD insights in response to treatment.

The lack of efficacy in the PPY988 Phase II study does not detract from the viability of complement activation as an effective target for intervention in GA, given the therapeutic effect observed for complement inhibitors pegcetacoplan and avacincaptad pegol (Izervay; Iveric Bio) ^5,6^, and this study’s observation that pegcetacoplan achieves a much greater potency than FI *in vitro*. FI had a disproportionally larger effect upon Ba compared to C3a and C3b/iC3b, which may be due to the lability of C3a, differences in diffusion capabilities of the molecules from the retina, and possible contribution of C3 concentration from the choriocapillaris as well as biosynthesis by local immune cells. In addition, we hypothesize FI’s capacity to regulate the production of Ba without causing significant changes in the levels of C3b and C3a in the fluid phase could be contributed to by regulation of AP tickover, resulting from binding and activation of FB on water-hydrolysed C3 (C3H2O) followed by release of Ba^10^. Similarly, the intravitreal administration of a neutralizing factor D Fab, which was found to be ineffective in Phase III trials for GA, was also able to reduce ocular Ba levels ^29^. However, it did not have a significant impact on the generation of VH C3 breakdown products. In this study, by week 96, there was an observed rise in the levels of full-length C3 and a corresponding decrease in the ratios of breakdown products to full length. While the specific implications of this change are uncertain, it potentially suggests a reduction in C3 consumption, which aligns with the known activity of FI, but this may be confounded with upregulated biosynthesis of C3 given its acute phase nature ^38^. C3 breakdown products only partially diffuse due to cell-surface adherence. Based on this understanding, we hypothesize that changes in C3 breakdown products may only become detectable when a certain threshold of modulation in tissue complement activity is reached. Overall, the data suggests that fully functional FI is being secreted into the VH, but the regulatory effects on the level of Ba may be representative of convertase formation in the VH and not the macula retina/RPE nor the choroid. To validate this hypothesis, one approach would be to conduct vitreal sampling using an effective drug targeting C3 for GA in a larger cohort. Further investigations are needed to gain a better understanding of these possibilities.

The study has several limitations that should be considered when interpreting the results. Firstly, the study did not include a control group, which means that the observed changes in complement biomarkers may not be solely attributed to transgene expression. Other factors, such as the impact of subretinal gene therapy administration triggering local inflammation or the effects of total vitrectomy on the protein environment, could also contribute to these changes. During the study, no significant safety concerns or adverse events were reported aside from dose-responsive incidents of RPE changes in the area of the subretinal injection. However, increased levels of antibodies to the vector and transgene were observed following treatment ^39^. It is important to note that these antibody levels were transient and resolved shortly after treatment, indicating that the observed antibody responses were not persistent or associated with any significant clinical consequences. It is worth noting that the subjects included in this early clinical trial had more advanced disease compared to the typical clinical trial GA population which may impact the generalizability of the findings to the broader GA population. Additionally, the study had a relatively small sample size, which increases the risk of bias. Furthermore, the biomarker conclusions drawn from the study subjects may not be representative of the subjects treated in the subsequent Phase II study. Therefore, caution should be exercised when generalizing the findings to larger populations. Despite convincing and robust genetic links to AMD reported for Type I *CFI* variants ^19–22^, this study observed no difference in biomarker response when comparing *CFI* RV carriers to the broad population. Interestingly, emerging data have revealed more modest ORs for *CFI* RVs in progression from early to late AMD ^23^ and macular thinning was observed in UK Biobank eyes of carriers <60 years old ^40^. Moreover, the long-term effects of complement therapy on the progression of GA are still uncertain. Another limitation to consider is the translatability of the *in vitro* VH modelling assay to the disease processes occurring in GA, and the relevance of the assay to cell-surface complement activation occurring in the area of GA. The LPS-coated surface used for the VH modelling assay would most likely result in more acute complement activation than in the GA retina. Finally, this is a relative potency assay and does not take into consideration the biodistribution of different modalities and routes of administration. Despite these limitations, the conclusions drawn from this study are compelling and hold significant implications for other gene therapy programs. The introduction of the innovative *in vitro* complement activation assay serves as a valuable benchmark for gene therapy programs.

The superior potency of FH compared to FI highlights the importance of conducting potency comparisons using *in vitro* assays that reflect physiologically relevant levels of complement proteins to guide decision-making. This finding opens promising avenues for further investigation, with potential for engineering of more active RCA protein truncates and chimeras including CR1, MCP, DAF, FH, and FHR dimerization domains. To advance gene therapy programs, future research should focus on building upon the pharmacokinetic / pharmacodynamic findings. It is crucial to establish tissue concentrations and estimates of the target required for complement modulation before moving into clinical stages. Pre-clinical assays that can be effectively translated to GA remain elusive. Overall, the role of complement in GA and the potential benefits of a safe one-time gene therapy in reducing AMD progression suggest the need for further clinical development of complement gene therapy.

## Data Availability

All data produced in the present study are available upon reasonable request to the authors

## Acknowledgements

The authors thank all the subjects who took part in the FOCUS study, and Gyroscope Therapeutics and Novartis colleagues past and present who made this possible.

The authors thank Michael Wittpoth, Les McGuire, Jayashree Sahni, and Maria Costa from Novartis for assistance in providing critical comments on the manuscript, and Shyamtanu Datta for assistance with the *in vitro* modelling.

## FOCUS principal investigators

Dr Claire Bailey
*Bristol Eye Hospital*
*Bristol, United Kingdom (UK)*

Professor Sobha Sivaprasad
*Moorfields Eye Hospital - NHS Foundation Trust London, UK*

Professor David Steel
*Sunderland Eye Infirmary*
*Sunderland, UK*

Dr Tsveta Ivanova
*Manchester Royal Eye Hospital*
*Manchester, UK*

Dr Paulo Stanga
*The Retina Clinic London*
*London, UK*

Dr Kamin Xue
*Nuffield Laboratory of Ophthalmology University of Oxford & Oxford Eye Hospital, Oxford University Hospitals NHS Foundation Trust*
*Oxford, UK*

Dr Peter Charbel Issa
*Nuffield Laboratory of Ophthalmology*
*University of Oxford & Oxford Eye Hospital, Oxford University Hospitals NHS Foundation Trust*
*Oxford, UK*

Dr Jared Neilsen
*Wolfe Eye Clinic West Des Moines, Indianapolis, USA*

Dr Jeff Heier
*Ophthalamic Consultants of Boston*
*Boston, USA*

Dr Arshad Khanani
*Sierra Eye Associates*
*Reno, USA*

Dr Christopher Riemann
*Cincinnati Eye Institute Cincinnati, Ohio, US*

Dr Alan Ho
*Mid-Atlantic Retina, Wills Eye Hospital Philadelphia, Pennsylvania, US*

Dr Raj Maturi
*Midwest Eye Institute Northside*
*Indianapolis, US*

Dr Anthony De Beus
*Pepose Vision Institute Chesterfield, Missouri, US*

Prof. Robert E MacLaren
*Nuffield Laboratory of Ophthalmology*
*University of Oxford & Oxford Eye Hospital, Oxford University Hospitals NHS Foundation Trust*

Dr Nancy Holekamp
*Pepose Vision Institute Chesterfield, Missouri, US*

## ABBREVIATIONS

AEs: Adverse events
AH: Aqueous humor
AMD: Age-related macular degeneration
AP: Alternative Pathway
BL: Baseline
C3: Complement component 3
cDNA: Complementary deoxyribonucleic acid
CR1: Complement receptor 1
CS: Complement system
DAF: Decay-accelerating factor
EDTA: Ethylenediaminetetraacetic acid
FAF: Fundus autofluorescence
FB: Factor B
FD: Factor D
FDA: Food and Drug Administration
fdr: False discovery rate
FH: Factor H
FI: Factor I
GA: Geographic atrophy
GMP: Good Manufacturing Process
IVT: Intravitreal
LLOQ: Lower limit of quantification
LOD: Limit of detection
LPS: Lipopolysaccharide
MBL: Mannose binding lectin
MCP: Membrane cofactor protein
NPX: Normalized protein expression
PK: Pharmacokinetics
RCA: Regulators of complement activation
RPE: Retinal pigment epithelium
RV: Rare variant
SDS: Subretinal delivery system
TP: Total protein
TVSI: Transvitreal surgical intervention
UK: United Kingdom
ULOQ: Upper limit of quantification
US: United States
VH: Vitreous humor

## Supplementary Methods

### Study design

FOCUS was conducted at 13 sites over two countries and was sponsored by Gyroscope Therapeutics, a Novartis company. Key inclusion criteria included a clinical diagnosis of GA secondary to AMD in the study eye as determined by a retinal specialist, and a diagnosis of AMD in the contralateral eye (except if subject was monocular). Study eye GA lesion(s) total size was ≥1.25mm^2^ and ≤17.5mm^2^ (cohort 1 to 6) or ≥1.25mm^2^ (cohort 7), and GA lesions resided completely within the fundus autofluorescence (FAF) fundus image. Key exclusion criteria included presence of the following in the study eye; CNV, non-proliferative diabetic retinopathy, glaucomatous optic neuropathy, or cataract. Subjects were also excluded if they had an active malignancy or a history of vitrectomy, sub-macular surgery, macular photocoagulation or previously receiving a gene or cell therapy in the study eye.

As outlined in **Supplementary Table 1,** the study had four parts, part 1 (cohorts 1-3); dose escalation, Part 2 (cohort 4); dose expansion, Part 3 (cohorts 5-6); dose escalation with the Orbit Sub-retinal Delivery System (SDS), and Part 4 (cohort 7); dose-expansion with the Orbit SDS.

PPY988 gene therapy aimed to transduce RPE and PR cells, considered a key site for AMD pathology and FI production. Parts 1 and 2 (cohorts 1-4) delivered PPY988 via standard transvitreal subretinal injection (TVSI), previously used for subretinal gene therapy delivery ^1,2^. The surgeon then performed the TVSI procedure involving a standard 3-port pars plana vitrectomy. The retina was partially detached through a cannula using buffered salt solution for bleb formation, and a fixed volume of 0.1mL containing the gene therapy dose was injected into the bleb, which was expected to flatten after 24 hours.

Subjects in Parts 3 and 5 (cohorts 5-7) received PPY988 subretinally via suprachoroidal cannulation using Orbit SDS, a 510(k) cleared device in the US (K200325, 14-Jul-2020, US FDA), developed to improve the safety profile of subretinal injections as compared with TVSI ^3^. However, because VH samples were not collected in Parts 3 to 5, analysis of biomarker data from subjects treated in these cohorts is not discussed herein.

VH, AH and plasma samples were collected from cohort 1 to 4 subjects at BL prior to surgery, at weeks 12, 36 and 96 after dosing, except for sample collection at week 5 (not week 12) in cohorts 1 and 2, and AH (not VH) was collected for cohort 4 at Week 12.

Approximately 25 to 100µL of AH and VH samples were collected under microscopic visualization. AH samples were collected using a 30G needle attached to a 1mL syringe. The 30G needle was entered into the anterior chamber through the temporal limbus using a clear corneal approach. VH samples were collected at BL using a non-primed 23G vitrectomy probe connected to a 1mL syringe, and post-BL using a 30G needle through the *pars plana*. Immediately following collection VH and AH samples were transferred to a sterile collection vial and frozen at –80°C within 30 minutes.

All subjects were assessed for the occurrence of adverse events (AEs) at each follow-up visit and underwent functional visual and retinal imaging/anatomical assessments and biological sampling for safety and immunogenicity monitoring at periodic timepoints up to week 48, considered the primary endpoint timepoint of the study.

It was intended that subjects would be invited to consent to an additional long-term follow-up assessment until week 240 (5 years). At the time of Phase II study termination, FOCUS was fully enrolled. As a consequence of the early termination of the Phase II study, all ocular sampling in FOCUS from that point onward was terminated, and subjects who had not completed the study assessment window to week 48 were transferred onto a safety monitoring regimen.

### Olink proteomics

VH samples from 17 subjects were analyzed using the OLINK^®^ Target 96 platform (OLINK^®^ Proteomics, Uppsala, Sweden) to identify broader pathway changes affected by subretinal delivery of PPY988. A single measurement was performed for each sample. The following 4 panels were assessed by Olink: inflammation and inflammation II, cardiometabolic and neurology. Protein levels were measured on a relative scale and presented as the normalized protein expression (NPX), which is an arbitrary unit on a log2 scale. Principal component analysis was performed to detect outlier samples. Olink provides a limit of detection (LOD) for each protein based on the background noise generated in negative controls (buffer run as a normal sample). Nonetheless, the Olink protein expression data reported NPX values even for measurements below the LOD. Low abundance proteins were excluded from the statistical analyses. To identify the proteins changing over time relative to BL, a linear mixed effect model was used. The model included the following covariates: timepoint, subject age and sex, sample total protein content and *CFI* RV status (subject ID as random effect). All analyses were carried out using R version 4.2.3. Nominal p-values were adjusted using false discovery rate.

### *In vitro* FI potency comparability assay

HEK293 adherent cells with the *CFI* and *CFH* genes knocked out by CRISPR-Cas9 were transduced with PPY988 (AAV2-*CFI*) vector material produced under good manufacturing practice (GMP) at 10 different MOIs. After a 24-hour incubation (37°C at 5% CO_­_) with the vector, the media was replaced and left for a further 48 hours. Supernatants were collected and stored at −20°C. Supernatants were thawed, centrifuged (5000xG for 15 mins) then mixed with a fixed volume of human purified C3b and FH (A114 and A137, Comptech) and incubated for 10 minutes at 37°C. The mixture was then reduced and labelled by incubating for 5 minutes at 90°C with a mixture of DTT, SDS buffer and the LIF dye Chromeo P503 (30693, Sigma). A standard curve of Purified FI (Comptech, Tyler, TX) was made starting at a final concentration of 4000 ng/mL down to 11 ng/mL. This was diluted in non-transduced control sample media to ensure the same background matrix for both transduction supernatants samples and Purified FI standard curve samples. Proteins were separated by CE-SDS using LIF detection and an adapted version of a published method ^4^. The areas of the peak corresponding 114 kDa alpha prime subunit (normalised to the 75 kDa beta subunit) are collected for both the Reference (Comptech Purified FI) and the Test sample (AAV2-*CFI* GMP). The depletion of the peak of interest is plotted using QuBAS 3.0 Statistical Software (Quantics Biostatistics) which performs a parallel line comparison of the Test compared to the reference and produces a Relative Potency value corresponding to this analysis. Outliers are statistically identified by a Grubbs Test and removed automatically from the analysis prior to calculating the Relative Potency.

---

**Supplementary Figure 1.**
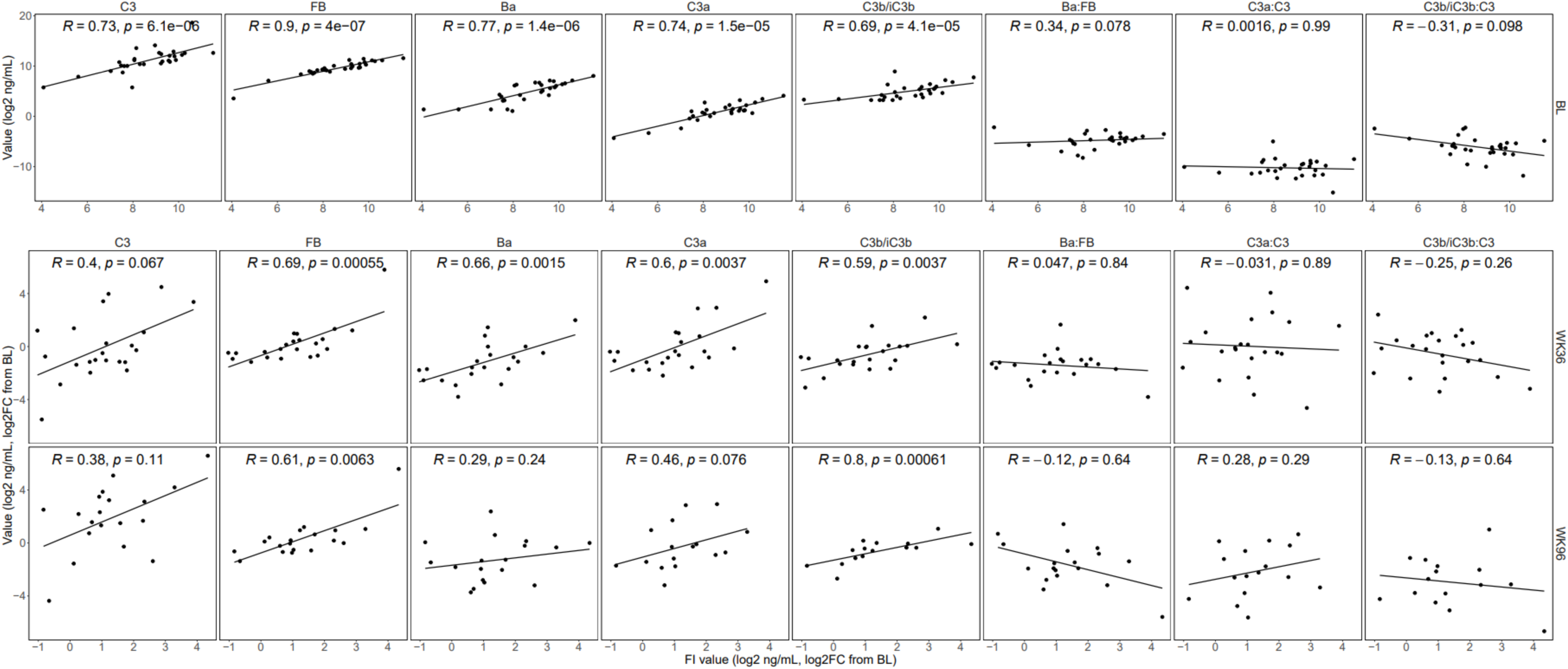
VH complement biomarker pairwise correlation analysis. Pairwise correlation analysis of FI with intact complement proteins and breakdown products and ratios in the vitreous at baseline, week 36 and week 96 from subjects in cohorts 1 to 4. FI concentrations are shown by the x-axis while biomarker concentrations and ratios are shown by the y-axis. Correlations at BL are determined using log2 ng/mL concentrations, correlations at weeks 36 and 96 are determined using log2 fold change from BL. BL = Baseline; WK = week; FC = Fold Change; R = Spearman Correlation Coefficient; p = p-value.

**Supplementary Figure 2.**
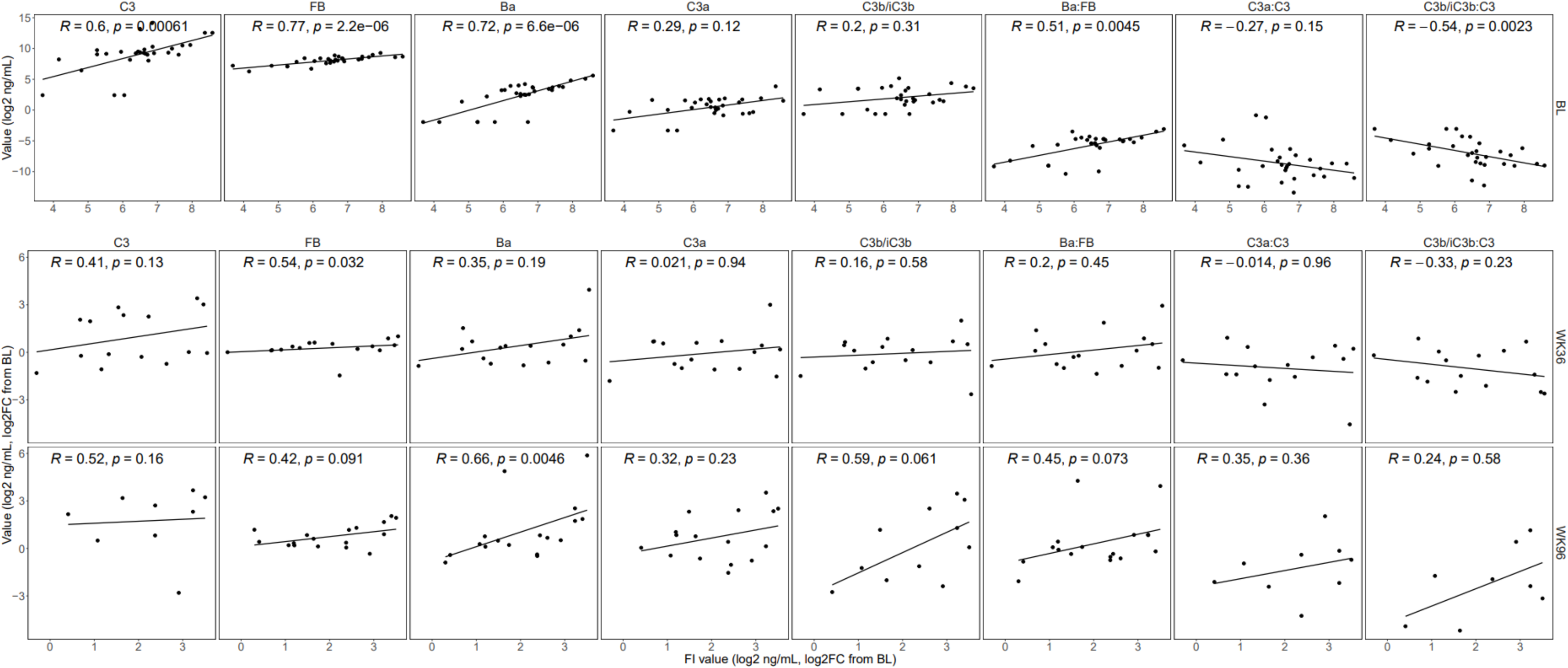
AH complement biomarker pairwise correlation analysis. Pairwise correlation analysis of FI with intact complement proteins and breakdown products and ratios in the aqueous humor at baseline, week 36 and week 96 from subjects in cohorts 1 to 4. FI concentrations are shown by the X-axis while biomarker concentrations and ratios are shown by the y-axis. Correlations at BL are determined using log2 ng/mL concentrations, correlations at weeks 36 and 96 are determined using log2 fold change from BL. BL = Baseline; WK = week; FC = Fold Change; R = Spearman Correlation Coefficient; p = p-value.

**Supplementary Figure 3.**
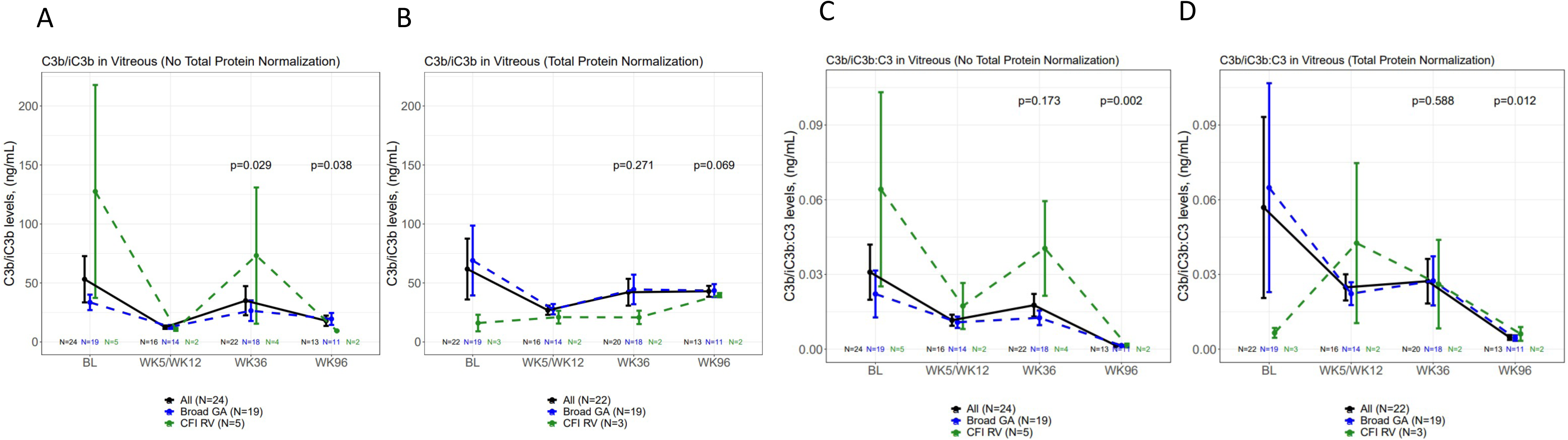
Modulation of C3b/iC3b and C3b/iC3b:C3 by PPY988 delivery. (A,B) C3b/iC3b concentrations (y-axis, ng/mL) in VH samples by week post-surgery (x-axis) from subjects in cohorts 1 to 4. Mean +/− SE is shown by bars for all (black lines) broad (blue lines) and CFI RV (green lines) groups. Significant reductions in C3b/iC3b were observed at week 36 for both non-normalised (P=0.029) (A) but not total protein (TP) normalised (P=0.271) (B) data, and at week 96 for non-normalised (P=0.038) but not TP normalised (P=0.069) data. Notably these reductions are modest compared to for Ba modulation. (C,D) Ratios of C3b/iC3b:C3 concentration (Y-axis, ng/mL) were calculated for each subject at each sampling post-surgery week (X-axis). Mean +/− SE is shown by bars for all (black lines) broad (blue lines) and CFI RV (green lines) groups. No reductions were observed at week 36 for either non-normalised (P=0.173) (C) or TP normalised (P=0.588) (D) C3b/iC3b:C3 ratios. However, reductions were observed at week 96 for non-normalized (P=0.002) and TP normalized (P=0.012) C3b/iC3b:c3 ratios. Significance is defined as *P<0.05; **P<0.01, ***P<0.001. N numbers are shown above X-axis. BL = Baseline; WK= week.

**Supplementary Figure 4.**
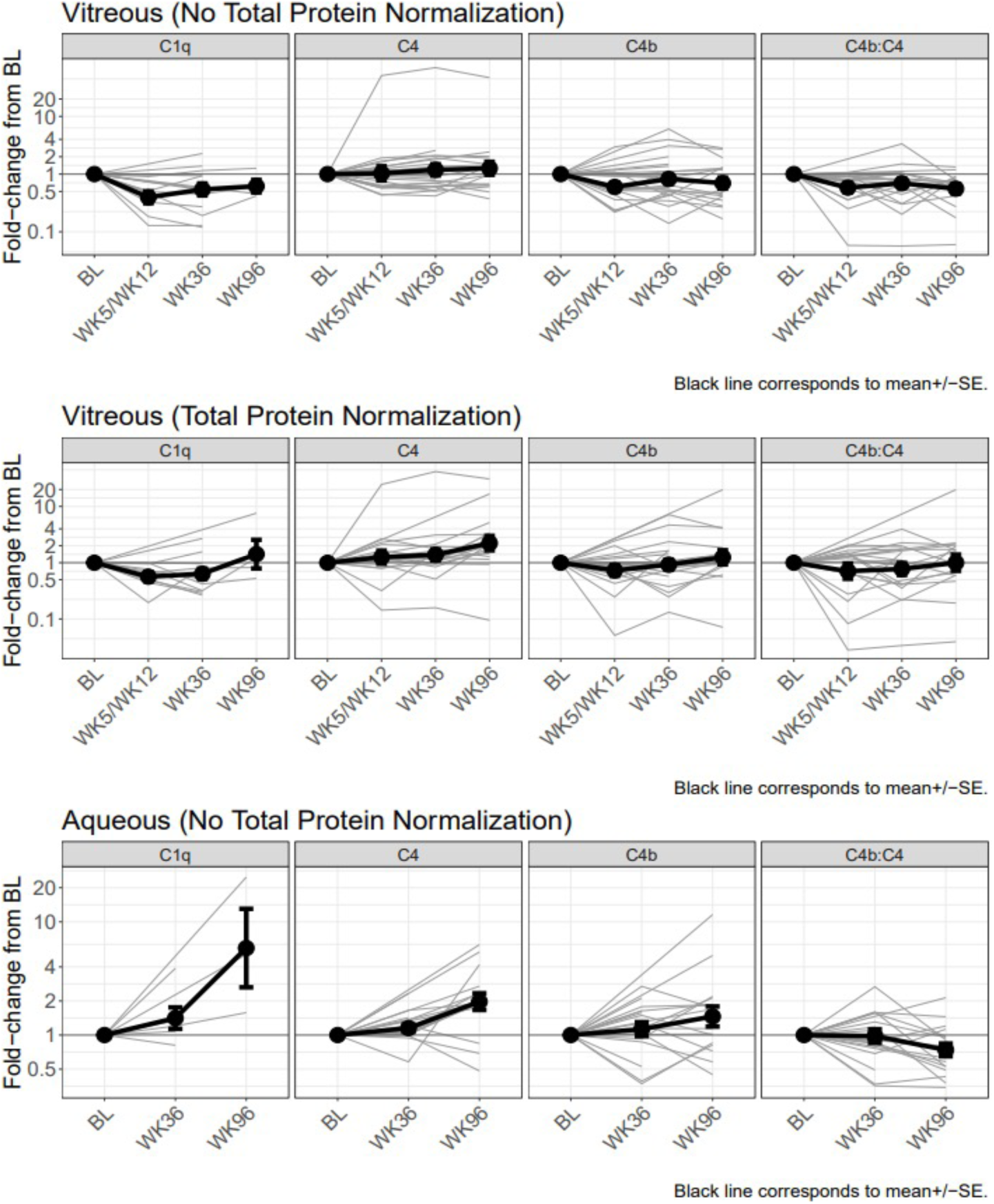
Classical pathway (CP) engagement post-treatment with PPY988. Fold-changes (y-axis) in CP complement biomarkers [C1q, C4, C4b and C4b:C4] concentrations from baseline at each post-surgery week (VH: week 5/12, 36 and 96; AH: week 36 and 96; X-axis) are shown in VH with no normalisation (A), VH with total protein normalisation (B), and AH with no normalisation. Mean is shown by thick black lines, +/− SE is shown by bars. Individual subject changes are shown by grey lines. BL = Baseline; WK= week.

**Supplementary Figure 5.**
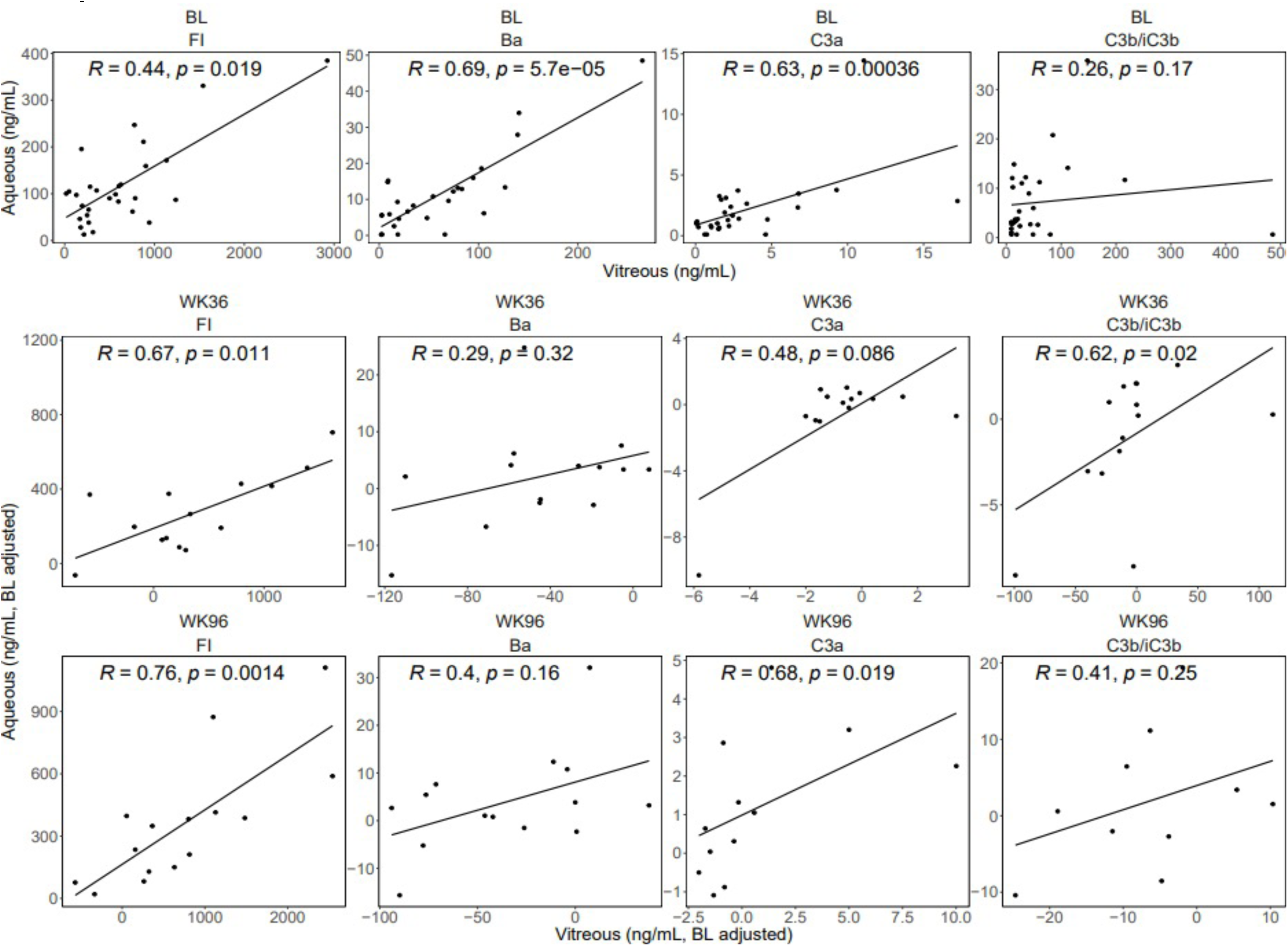
Pairwise correlation analysis of vitreous versus aqueous humor complement biomarkers at baseline and post-treatment from subjects in cohorts 1 to 4. VH concentrations on x-axis and AH concentrations on y-axis of FI, Ba, C3a and C3b/iC3b at baseline, week 36 and week 96 post-surgery. Vitreous and aqueous concentrations of FI and C3a significantly correlate at all time points, Ba concentrations correlate only at baseline, while C3b/iC3b concentrations only correlate significantly at week 36 post-treatment. BL = Baseline; WK = week; R = Spearman Correlation Coefficient; *p* = *p-value*.

**Supplementary Figure 6.**
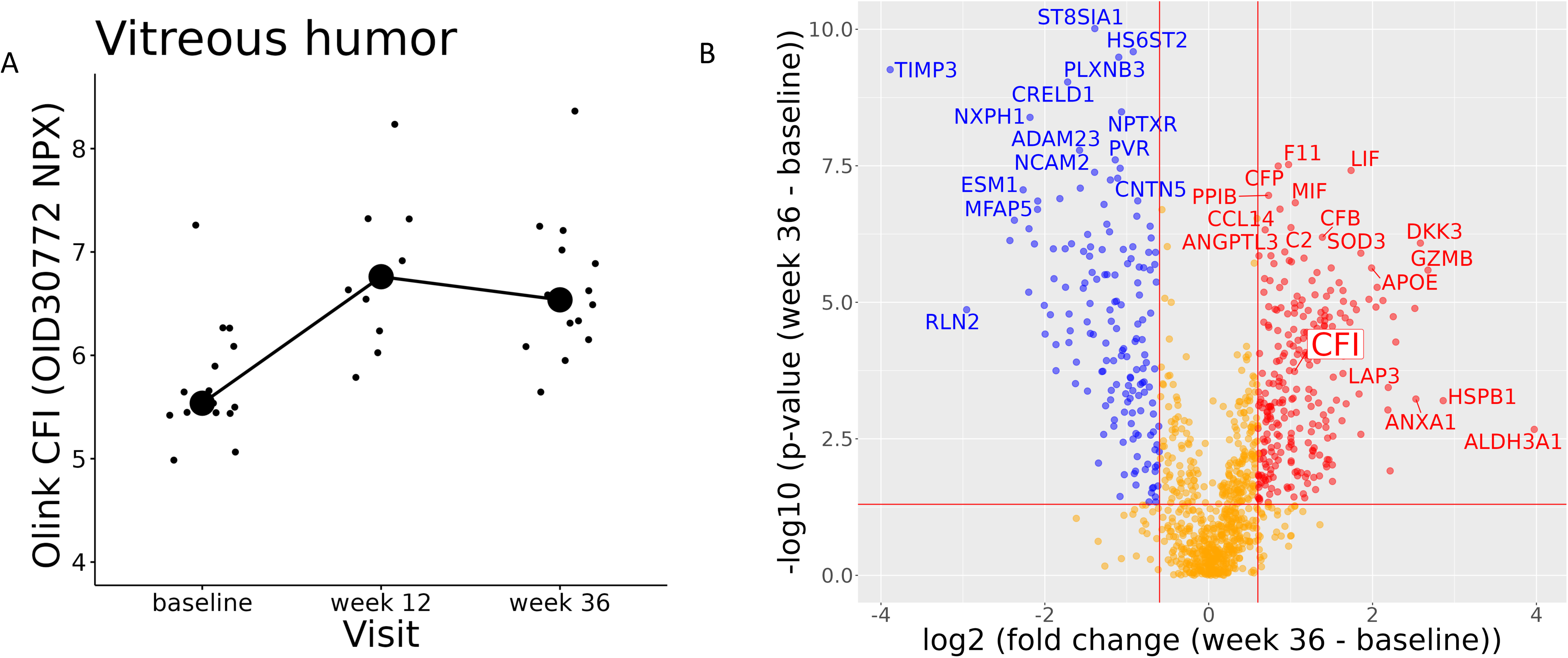
Proteomic analysis of vitreous humor using Olink. **(A)** Change in FI levels (Olink OID30772) over time. Median FI values measured at baseline, week 12, and week 36 are linked. Sample numbers were baseline; n=17, week 12; n=14, week 36; n=17. (B) Volcano plot of differential protein expression (week 36 relative to baseline). Minimum protein fold-change was set at absolute 1.5. Data points with a nominal p-value of less than 0.05 were considered statistically significant (red dots indicate significantly upregulated proteins, blue dots indicate significantly downregulated proteins and orange dots represent no significant change on a log2 scale).

**Supplementary Figure 7.**
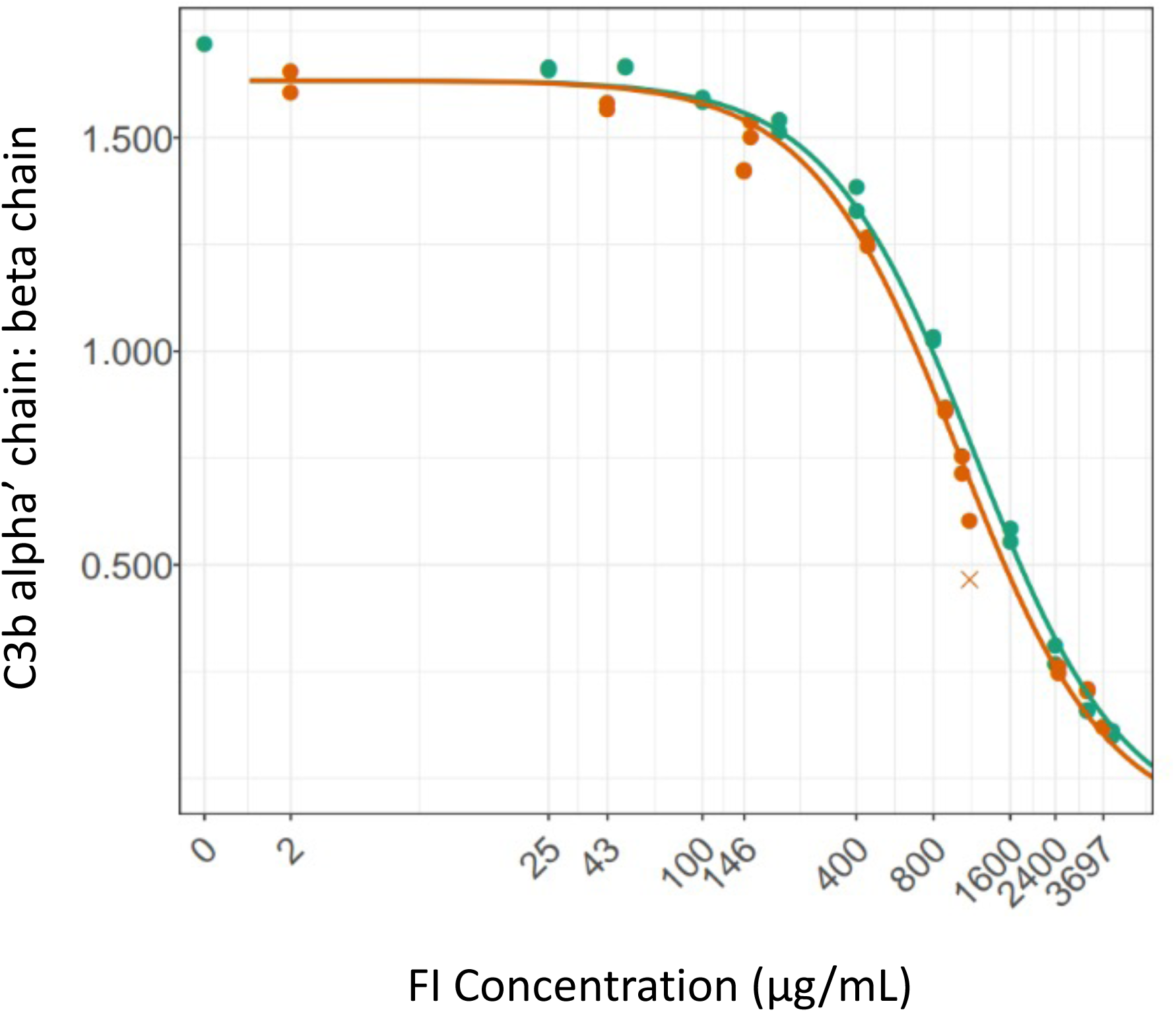
Comparability of commercial FI to AAV2-*CFI* potency. C3b alpha’ chain breakdown normalised to C3b intact beta chain (y-axis) produced by increasing concentrations of PPY988 (GMP#3) in orange compared to commercial FI (Complement Technology (CompTech), Inc) in green. The serum purified CompTech FI protein was diluted in non-transduced control sample so that both FI (reference sample) and PPY988 FI (test sample) were in the same background matrix to ensure comparability. No difference in potency was observed between PPY988 and CompTech FI Protein. Relative Potency of PPY988**: 1.151;** 95% CI range: 1.065-1.244; Precision: 1.168. See supplemental methods for further details.

**Supplementary Table 1.** FOCUS study overview. FOCUS consisted of four parts of either PPY988 dose escalation or expansion, divided by seven cohorts varying according to route of gene therapy administration, dose (vg; viral genomes), and biomarker matrix. Only Parts 1 and 2 (cohorts 1 to 4) are described given subretinal gene therapy delivery permitted vitreous humour (VH) sampling. VH, aqueous humour (AH) and plasma biomarker samples were collected from subjects prior to surgery at baseline (BL), and subsequent timepoints up to week 96 post-treatment. Emerging data and analysis on cohorts 1 and 2 indicated week 5 sampling from the VH was too early to evaluate an impact post-vitrectomy, so the first post-treatment evaluation for cohorts 3 and 4 was moved to a later time-point in the study (week 12), when it was expected that the proteome would have stabilised, and the transduced cells achieved maximal expression.

| Part (name) | Cohort | SR delivery method | PPY988 dose [vg/eye] | Subjects (n) | Biomarker matrix; sampling timepoints |
| --- | --- | --- | --- | --- | --- |
| 1 (dose escalation) | 1 | Standard | low, 2E10 | 3 | VH; BL, week 5, 36, 96<br>AH; BL, week 36, 96<br>Plasma: BL, week 5, 36, 96 |
|  | 2 |  | mid, 5E10 | 4 | VH; BL, week 5, 36, 96<br>AH; BL, week 36, 96<br>Plasma: BL, week 12, 36, 96 |
|  | 3 |  | high, 2E11 | 4 | VH, BL, week 12, 36, 96<br>AH; BL, week 36, 96<br>Plasma: BL, week 12, 36, 96 |
| 2 (dose expansion) | 4 | Standard | low, 2E10 (n=3);<br>mid, 5E10 (n=6);<br>high, 2E11 (n=11) | 20 | VH, BL, week 12, 36, 96<br>AH; BL, week 36, 96<br>Plasma: BL, week 12, 36, 96 |

**Supplementary Table 2.** Number of FOCUS samples analysed longitudinally across study. VH and AH samples from subjects in cohorts 1 to 4 that passed acceptance criteria taken at baseline (BL) and at each timepoint post treatment are provided. Only those subjects with sampling at both BL and at least one of week 36 or 96 time-points are included.

| Visit | FI | C3 | FB | FH | C1q | Ba | C3a | C3b/iC3b | C4 | C4b | Ba:FB | C3a:C3 | C3b/iC3b:C3 | C4b:C4 | C5 | FD | MBL |
| --- | --- | --- | --- | --- | --- | --- | --- | --- | --- | --- | --- | --- | --- | --- | --- | --- | --- |
| BL/pre-dosing | 24 | 24 | 24 | 24 | 15 | 23 | 24 | 24 | 24 | 24 | 23 | 24 | 24 | 24 | 23 | 24 | 24 |
| Week 5/12 | 16 | 16 | 16 | 16 | 8 | 16 | 16 | 16 | 16 | 16 | 16 | 16 | 16 | 16 | 15 | 16 | 16 |
| Week 36 | 22 | 22 | 22 | 22 | 14 | 21 | 22 | 22 | 22 | 22 | 21 | 22 | 22 | 22 | 21 | 22 | 22 |
| Week 96 | 17 | 17 | 17 | 17 | 4 | 16 | 15 | 13 | 17 | 16 | 16 | 15 | 13 | 16 | 16 | 17 | 17 |

Aqueous
| Visit | FI | C3 | FB | FH | C1q | Ba | C3a | C3b/iC3b | C4 | C4b | Ba:FB | C3a:C3 | C3b/iC3b:C3 | C4b:C4 | C5 | FD | MBL |
| --- | --- | --- | --- | --- | --- | --- | --- | --- | --- | --- | --- | --- | --- | --- | --- | --- | --- |
| BL/pre-dosing | 20 | 19 | 20 | 20 | 8 | 20 | 20 | 19 | 20 | 20 | 20 | 19 | 19 | 20 | 19 | 20 | 20 |
| Week 5/12 | 0 | 0 | 0 | 0 | 0 | 0 | 0 | 0 | 0 | 0 | 0 | 0 | 0 | 0 | 0 | 0 | 0 |
| Week 36 | 16 | 15 | 16 | 16 | 6 | 16 | 16 | 15 | 16 | 16 | 16 | 15 | 15 | 16 | 16 | 16 | 16 |
| Week 96 | 16 | 8 | 16 | 16 | 3 | 16 | 15 | 10 | 16 | 16 | 16 | 8 | 7 | 16 | 15 | 16 | 16 |

Plasma
| Visit | FI | C3 | FB | FH | C1q | Ba | C3a | C3b/iC3b | C4 | C4b | Ba:FB | C3a:C3 | C3b/iC3b:C3 | C4b:C4 | C5 | FD | MBL |
| --- | --- | --- | --- | --- | --- | --- | --- | --- | --- | --- | --- | --- | --- | --- | --- | --- | --- |
| BL/pre-dosing | 15 | 16 | 16 | 16 | 9 | 20 | 13 | 12 | 16 | 17 | 13 | 13 | 12 | 16 | 17 | 15 | 17 |
| Week 5/12 | 0 | 0 | 0 | 0 | 0 | 0 | 0 | 0 | 0 | 0 | 0 | 0 | 0 | 0 | 0 | 0 | 0 |
| Week 36 | 15 | 16 | 16 | 16 | 9 | 20 | 13 | 12 | 16 | 17 | 13 | 13 | 12 | 16 | 17 | 15 | 17 |
| Week 96 | 0 | 0 | 0 | 0 | 0 | 0 | 0 | 0 | 0 | 0 | 0 | 0 | 0 | 0 | 0 | 0 | 0 |

**Supplementary Table 3.** Descriptive statistics of vitreous humour baseline complement biomarkers concentrations from subjects in cohorts 1 to 4. The number of samples correspond to the total number which could be measured and had a value within acceptance criteria. All concentrations shown are expressed in ng/mL.

S Table 3. VH Complement Biomarker baseline descriptive stats (ng/mL)
| Biomarker | n | Mean | SD | Median | 25<sup>th</sup> Percentile | 75<sup>th</sup> Percentile | Minimum | Maximum | LOD | N LOD | ULOQ | N ULOQ |
| --- | --- | --- | --- | --- | --- | --- | --- | --- | --- | --- | --- | --- |
| FI | 29 | 608.88 | 587.42 | 499.91 | 215 | 785.62 | 16.91 | 2924.77 | 4.625 | 0 | 12500 | 0 |
| C3 | 29 | 17350.67 | 74622.03 | 2163.41 | 1104.46 | 5344.56 | 53.7 | 404817.6 | 53.7 | 2 | 60000 | 0 |
| FB | 29 | 1083.14 | 817.41 | 765 | 504.85 | 1402.1 | 11.81 | 3037.52 | 5.8 | 0 | 6000 | 0 |
| FH | 29 | 466.7 | 427.59 | 273.56 | 181.66 | 652.73 | 20.7 | 1430.34 | 20.7 | 1 | 30000 | 0 |
| C1q | 19 | 140.45 | 106.43 | 126.07 | 70.85 | 179.48 | 18.48 | 478.34 | 18.48 | 2 | 7200 | 0 |
| Ba | 28 | 58.07 | 60.43 | 41.3 | 10.06 | 86.07 | 2.08 | 264.86 | 2.6 | 4 | 416 | 0 |
| C3a | 28 | 3.36 | 3.83 | 2.06 | 1.34 | 3.68 | 0.05 | 17.23 | 0.05 | 1 | 125 | 0 |
| C3b/iC3b | 29 | 59.01 | 94.58 | 24.97 | 13.68 | 57.56 | 9.4 | 485.39 | 9.405 | 4 | 15000 | 0 |
| C4 | 29 | 2488.05 | 1998.79 | 1929.44 | 810.08 | 3472.82 | 31.51 | 7185.16 | 28.8 | 0 | 40000 | 0 |
| C4b | 29 | 130.76 | 137.09 | 112 | 17.49 | 180.12 | 3.77 | 640.53 | 15.55 | 3 | 25000 | 0 |
| Ba:FB | 28 | 0.05 | 0.05 | 0.04 | 0.02 | 0.06 | 0 | 0.22 | NA | 0 | NA | 0 |
| C3a:C3 | 28 | 0 | 0.01 | 0 | 0 | 0 | 0 | 0.03 | NA | 0 | NA | 0 |
| C3b/iC3b:C3 | 29 | 0.04 | 0.06 | 0.01 | 0.01 | 0.03 | 0 | 0.21 | NA | 0 | NA | 0 |
| C4b:C4 | 29 | 0.06 | 0.09 | 0.04 | 0.02 | 0.07 | 0 | 0.49 | NA | 0 | NA | 0 |
| C5 | 27 | 322.23 | 412.28 | 137 | 137 | 320.05 | 137 | 2022 | 137 | 14 | 100000 | 0 |
| FD | 29 | 98.45 | 116.77 | 59.43 | 41.42 | 113.73 | 4.81 | 600.14 | 0.8625 | 0 | 625 | 0 |
| MBL | 29 | 0.83 | 0.44 | 0.62 | 0.62 | 0.86 | 0.3 | 2.31 | 0.625 | 15 | 1250 | 0 |

**Supplementary Table 4.**
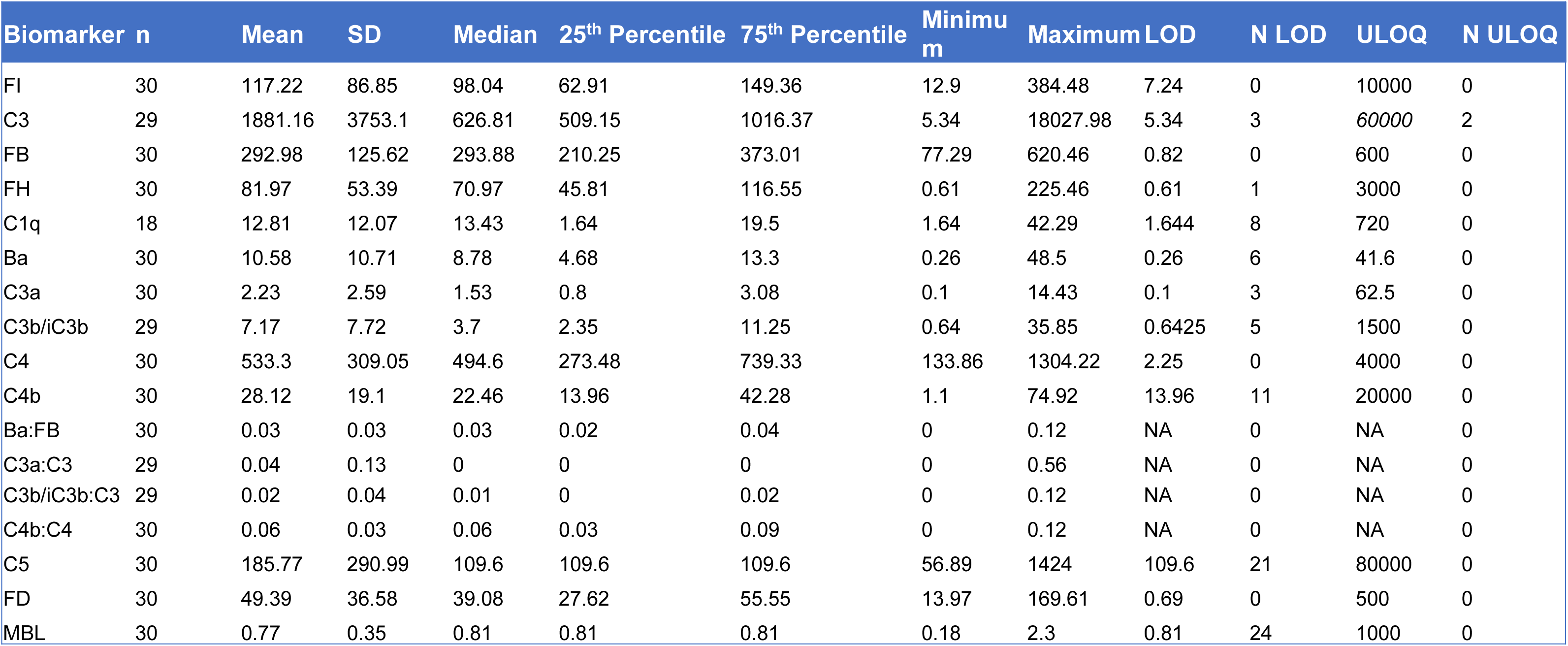
Descriptive statistics of aqueous humour baseline complement biomarkers concentrations from subjects in cohorts 1 to 4. The number of samples correspond to the total number which could be measured and had a value within acceptance criteria. All concentrations shown are expressed in ng/mL.

**Supplementary Table 5.**
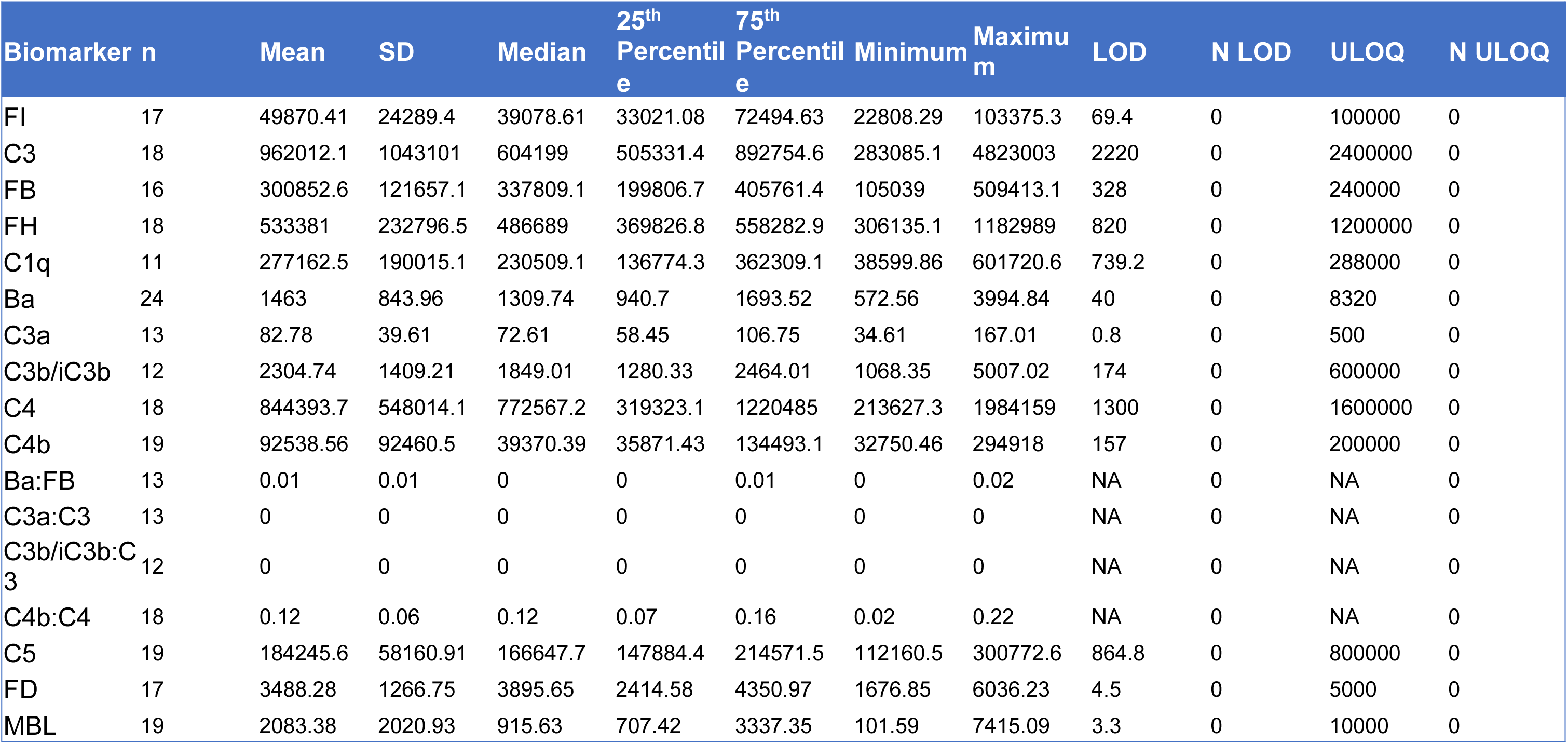
Descriptive statistics of plasma baseline complement biomarkers concentrations from subjects in cohorts 1 to 4. The number of samples correspond to the total number which could be measured and had a value within acceptance criteria. All concentrations shown are expressed in ng/mL.

**Supplementary Table 6.**
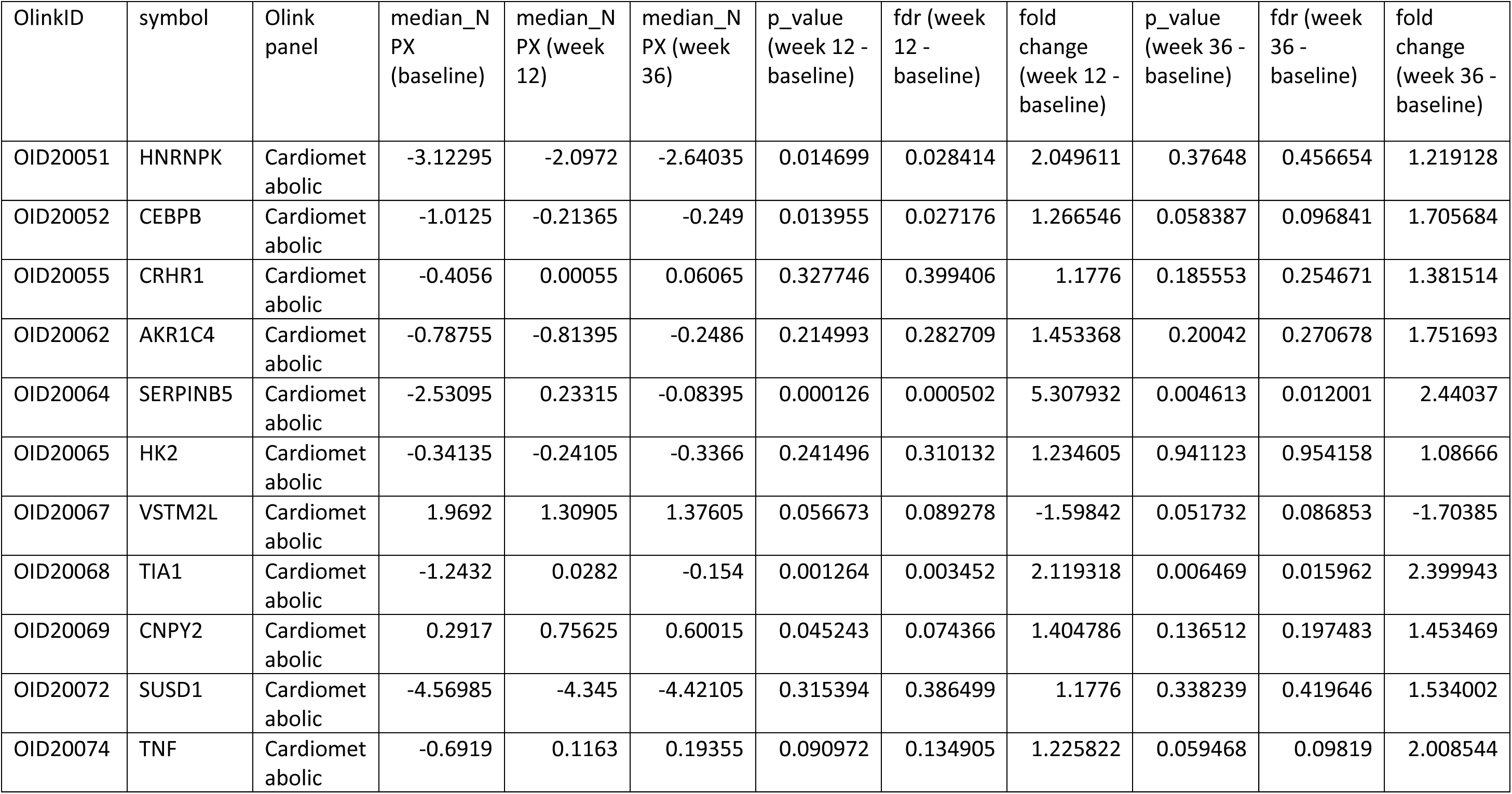
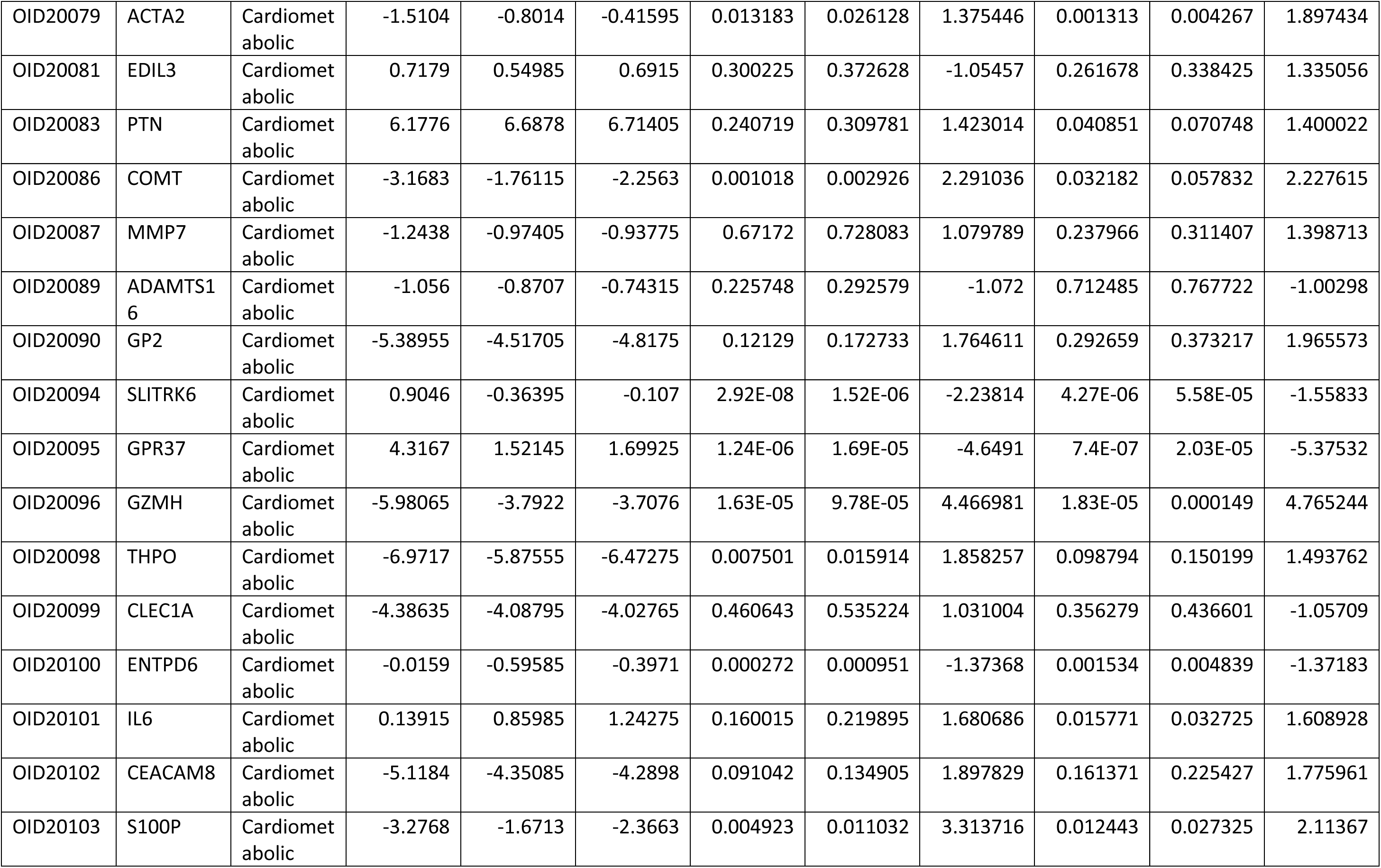
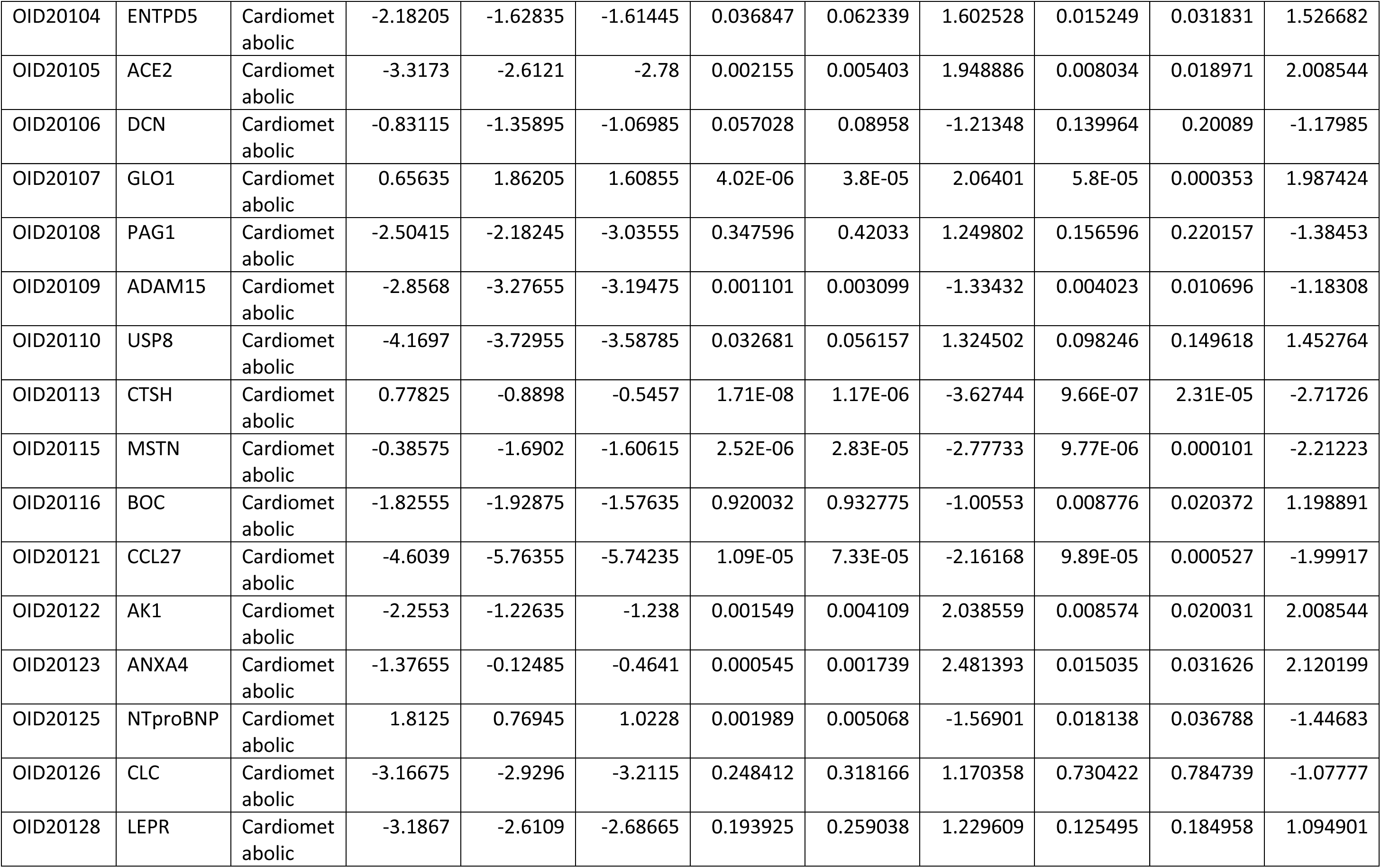
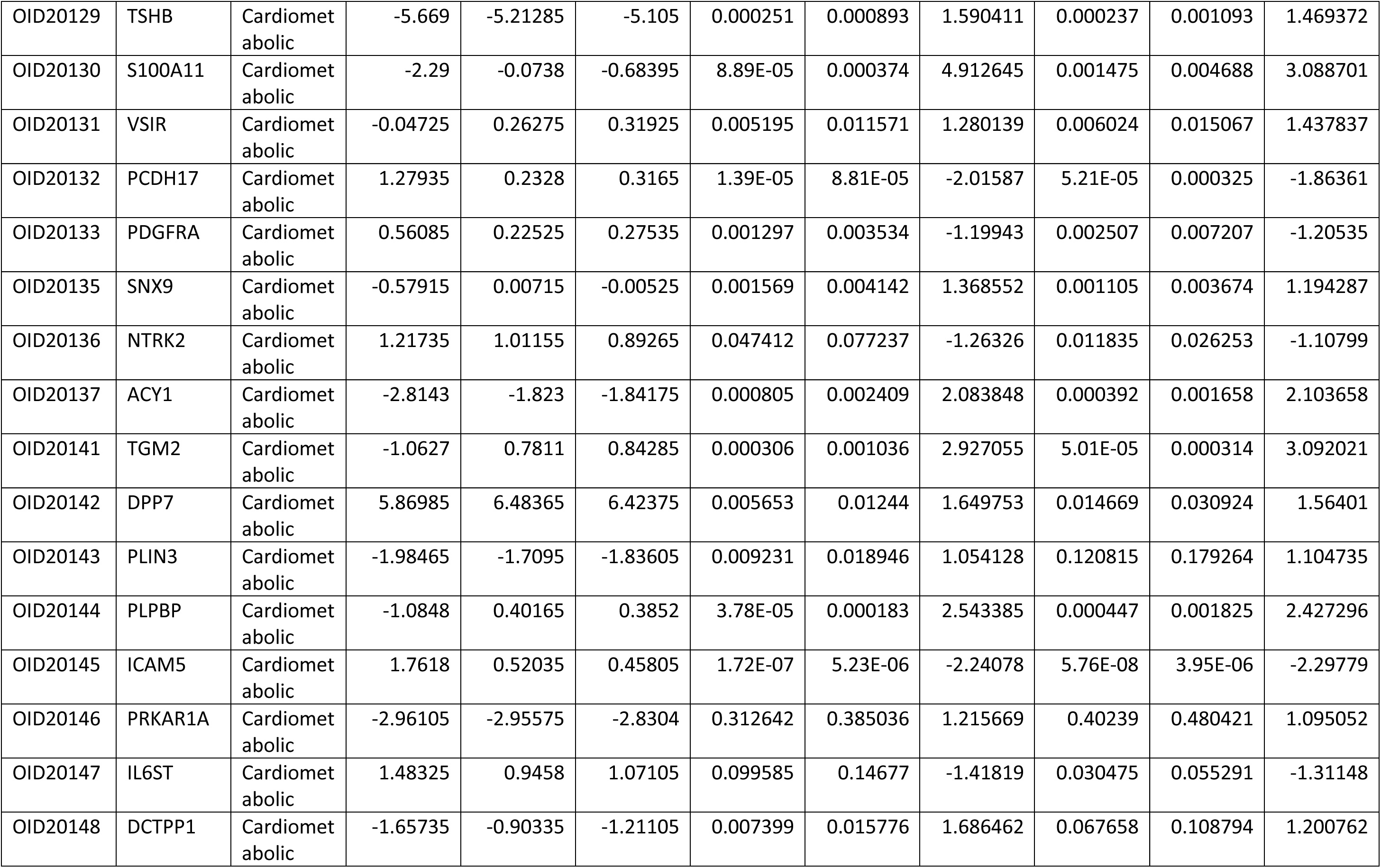
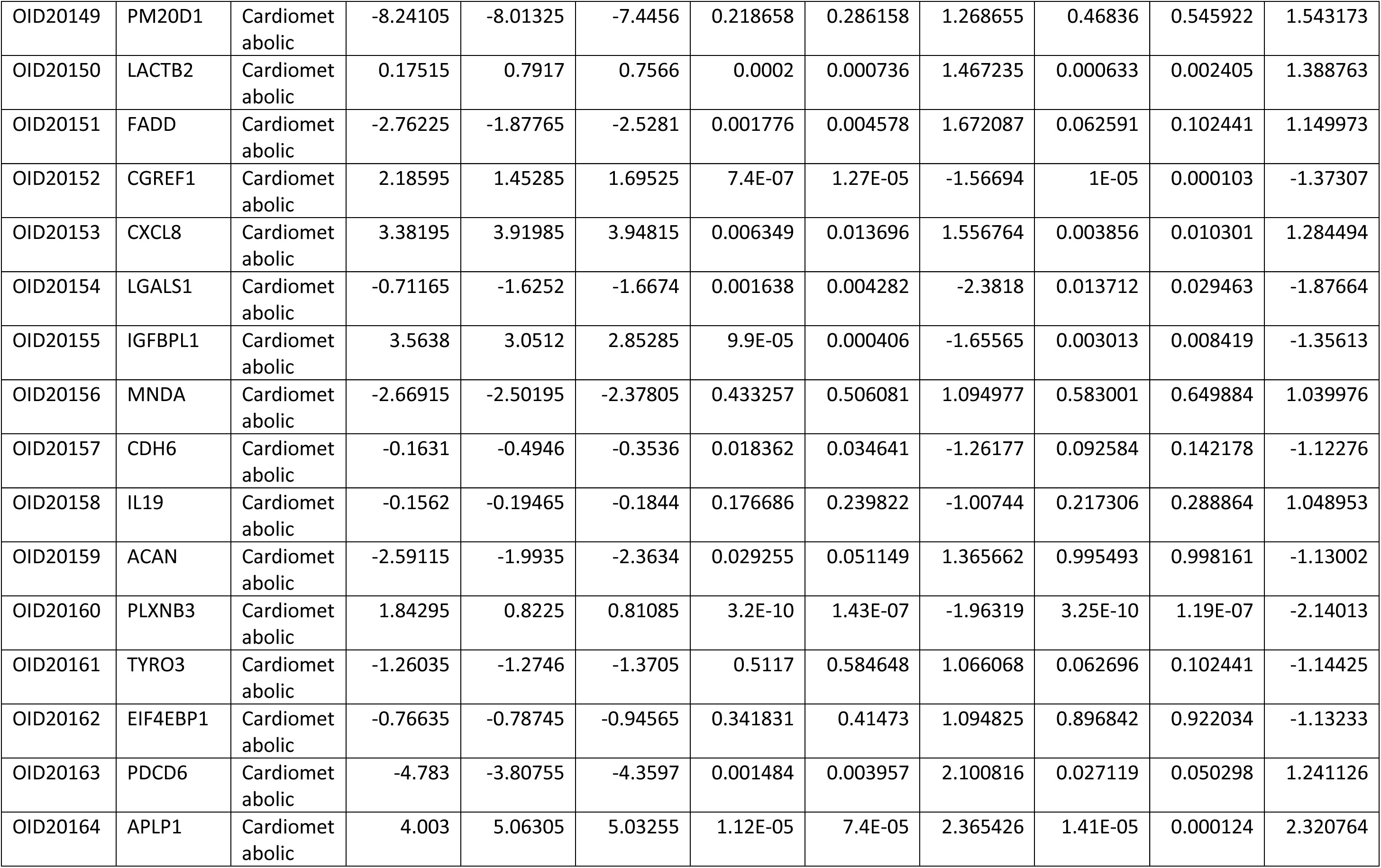
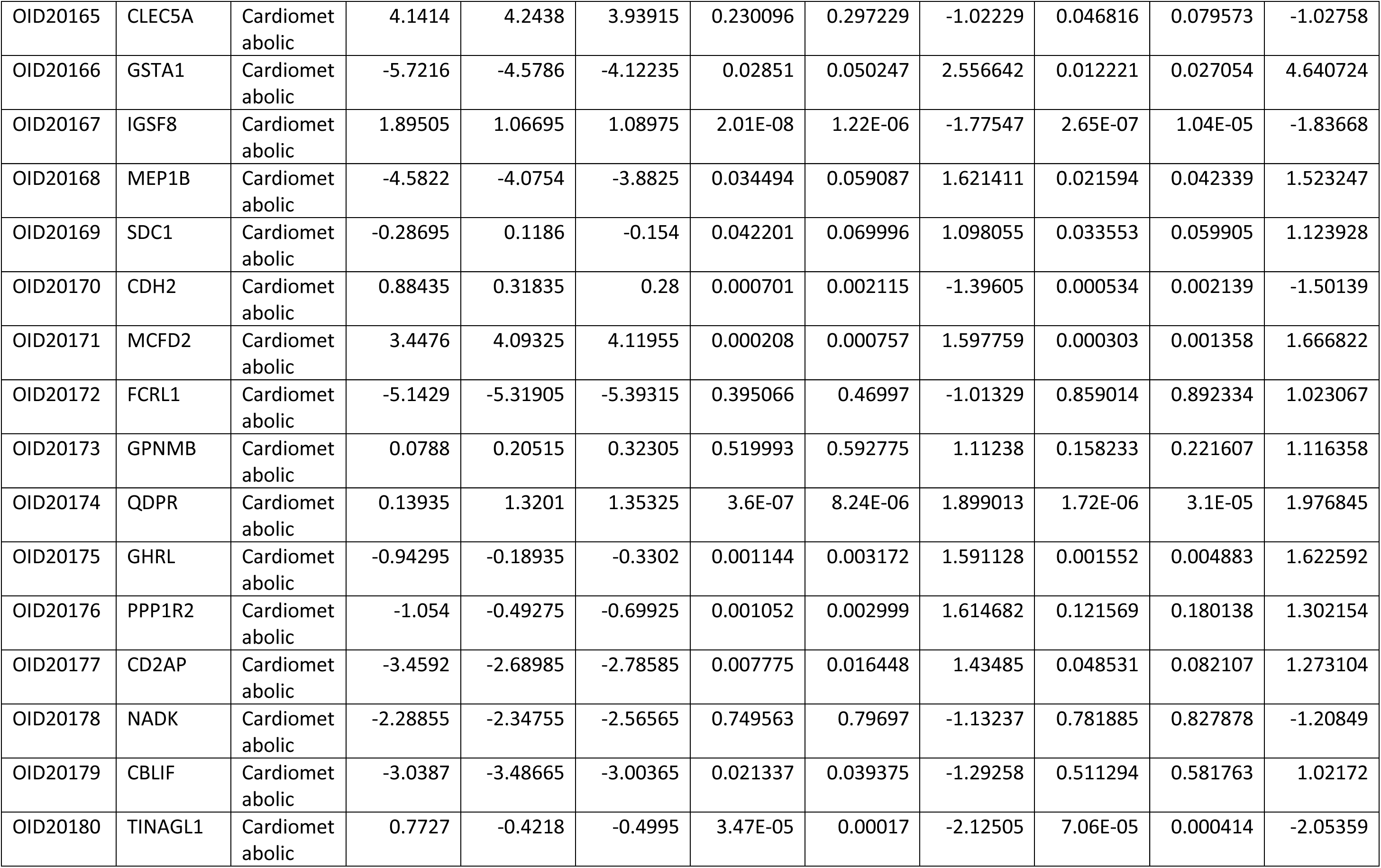
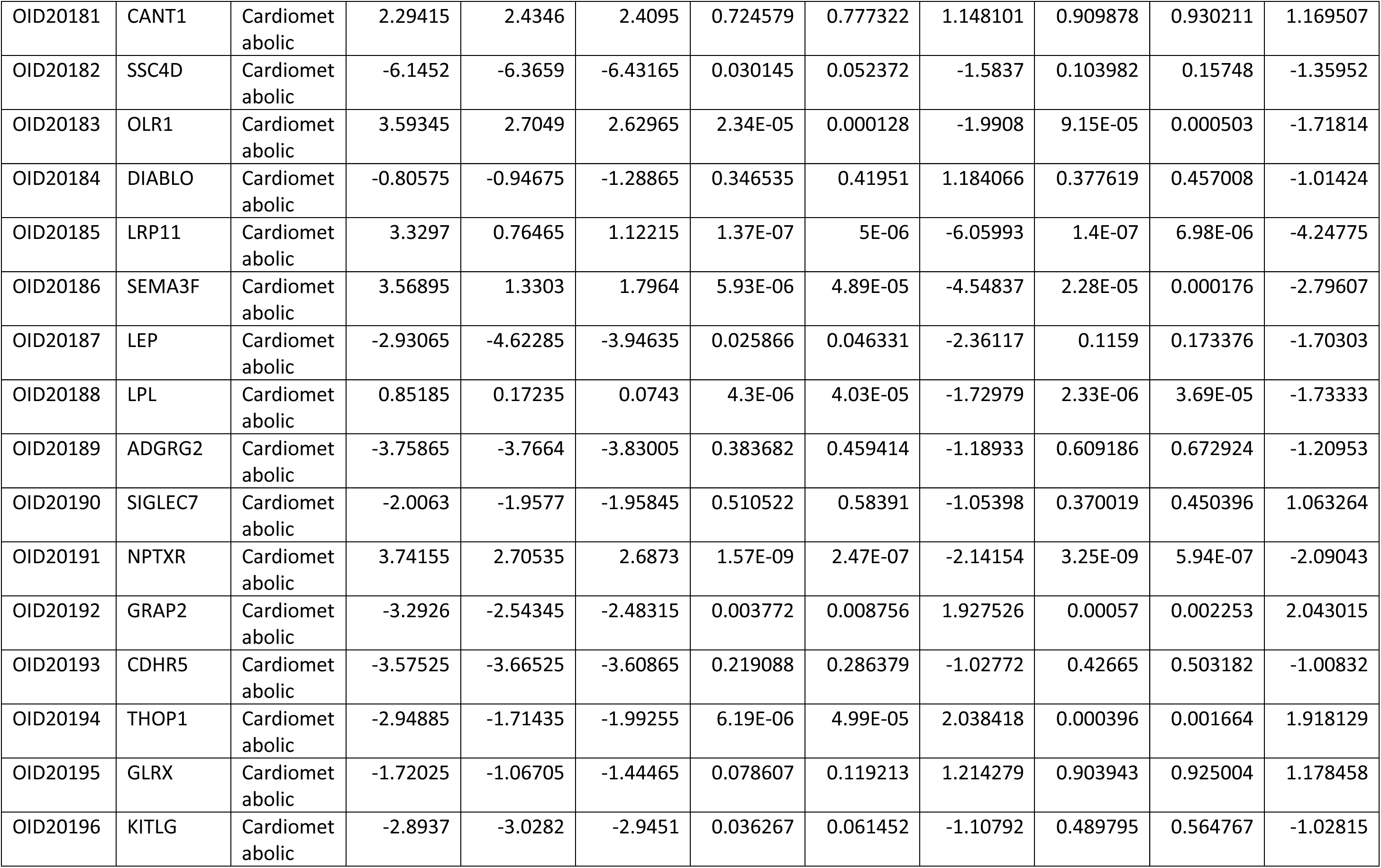
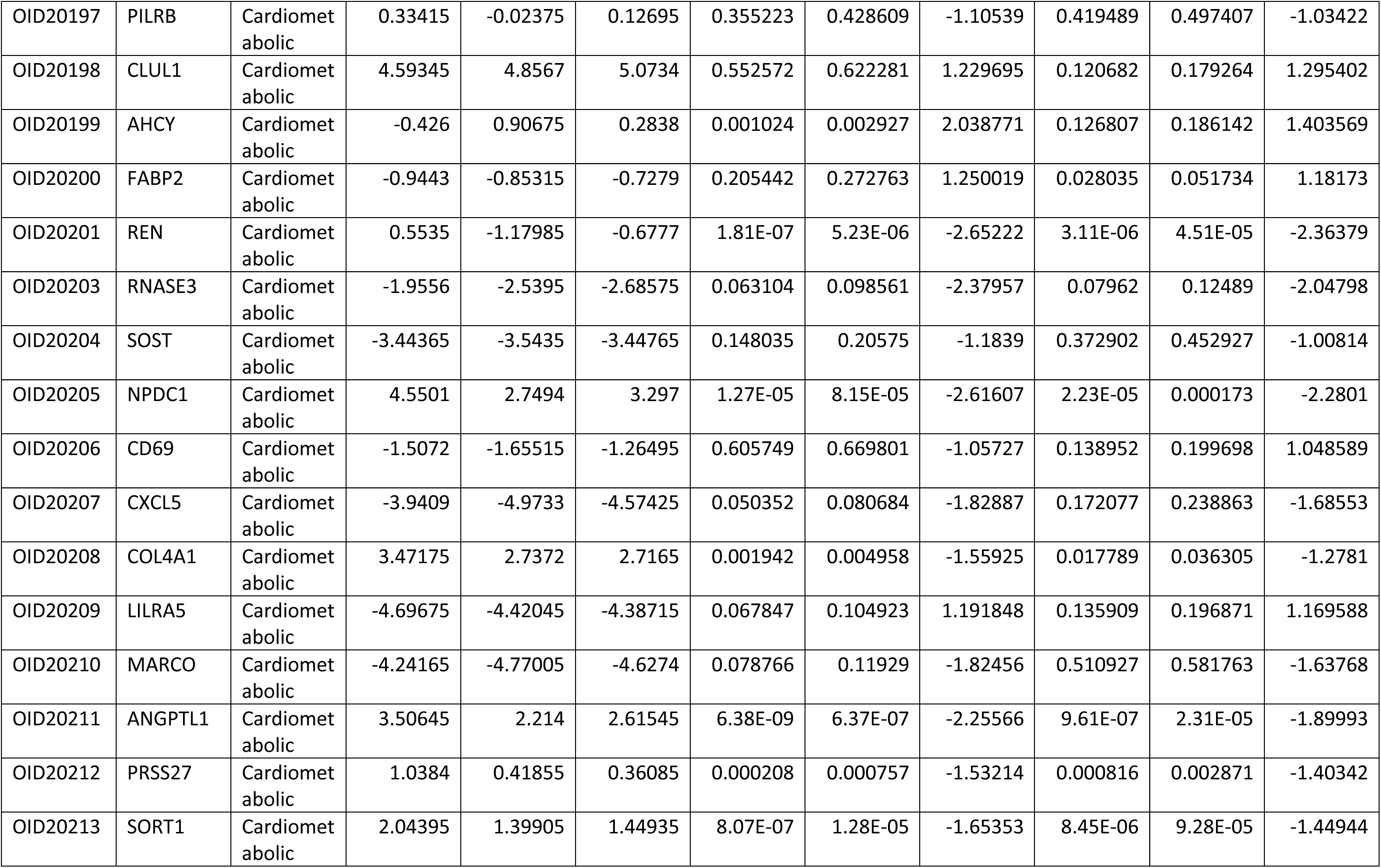
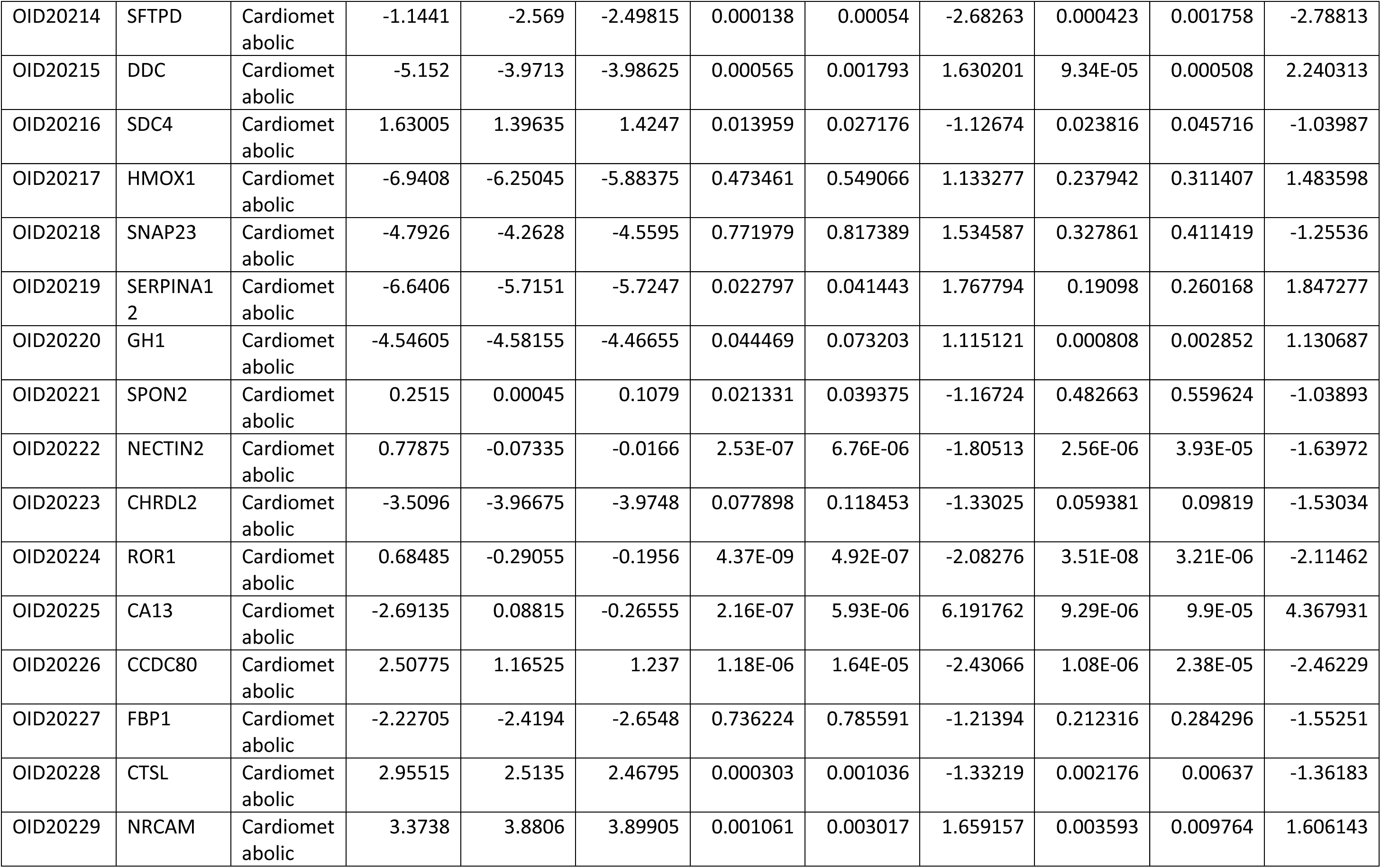
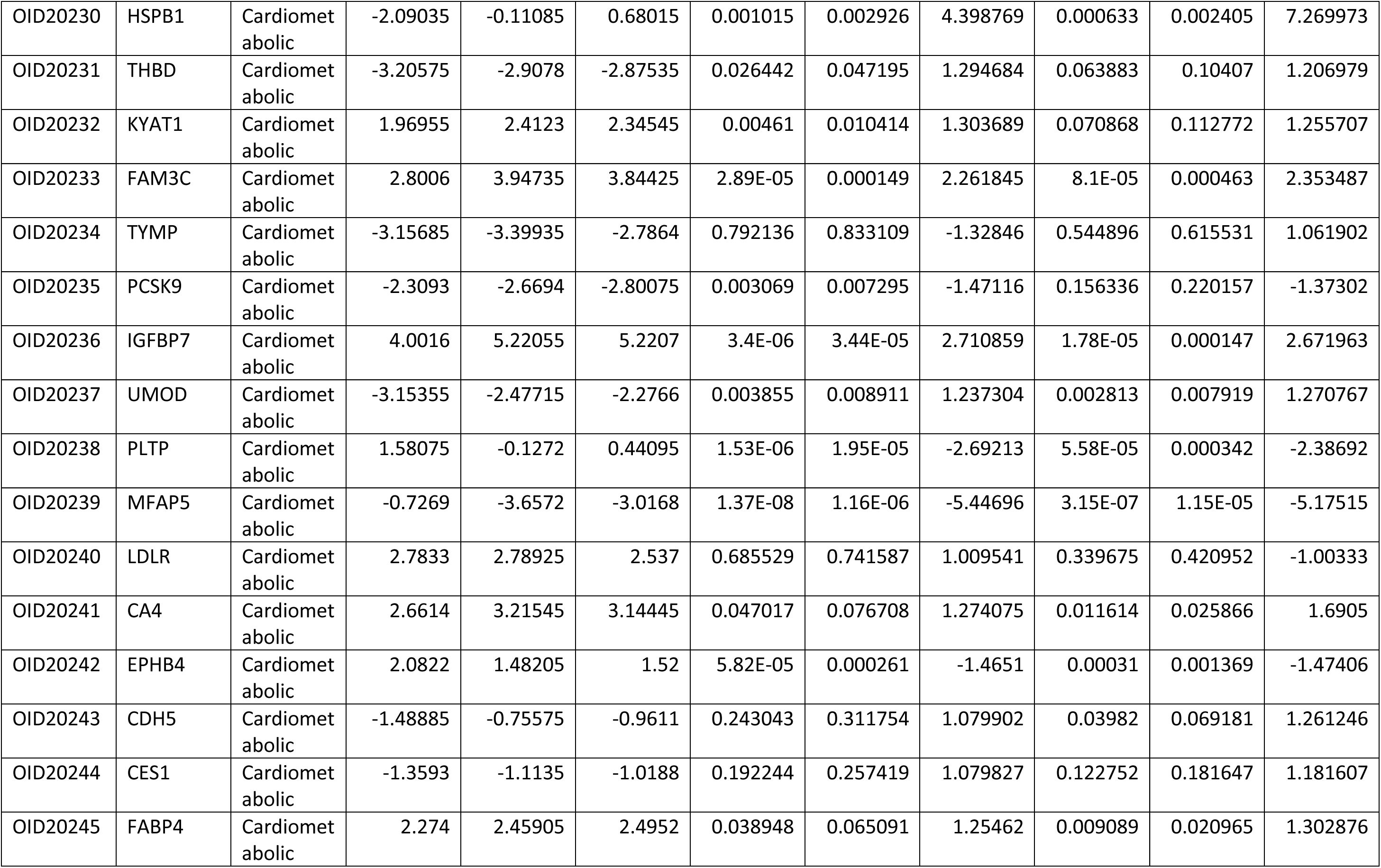
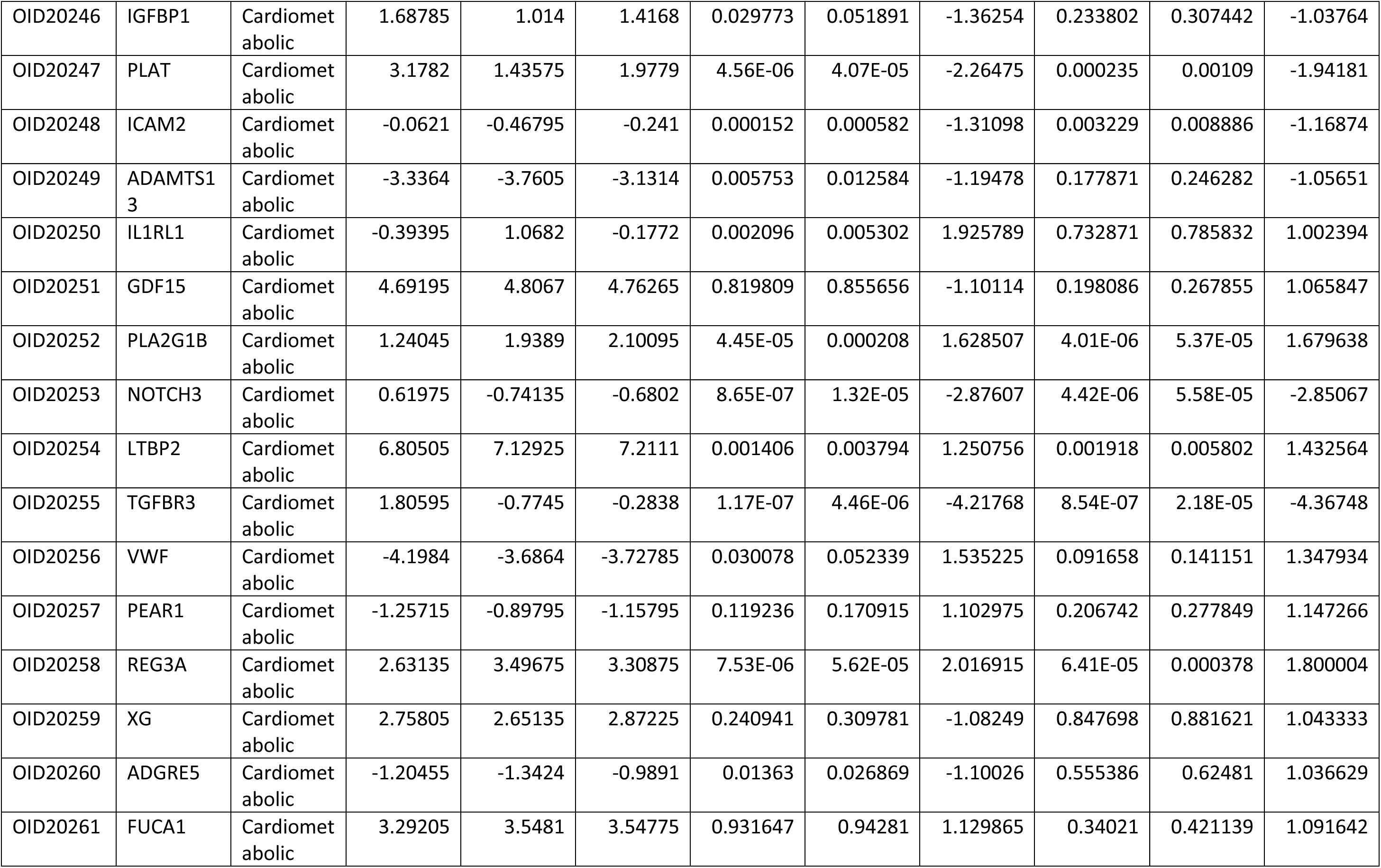
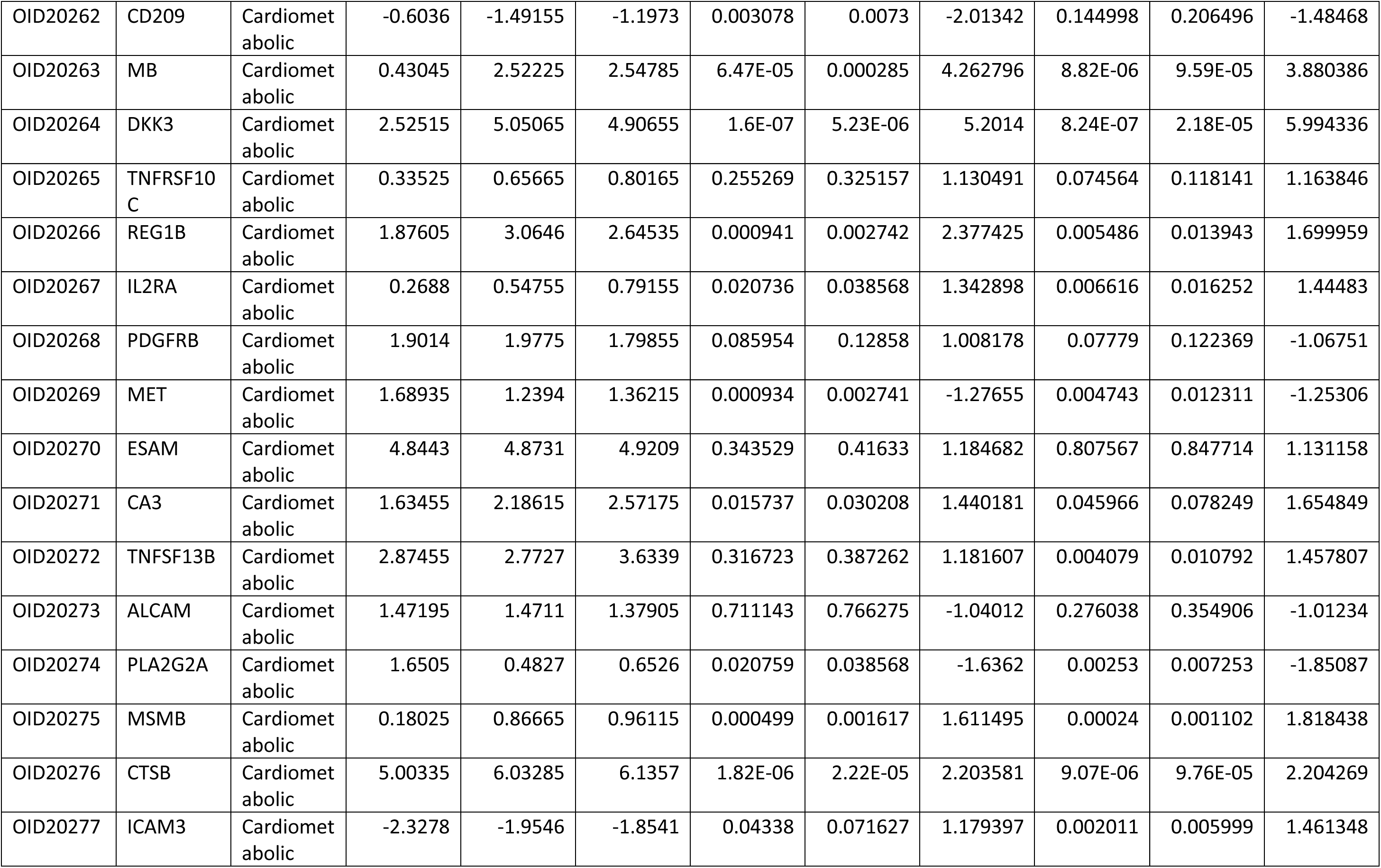
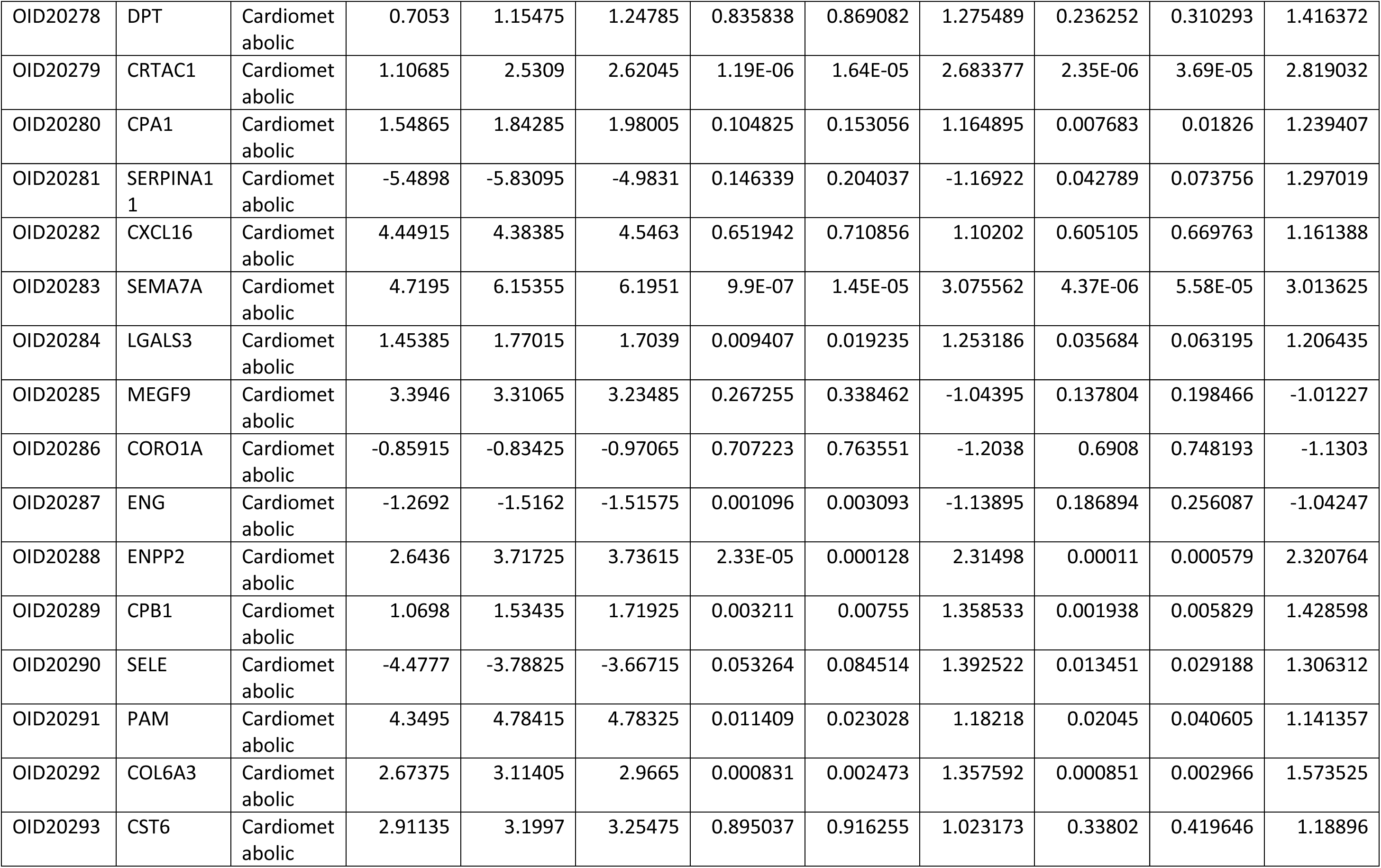
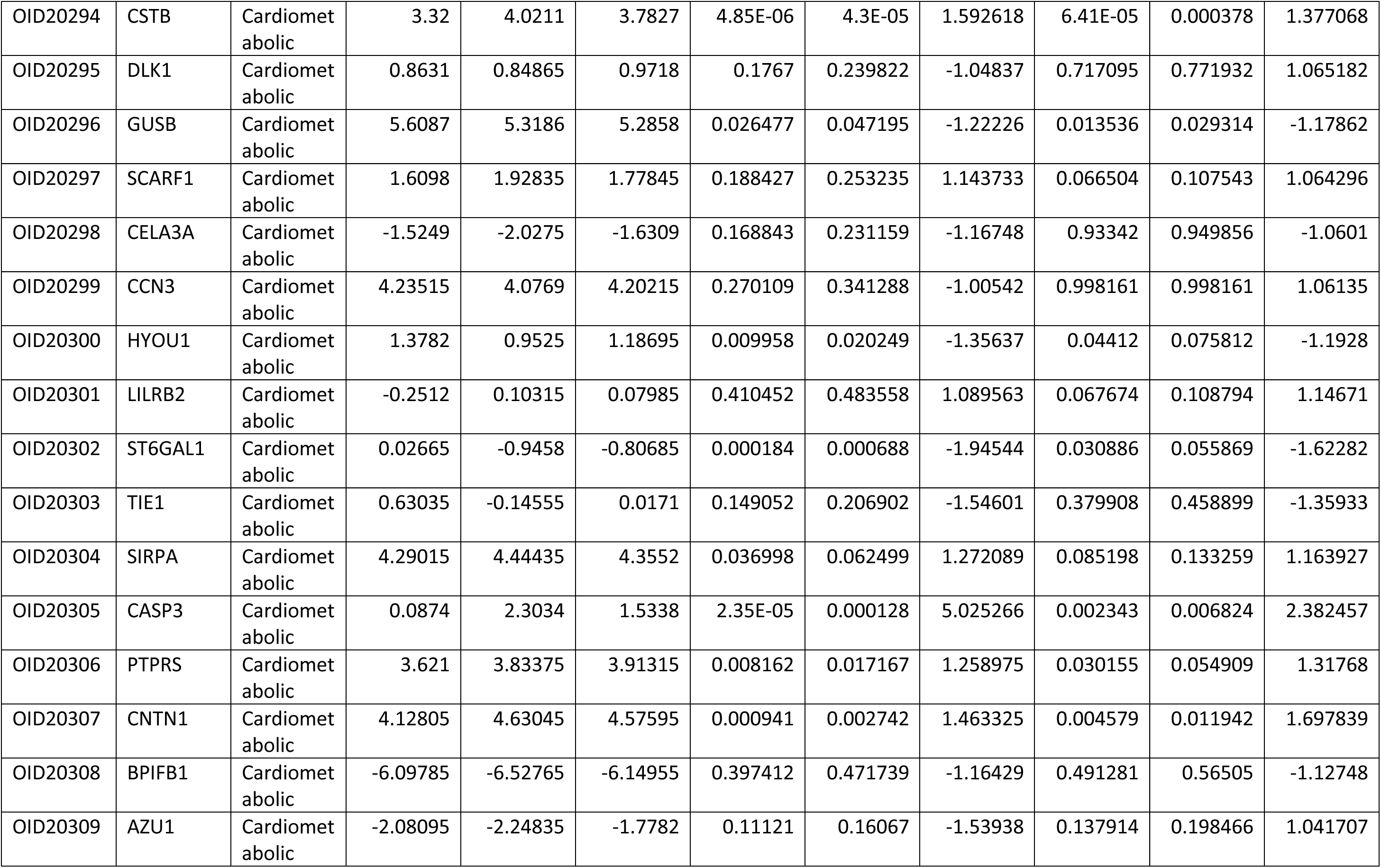
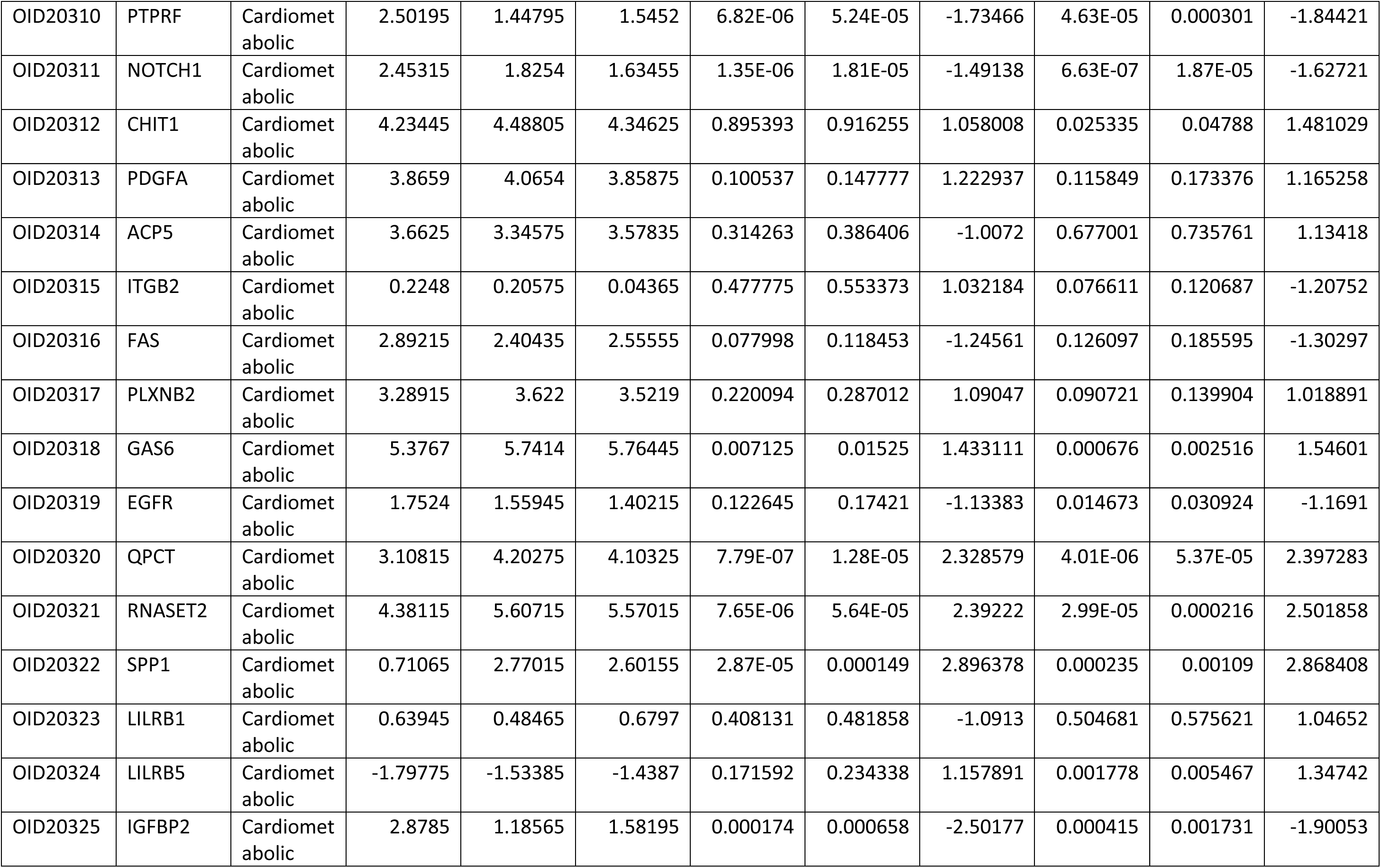
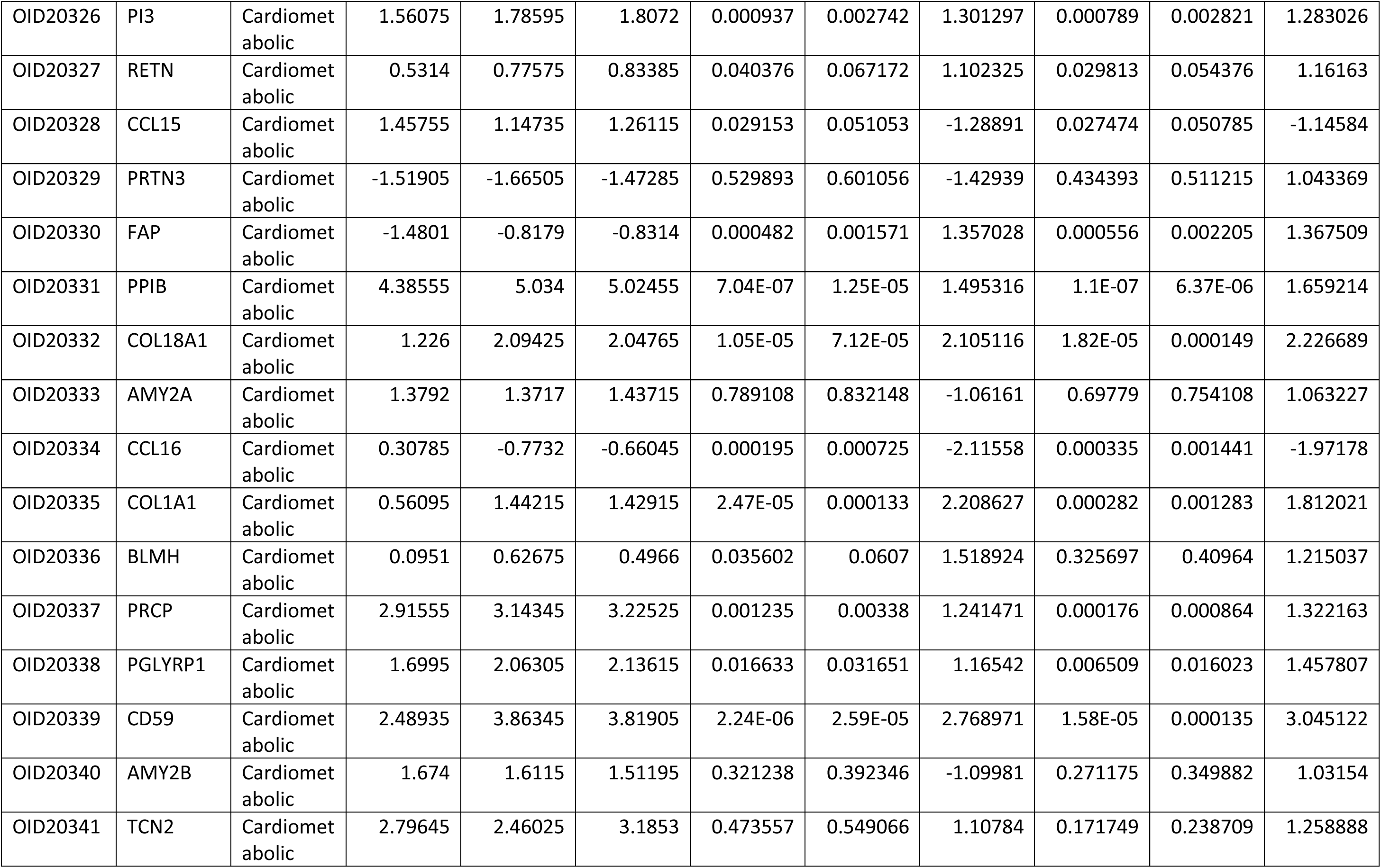
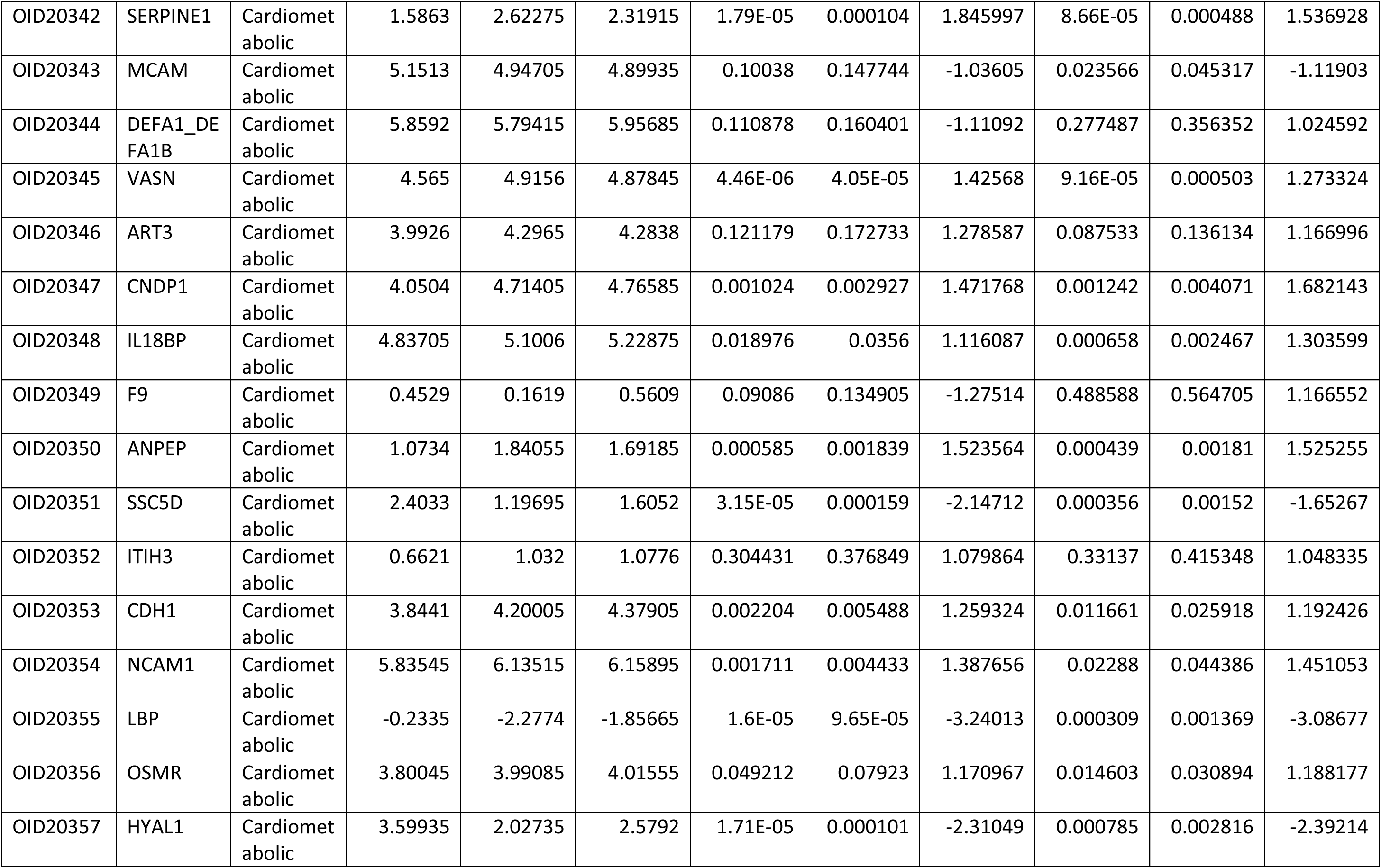
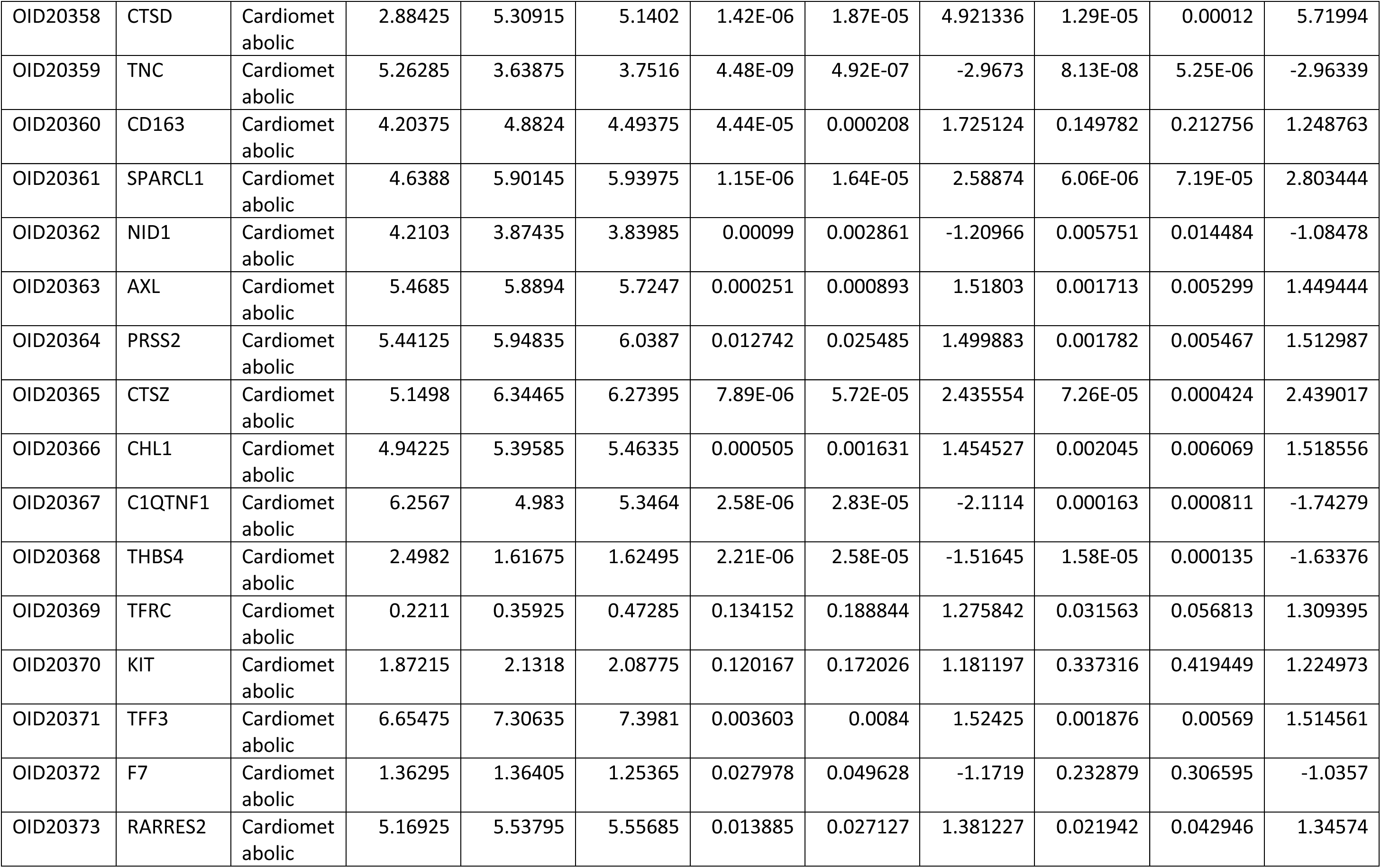
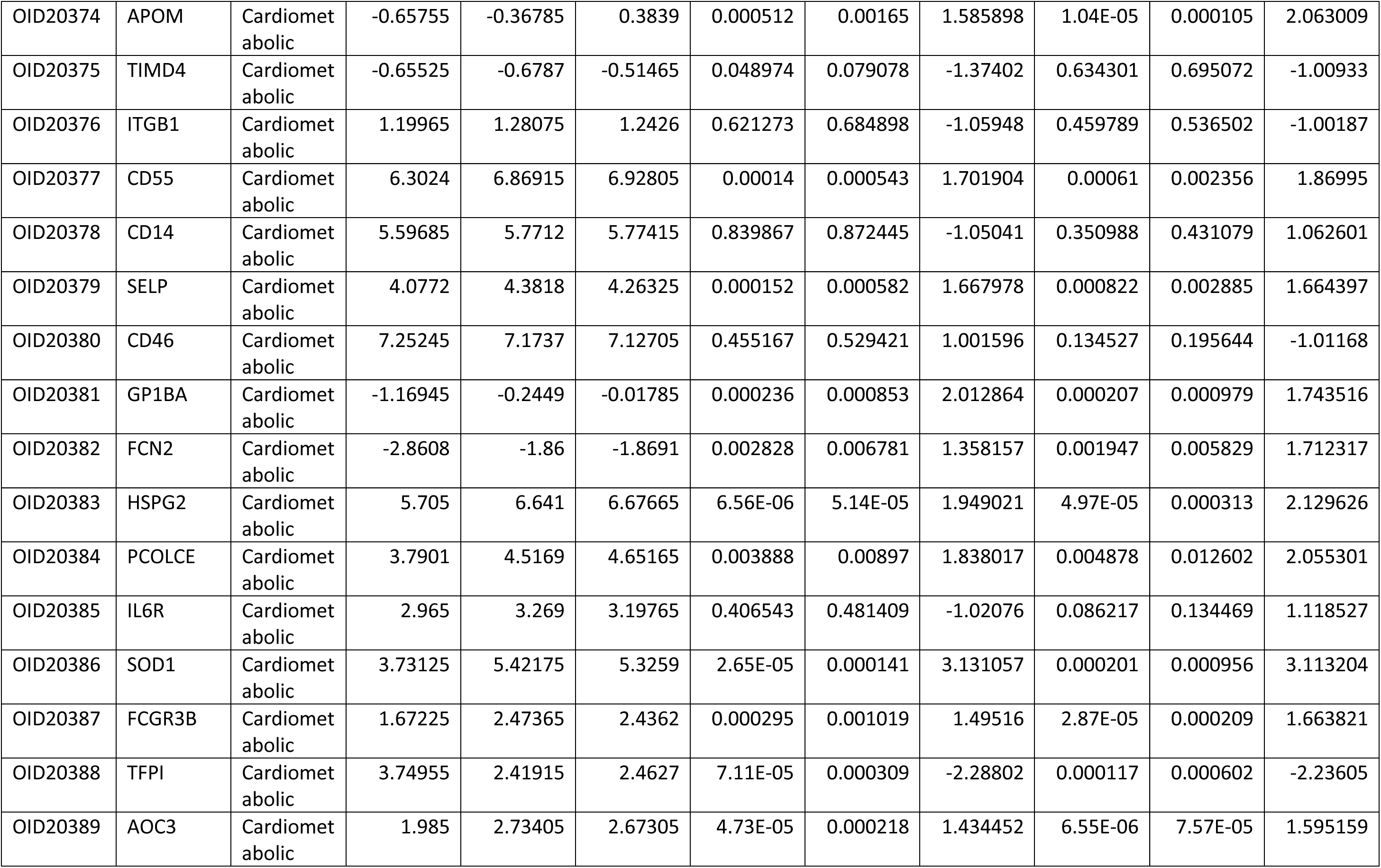
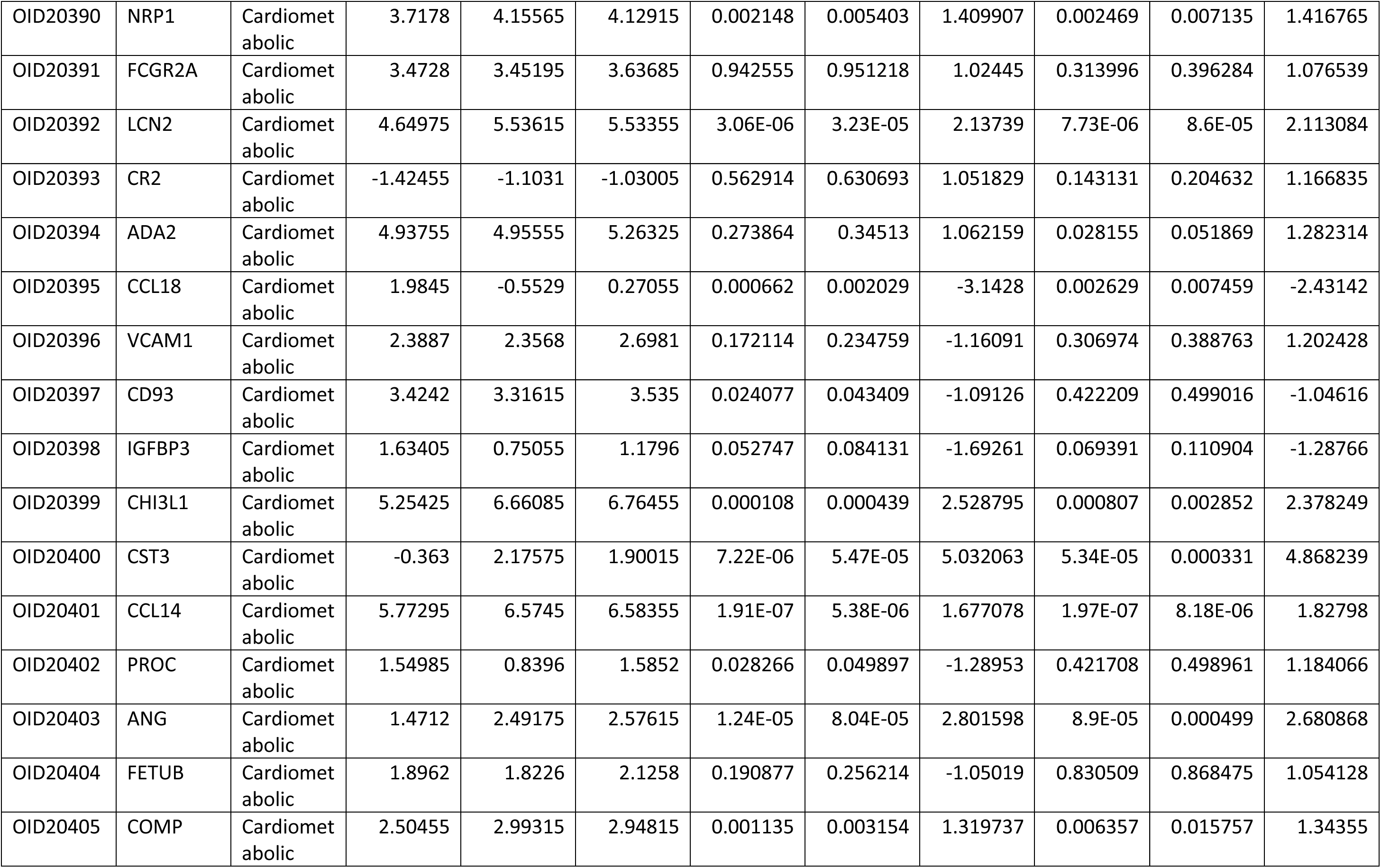
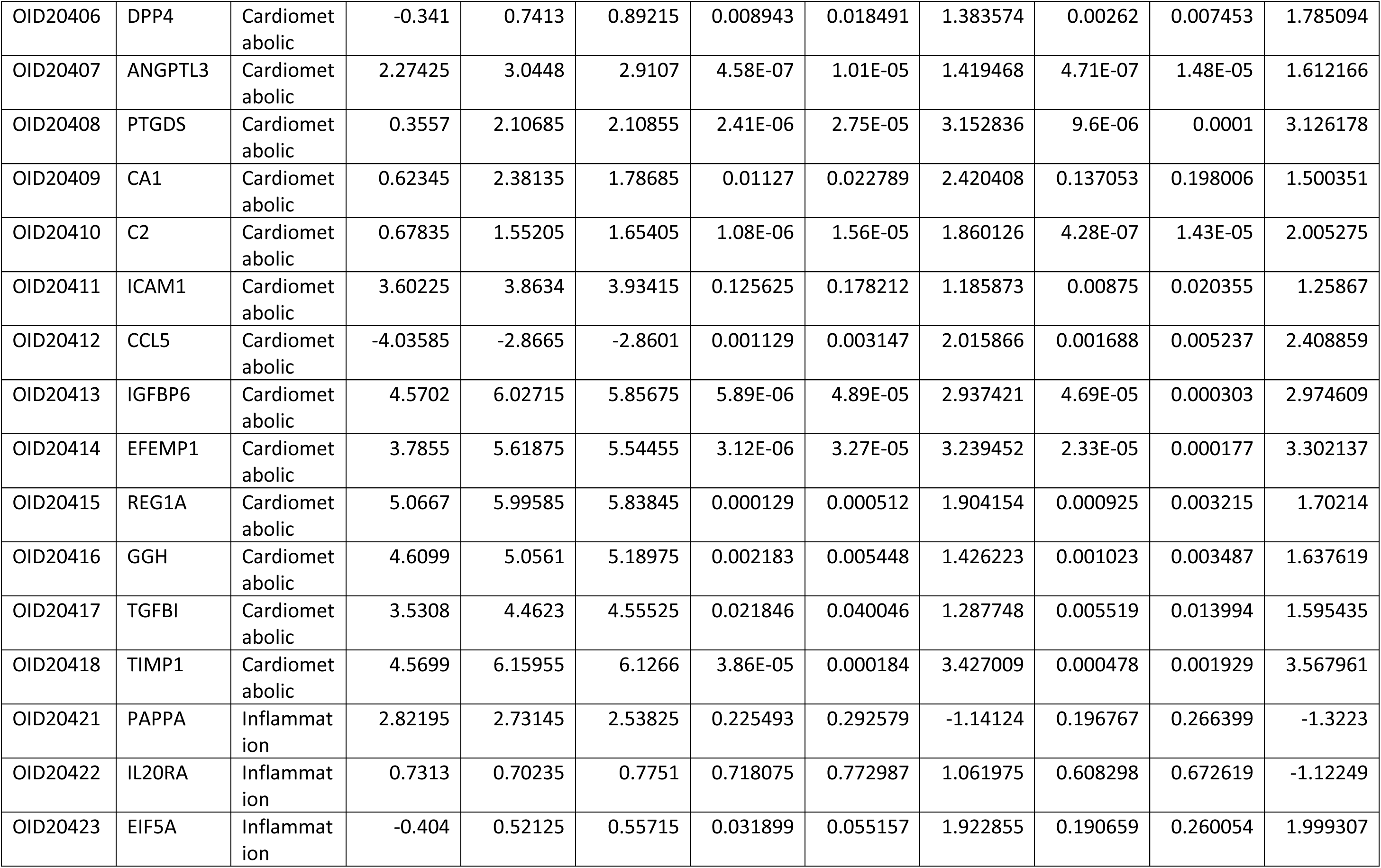
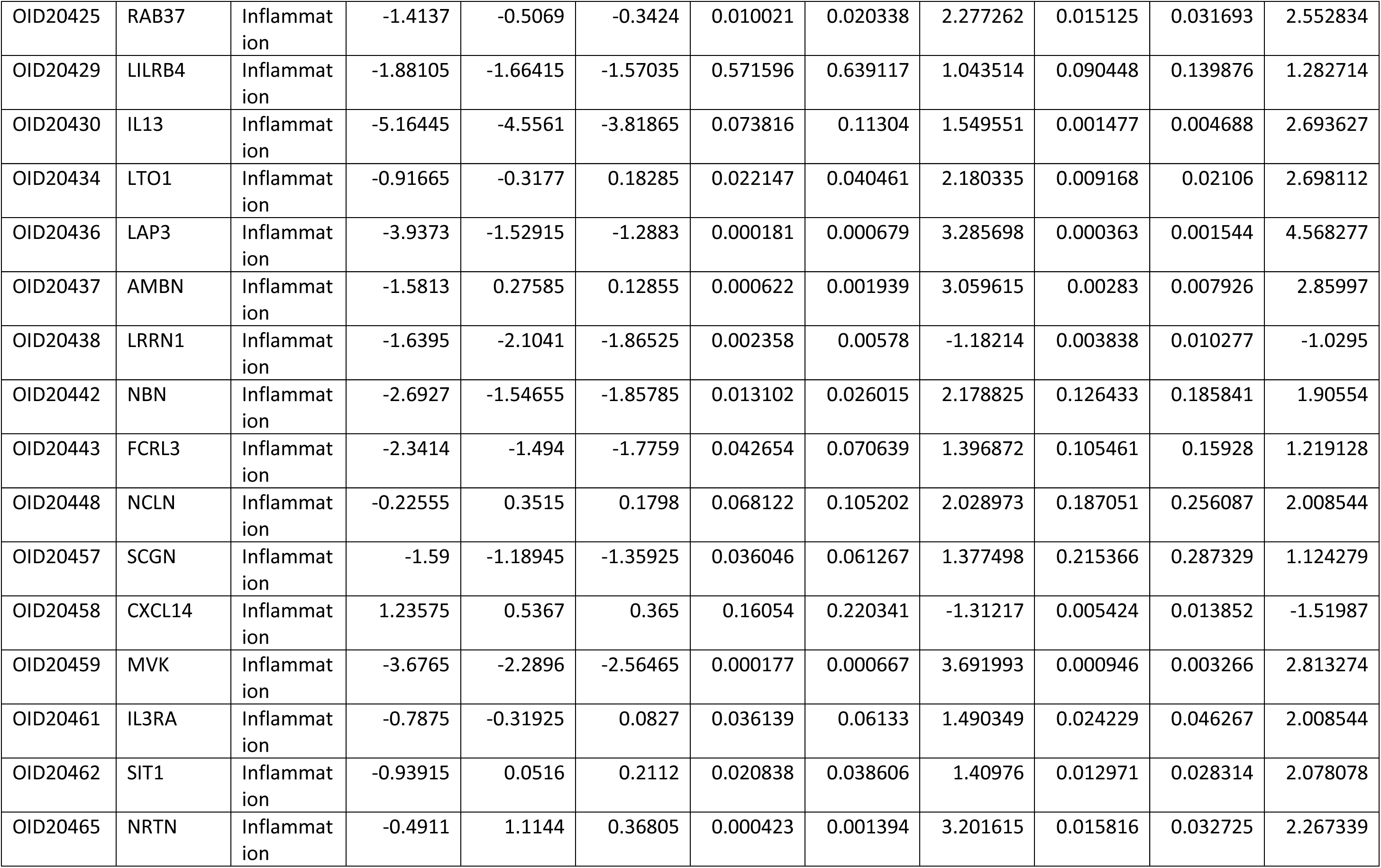
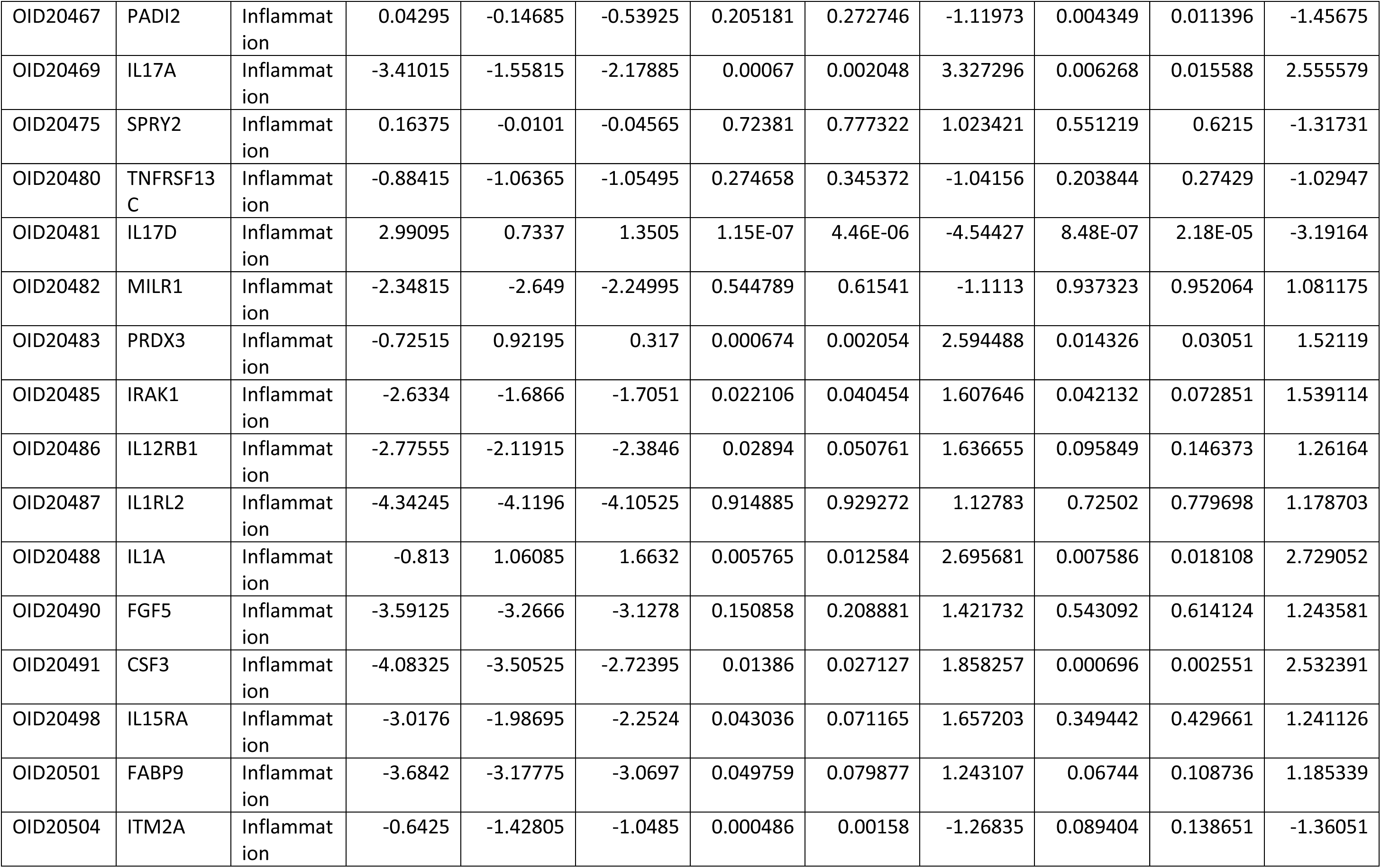
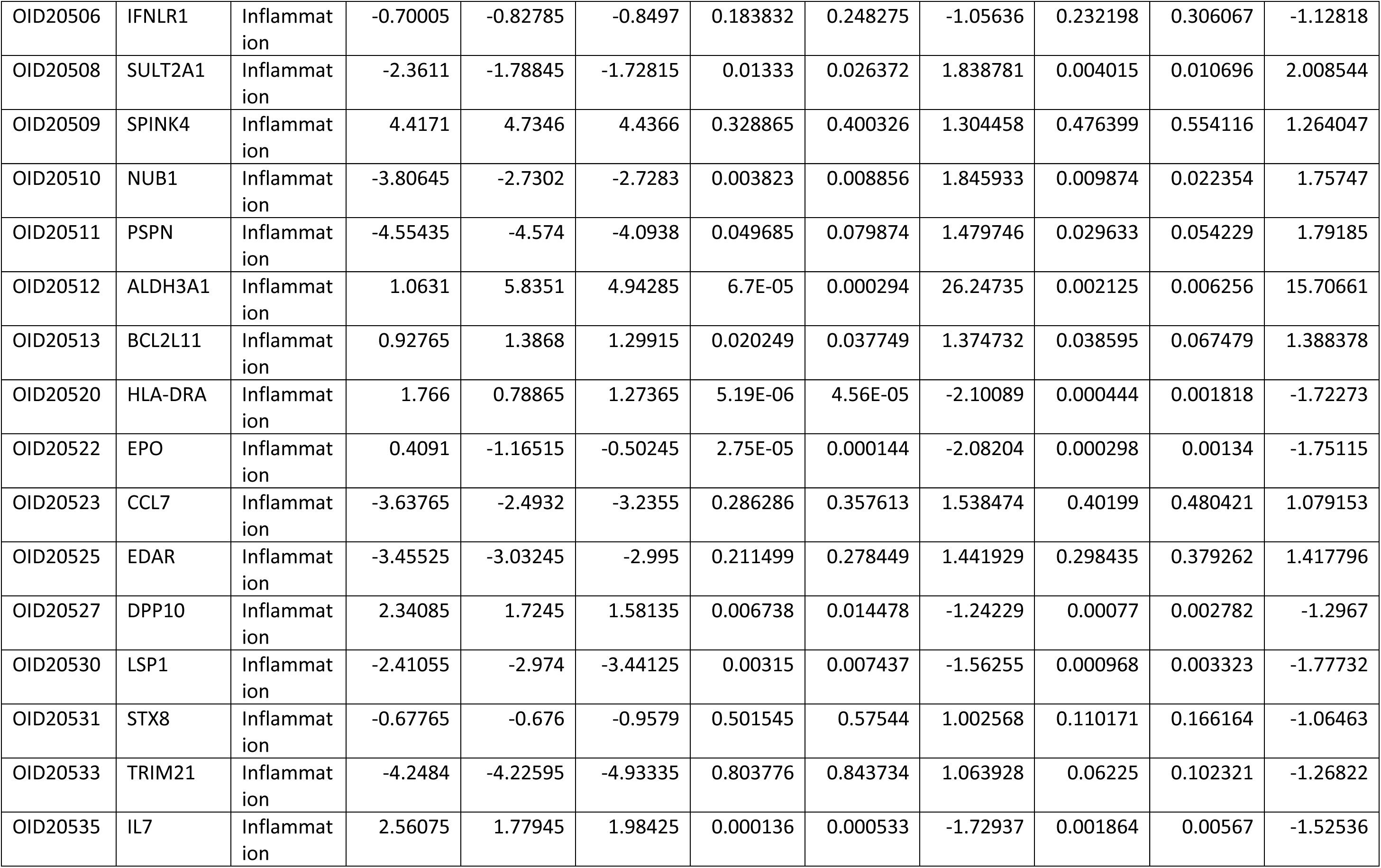
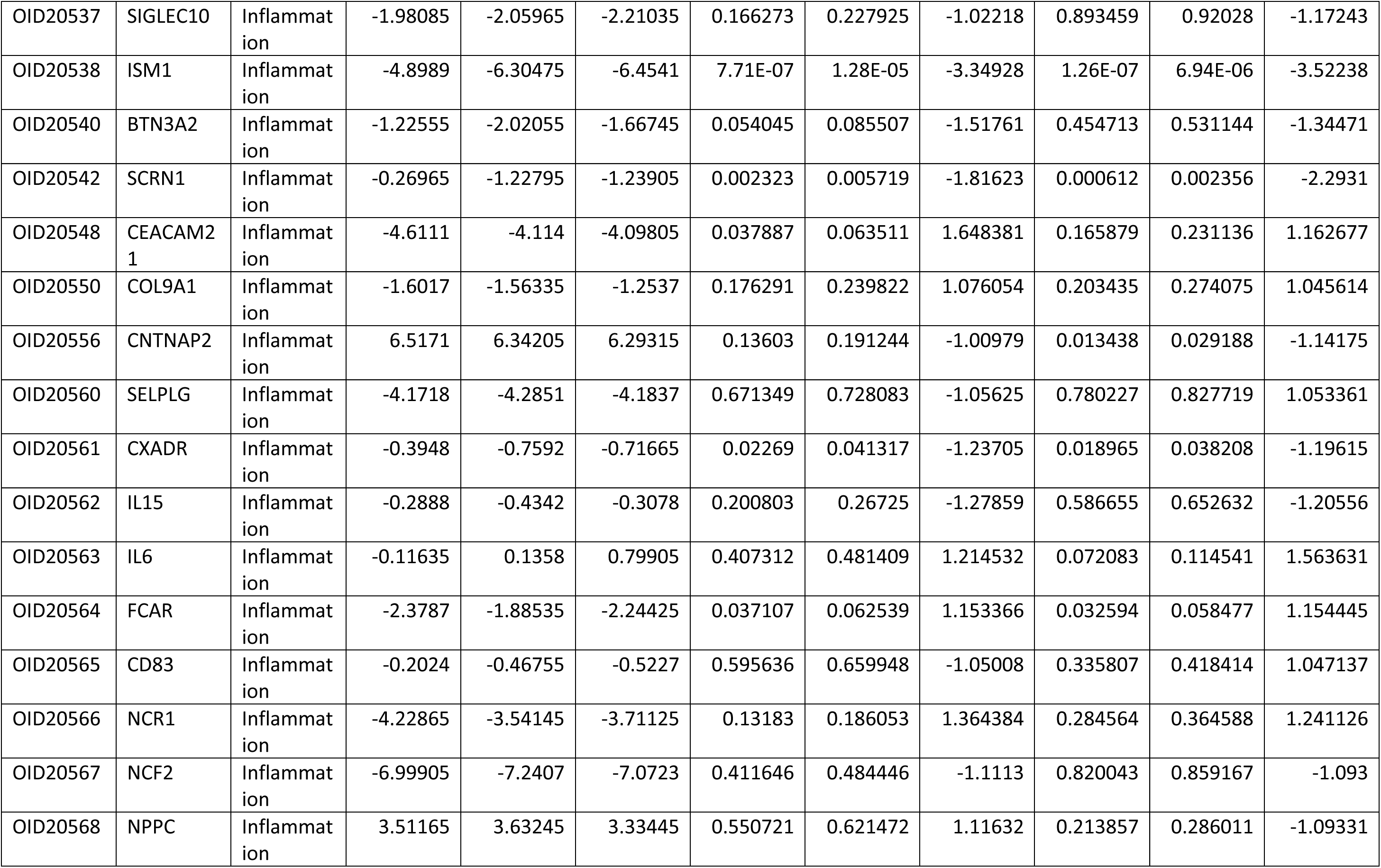
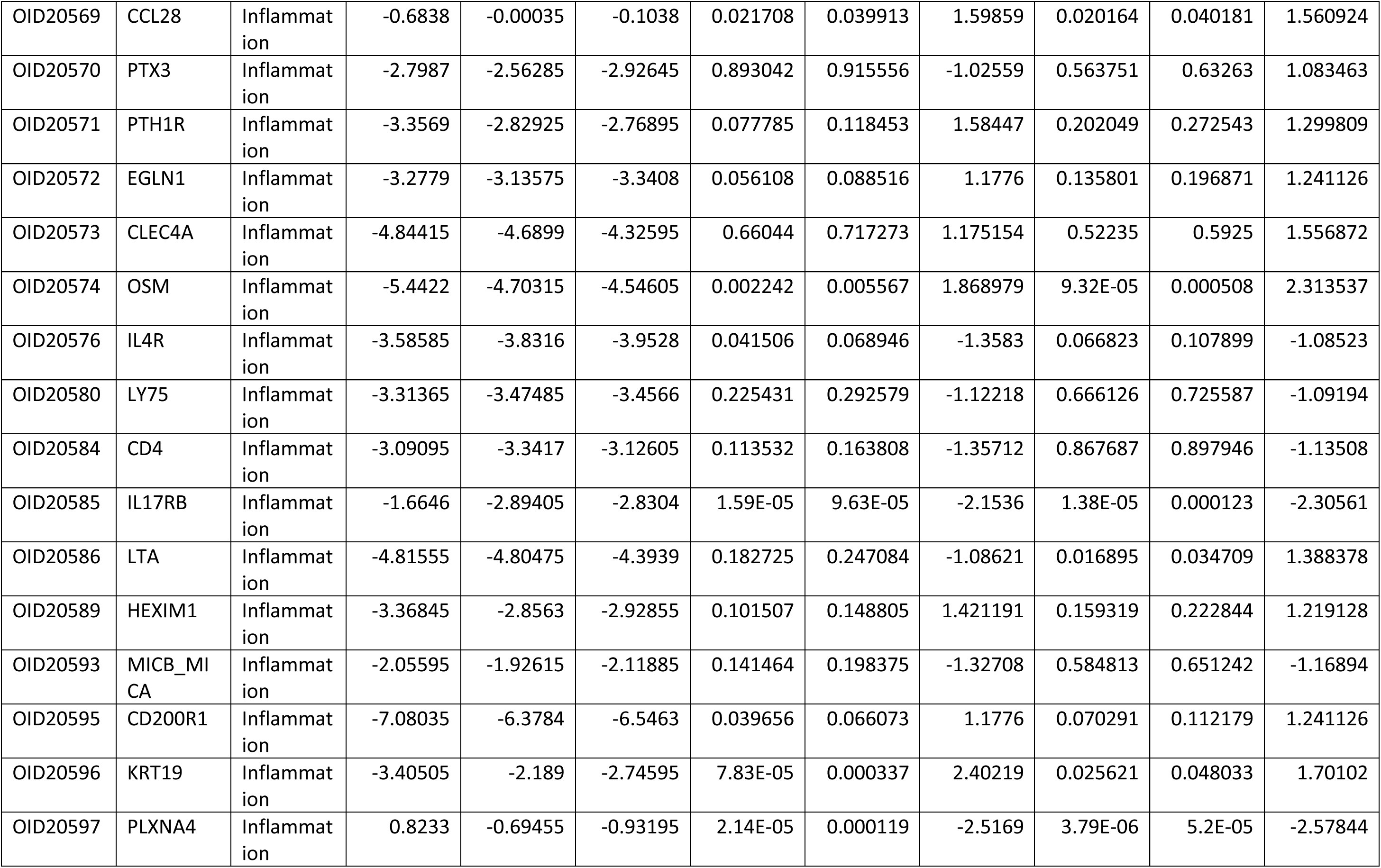
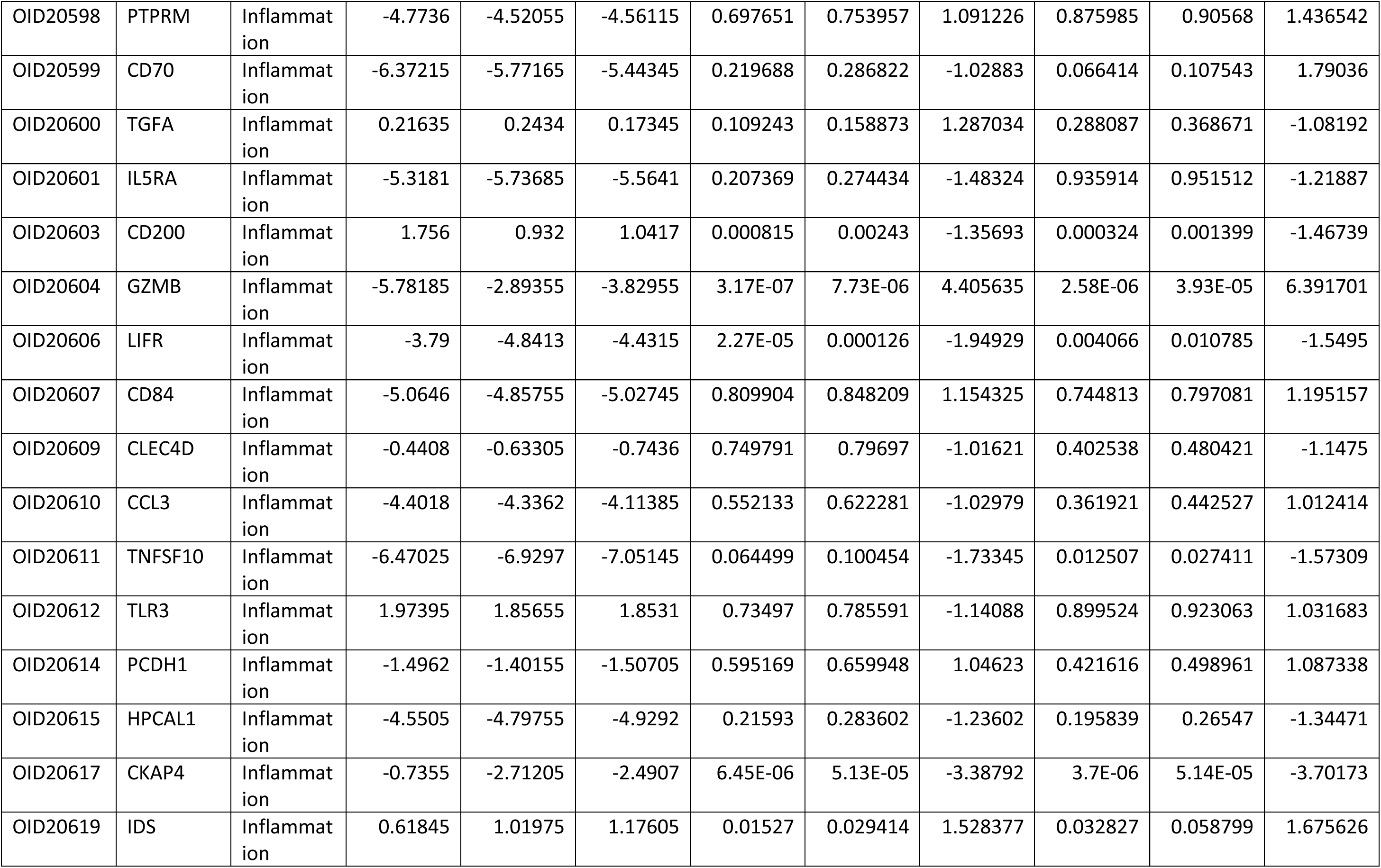
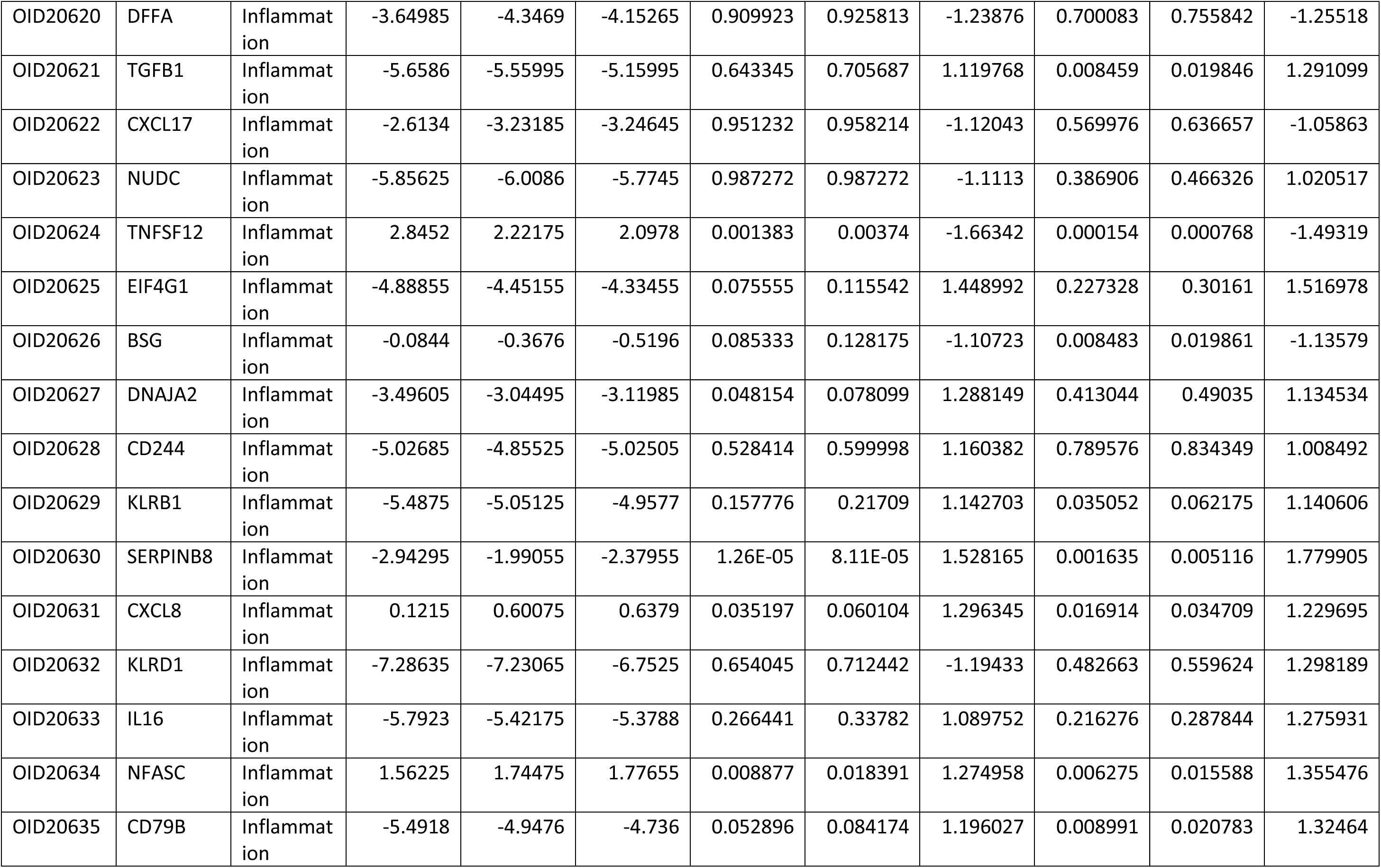
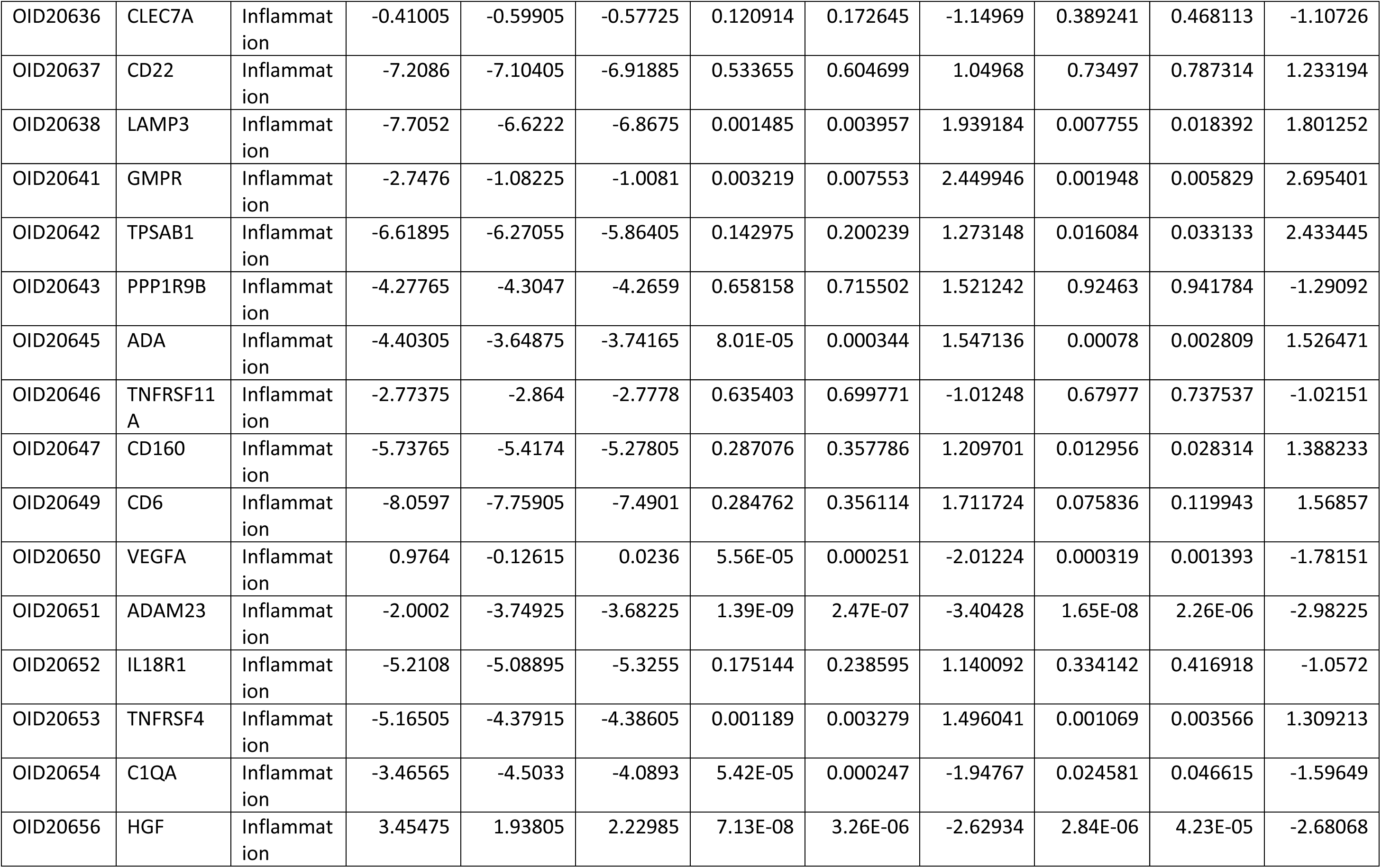
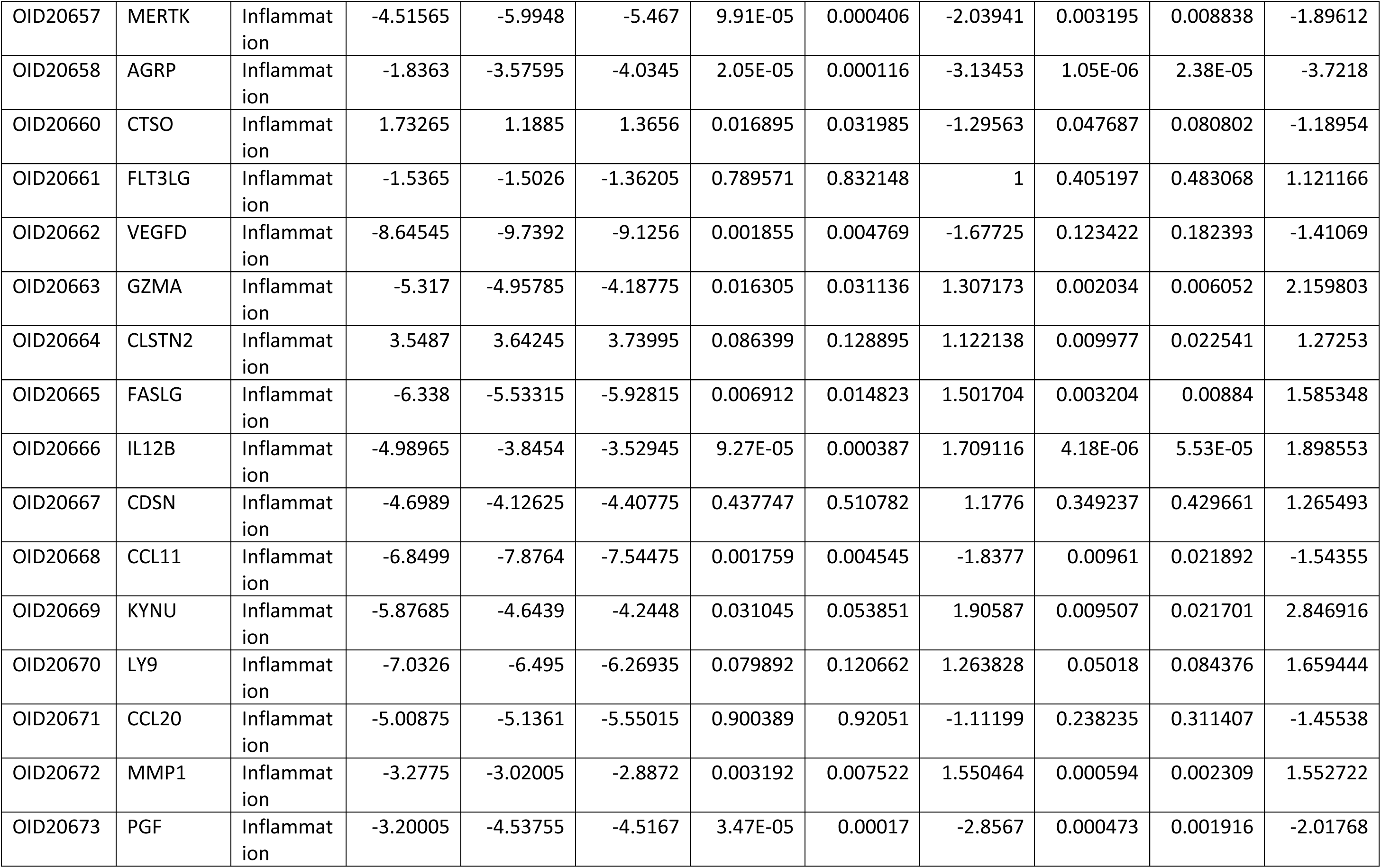
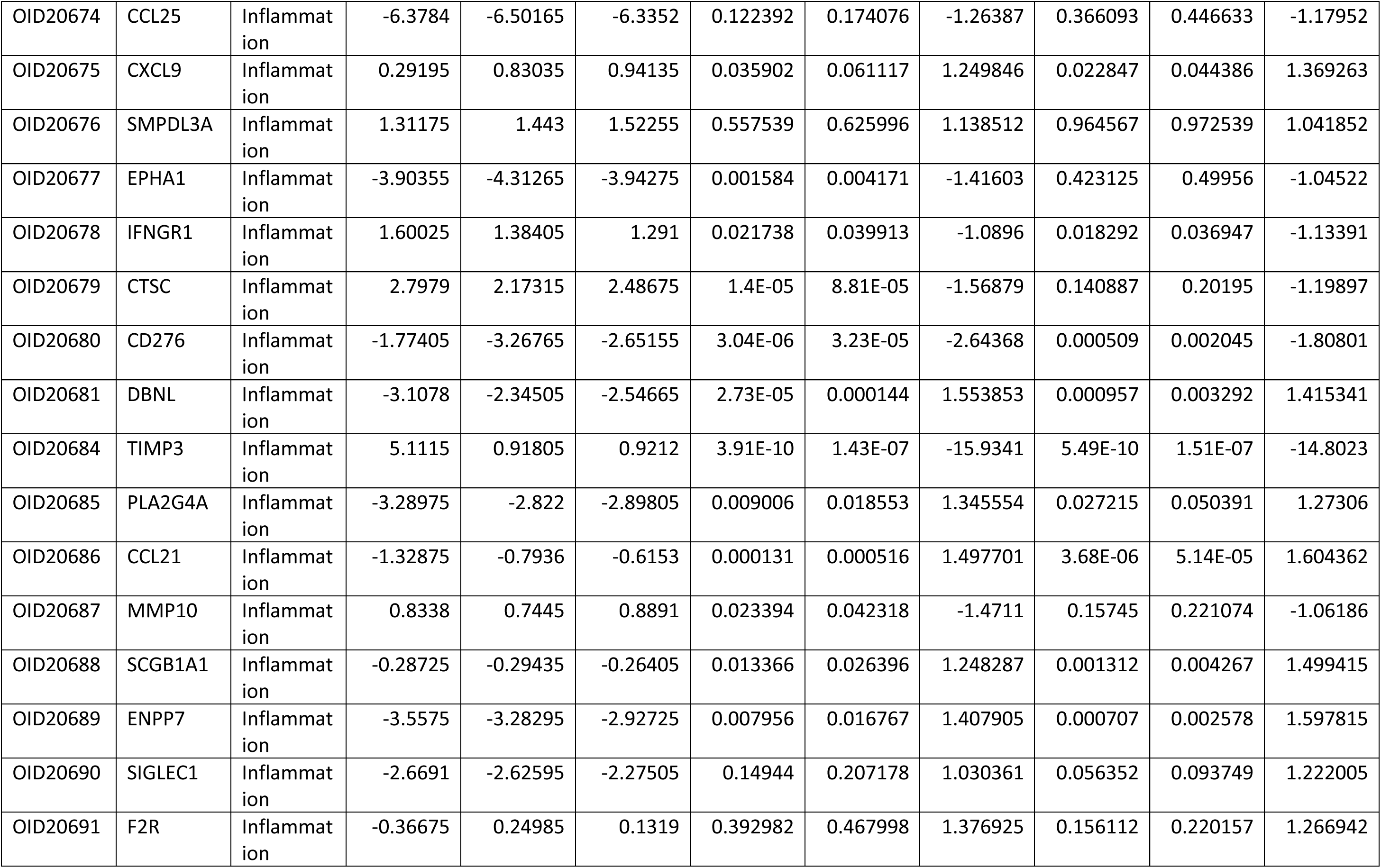
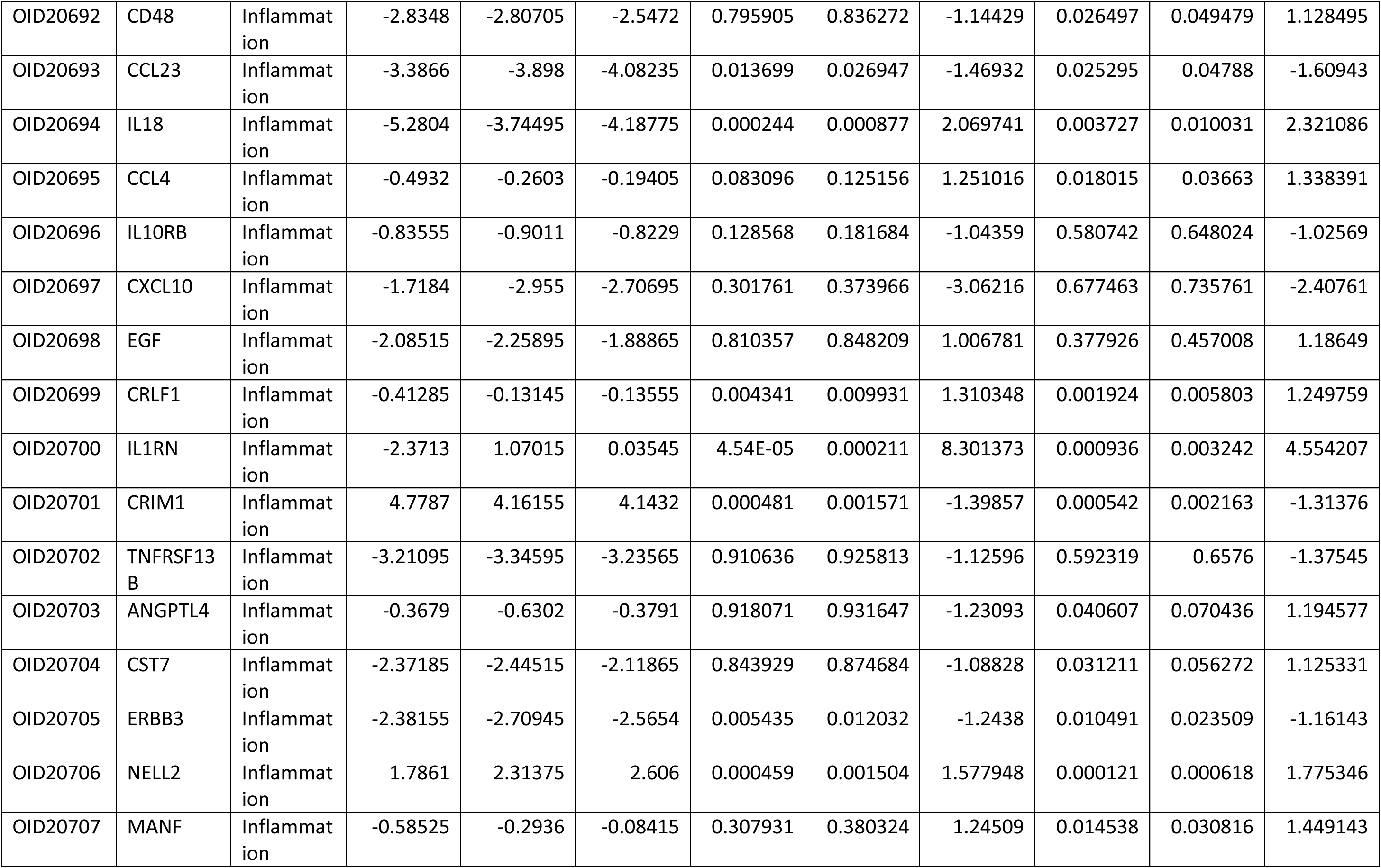
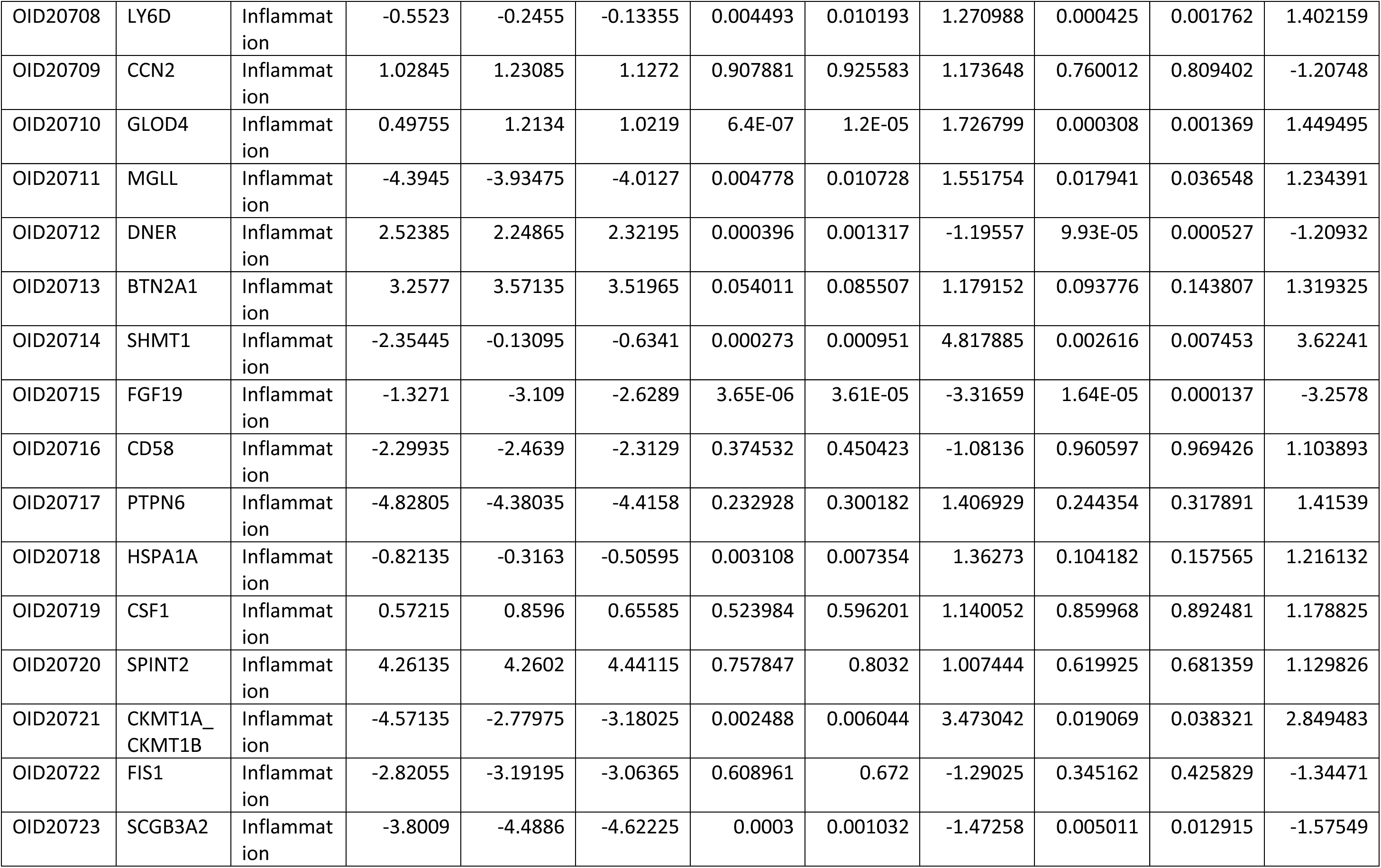
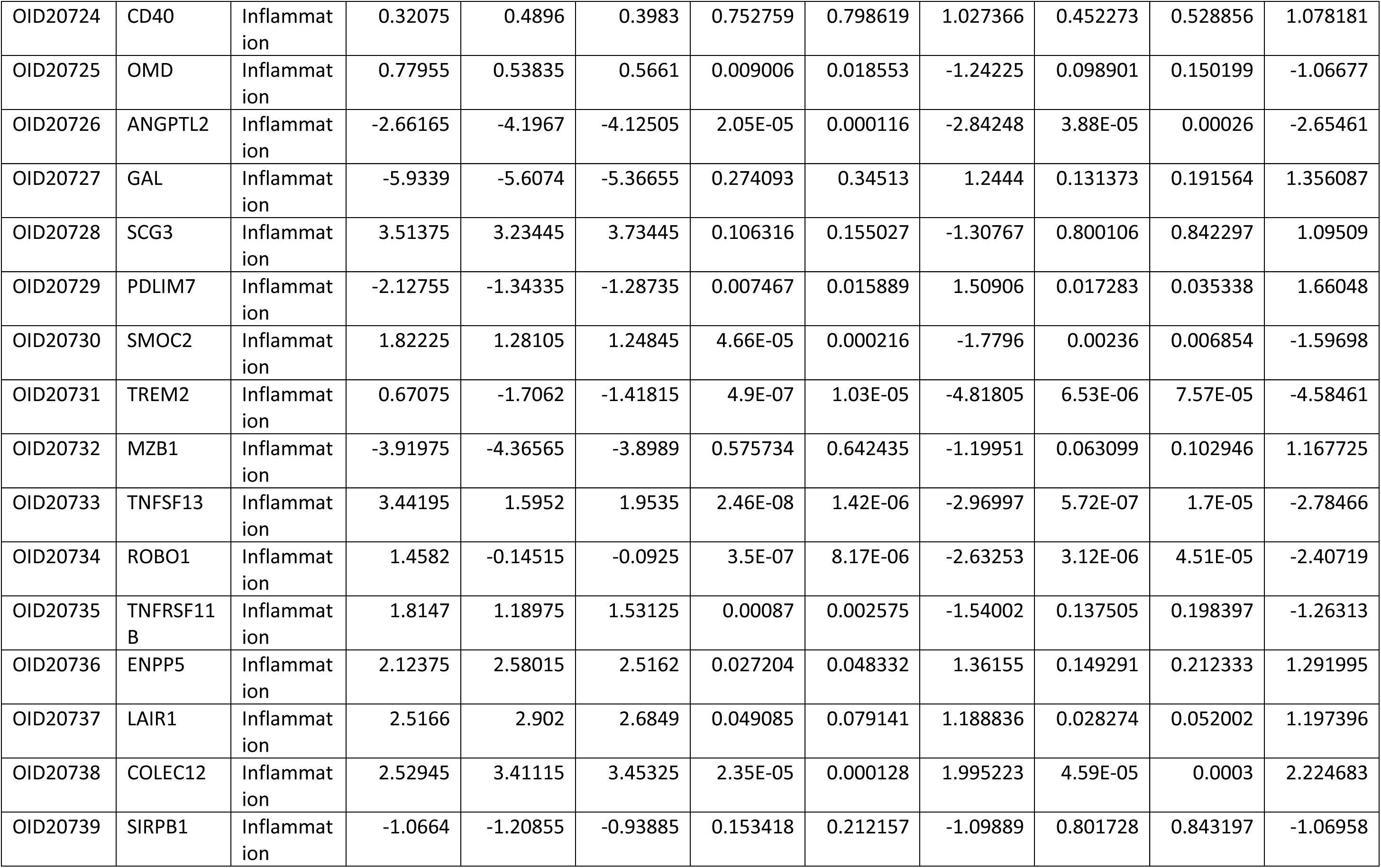
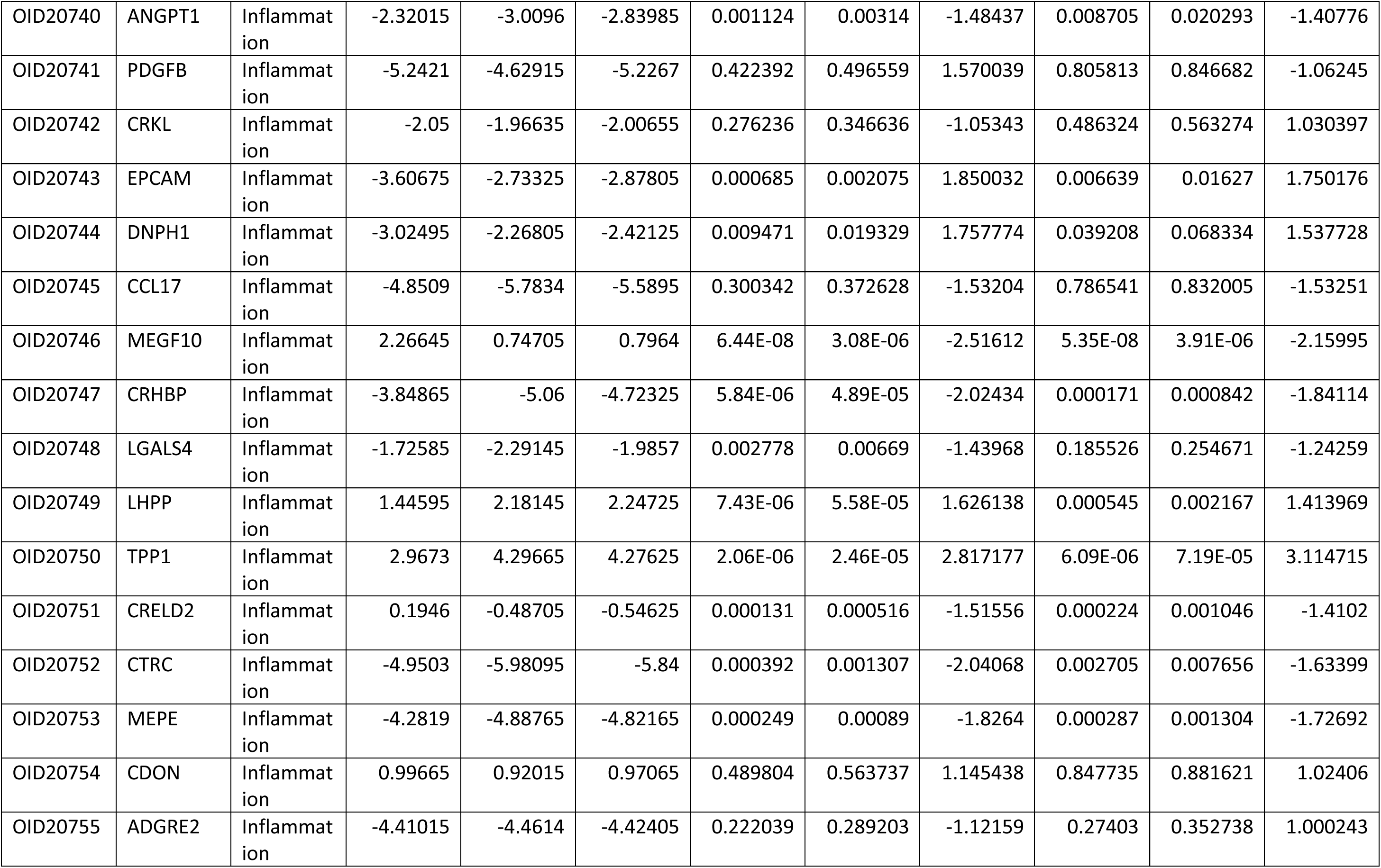
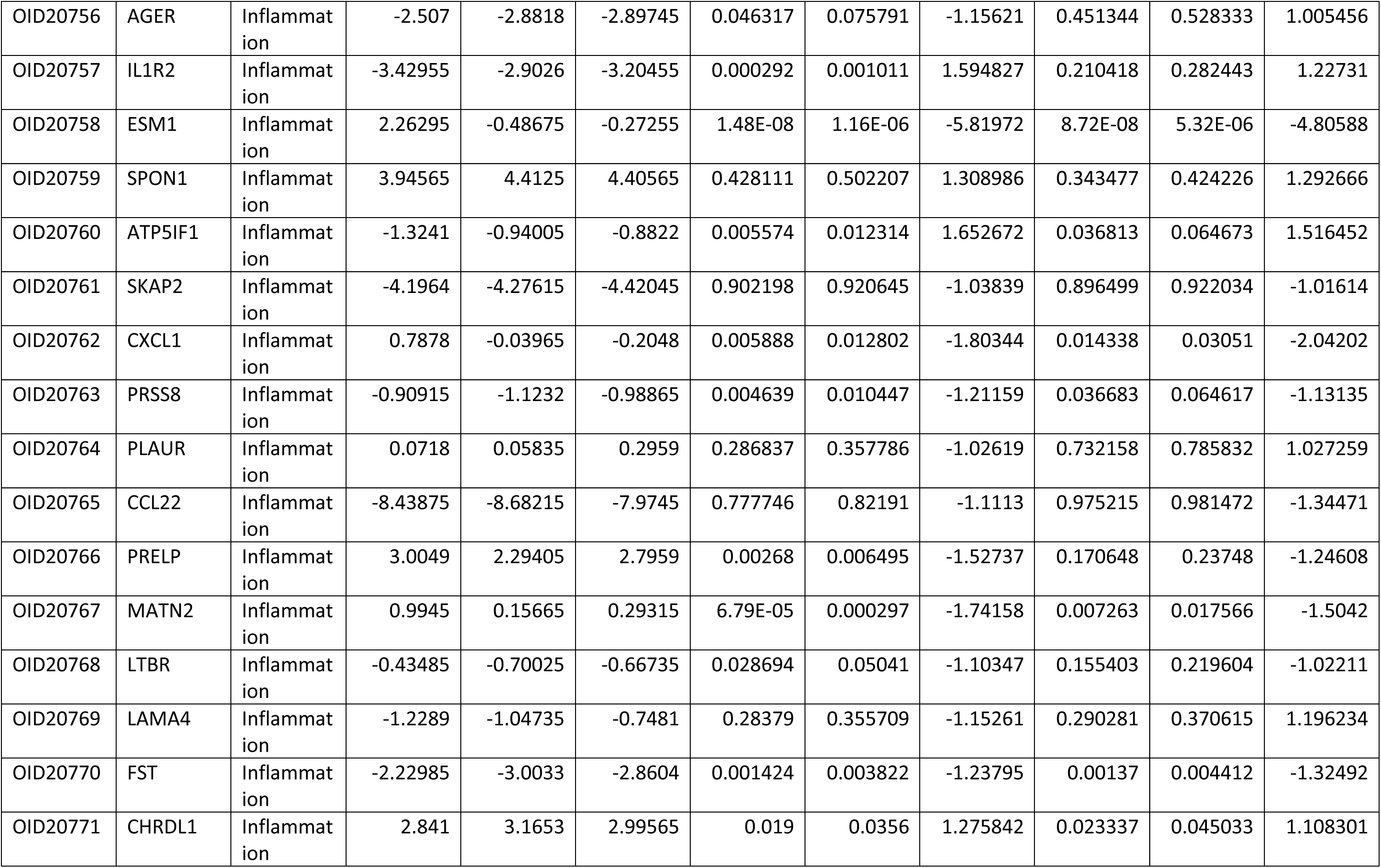
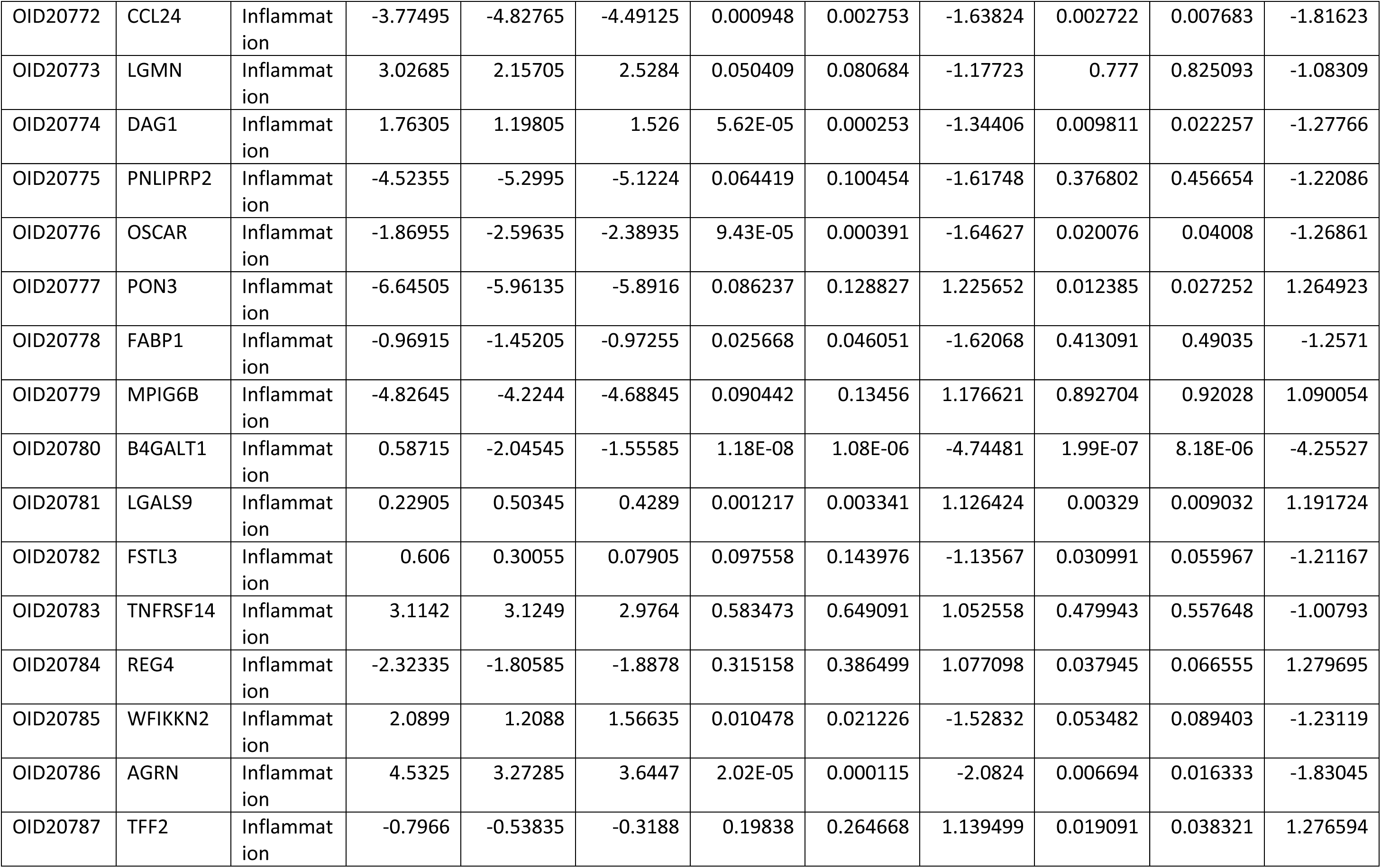
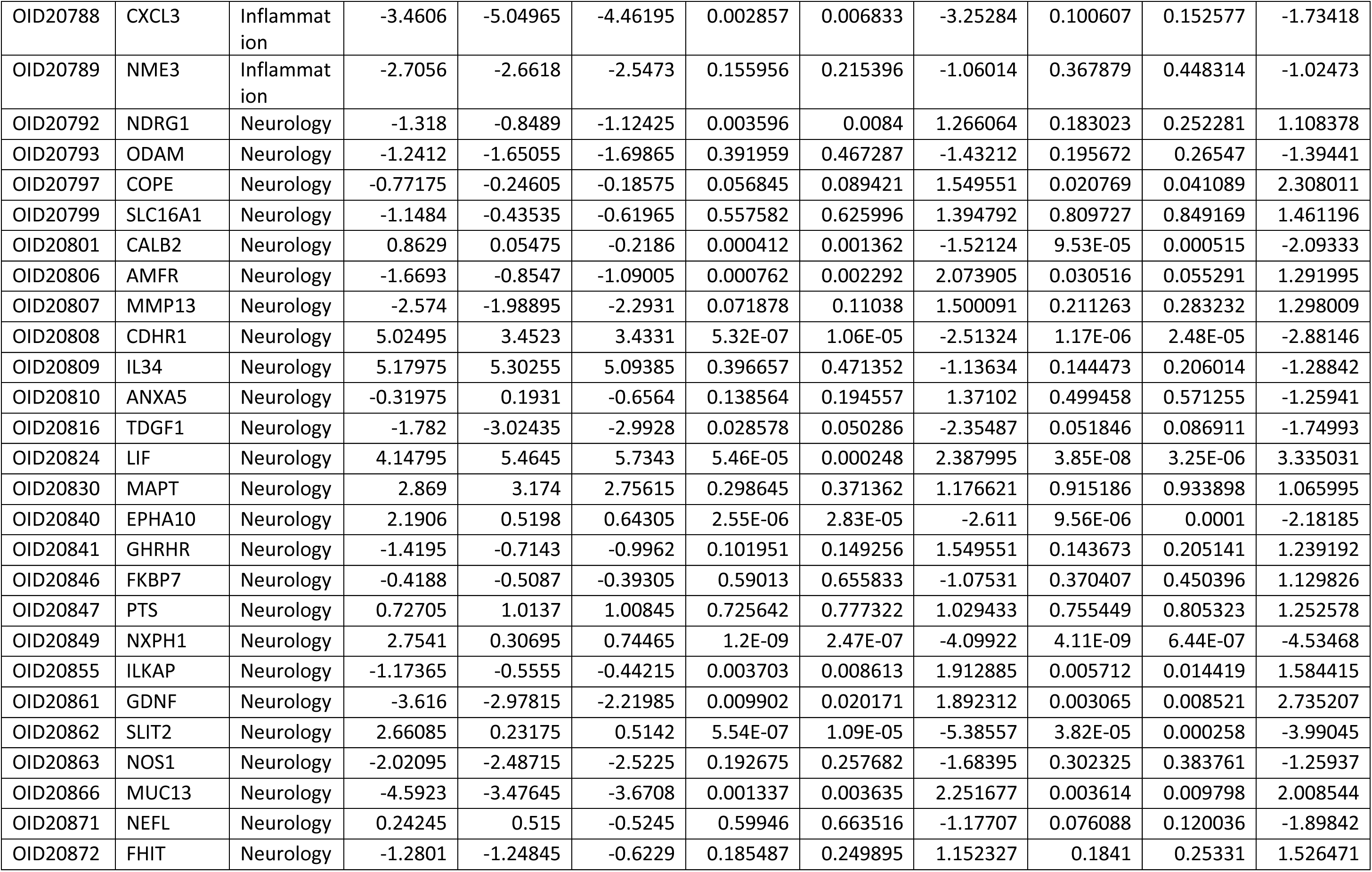
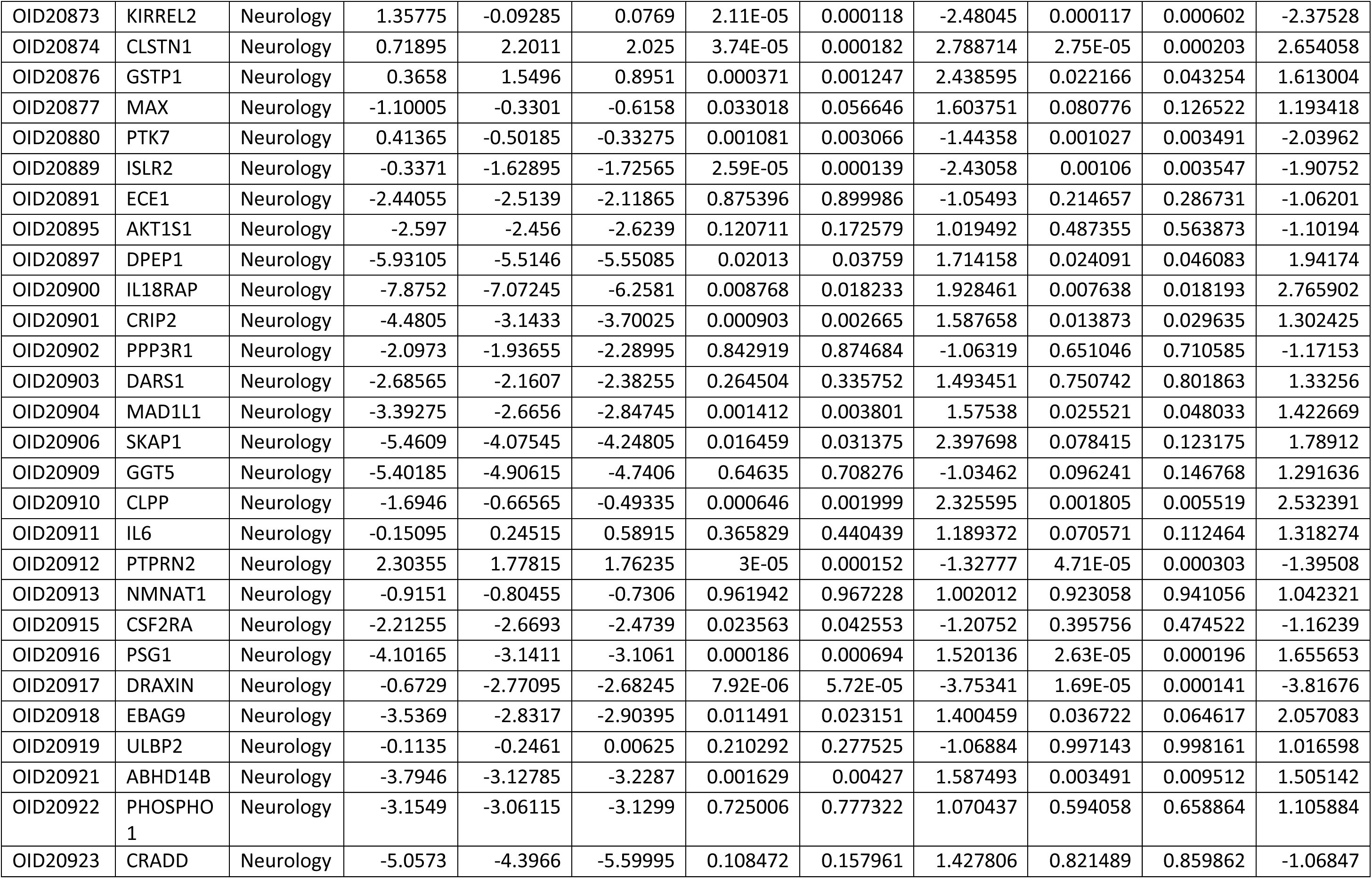
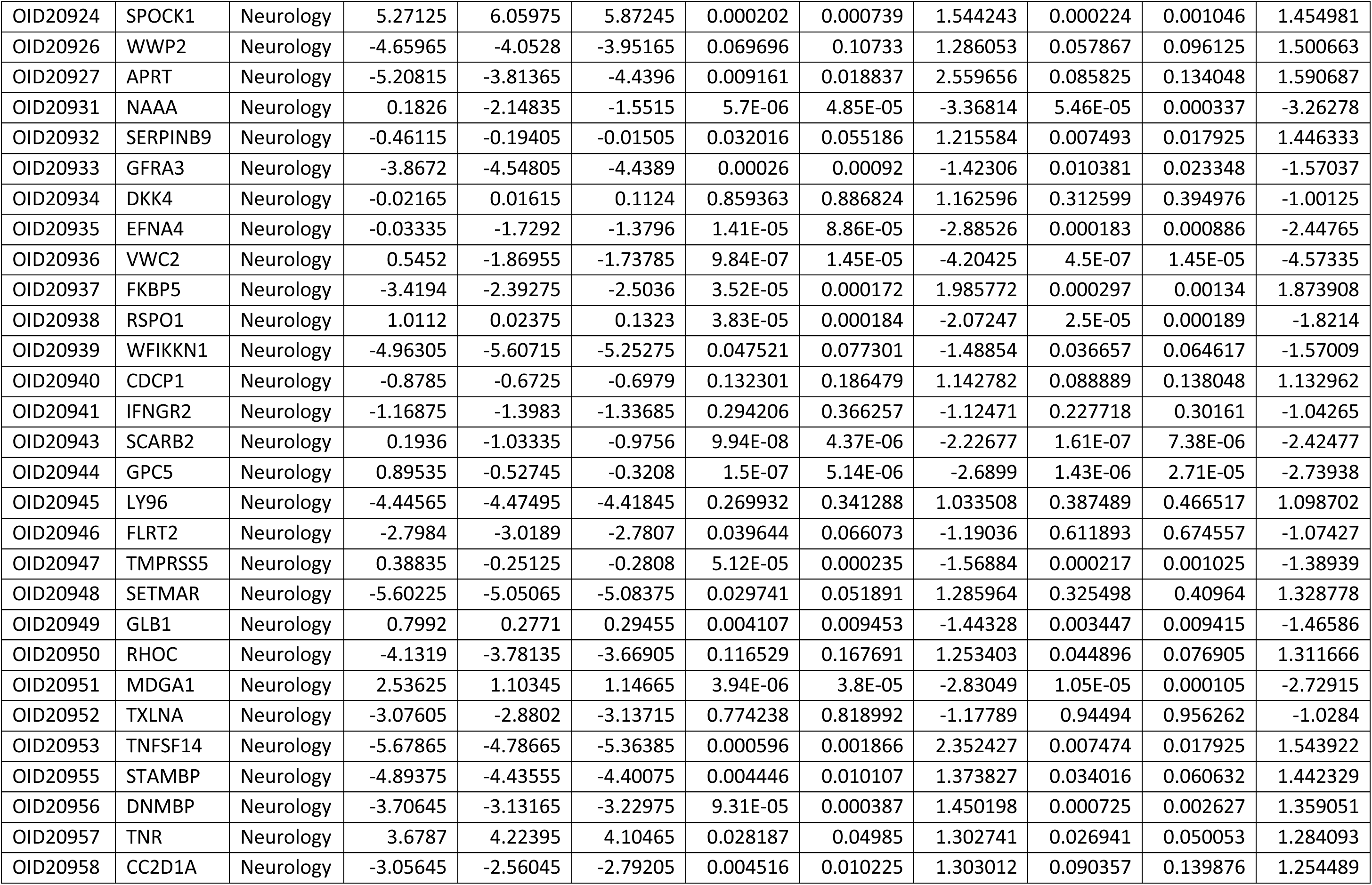
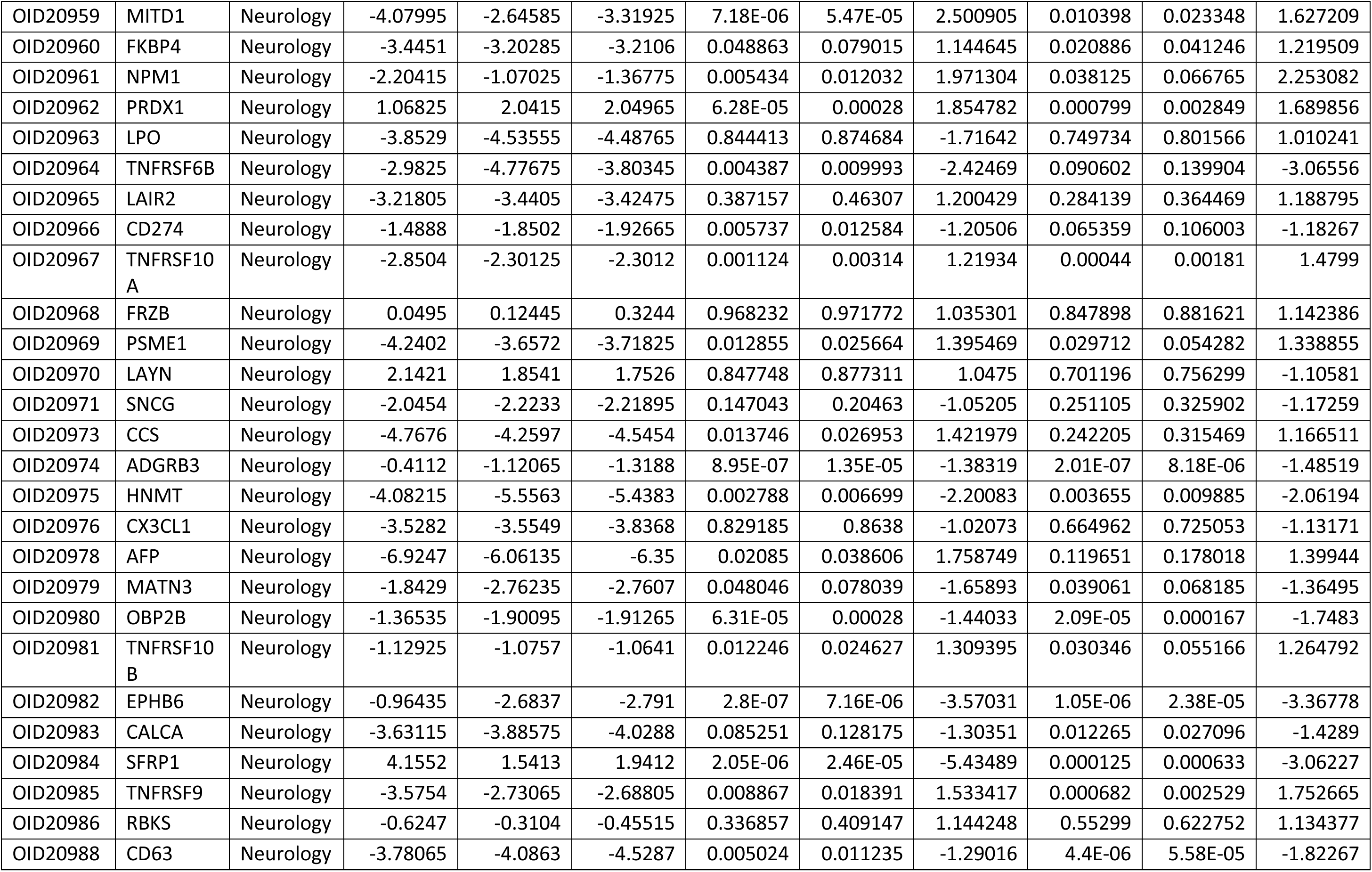
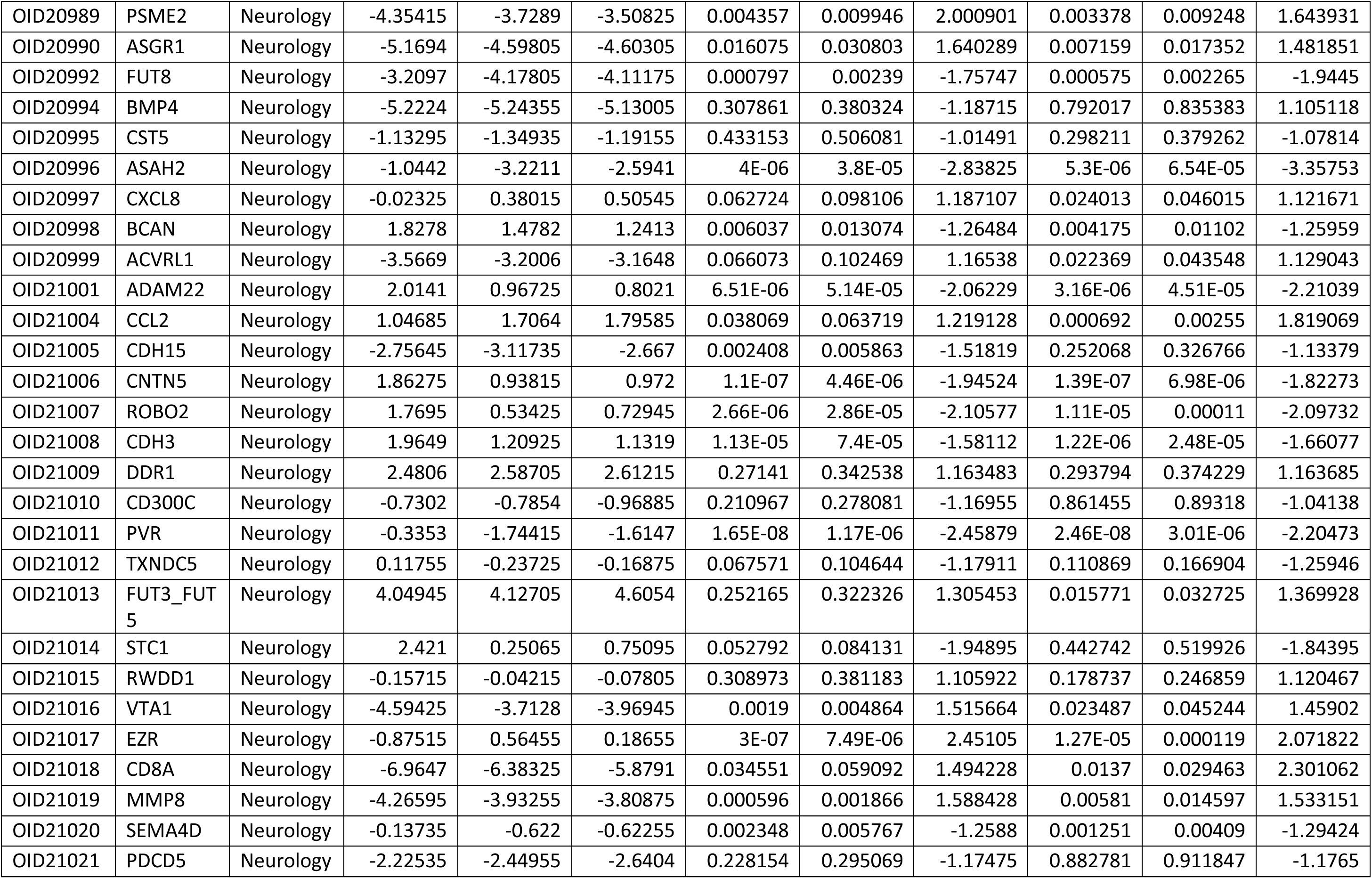
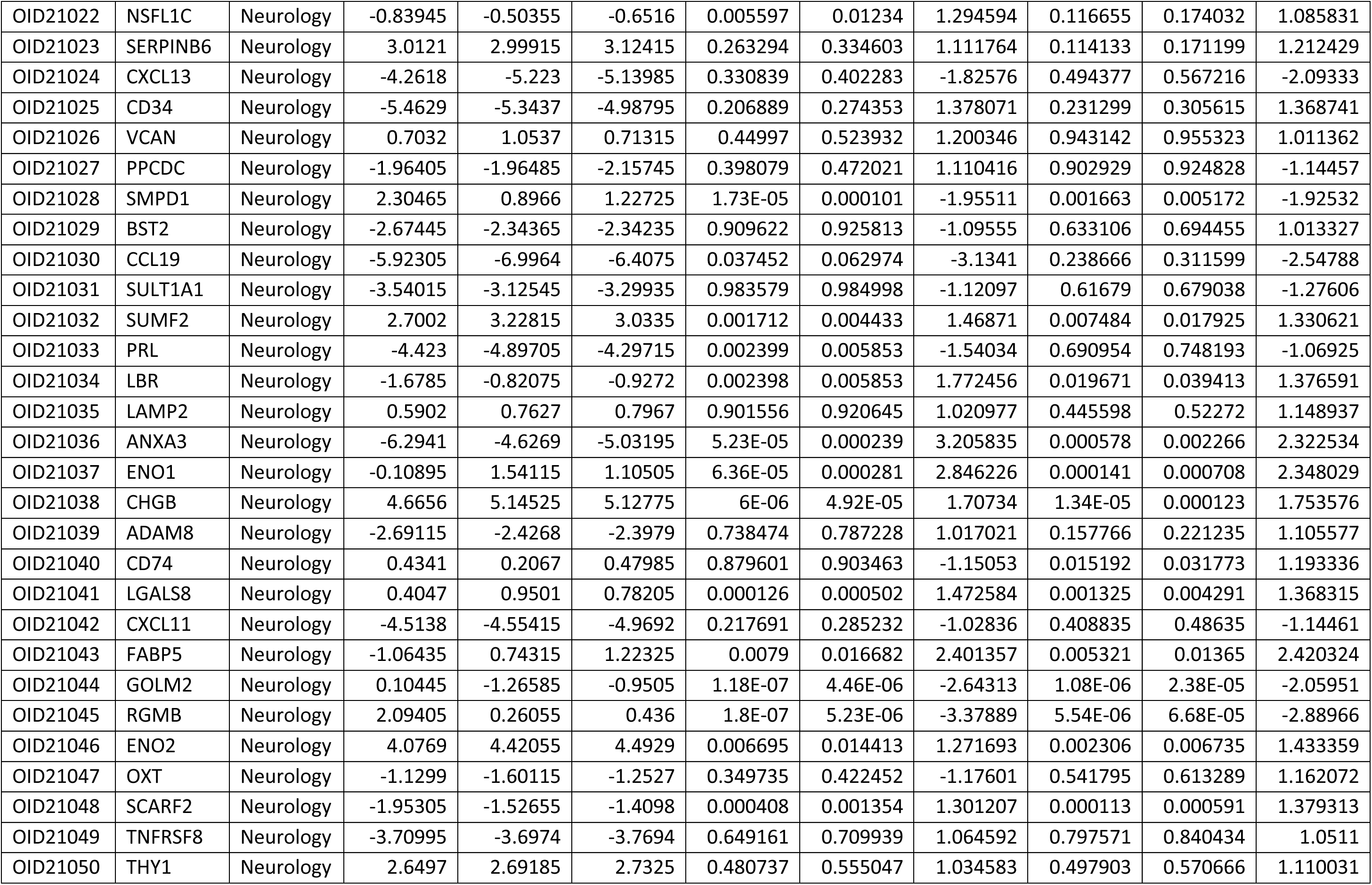
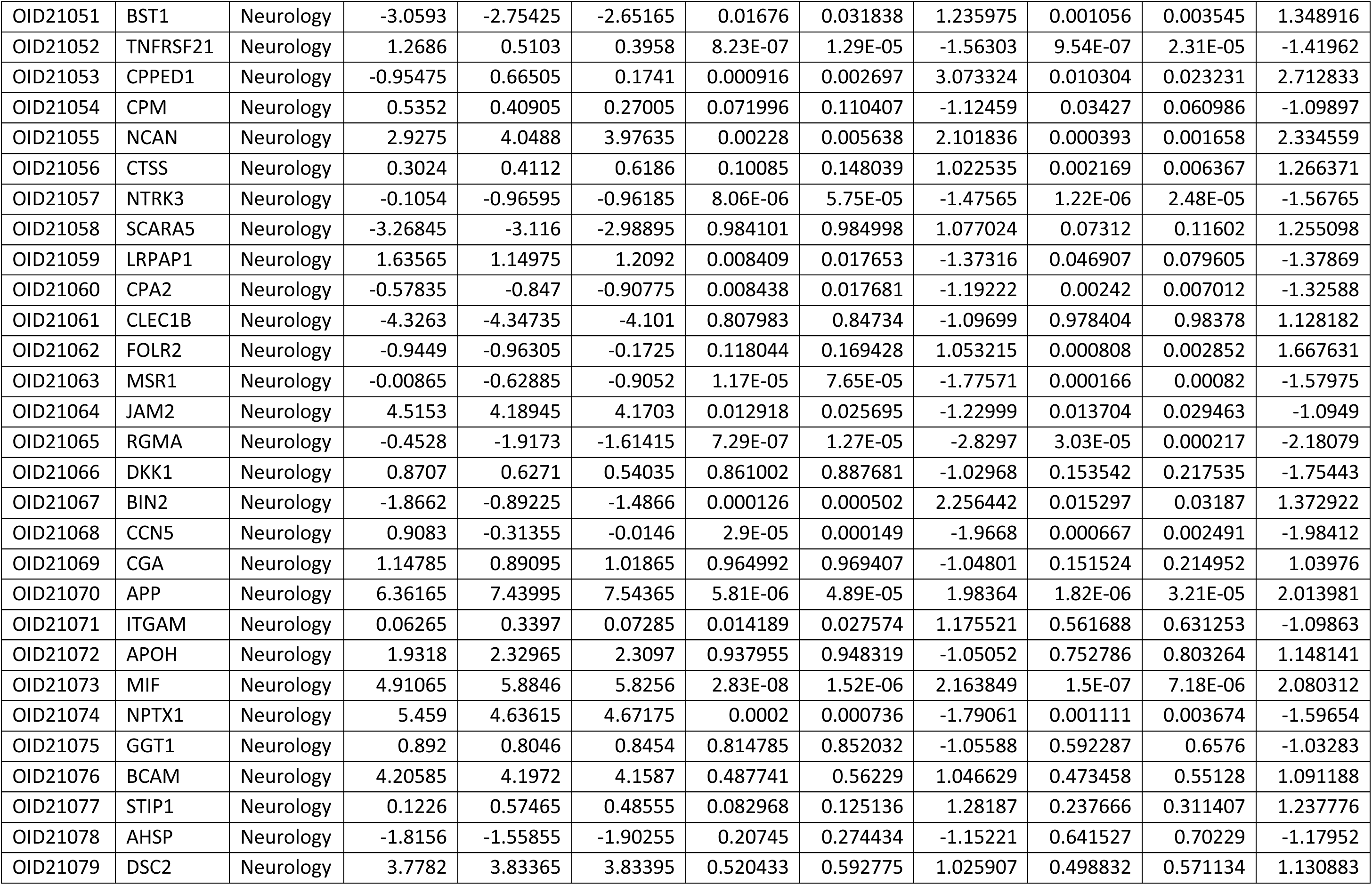
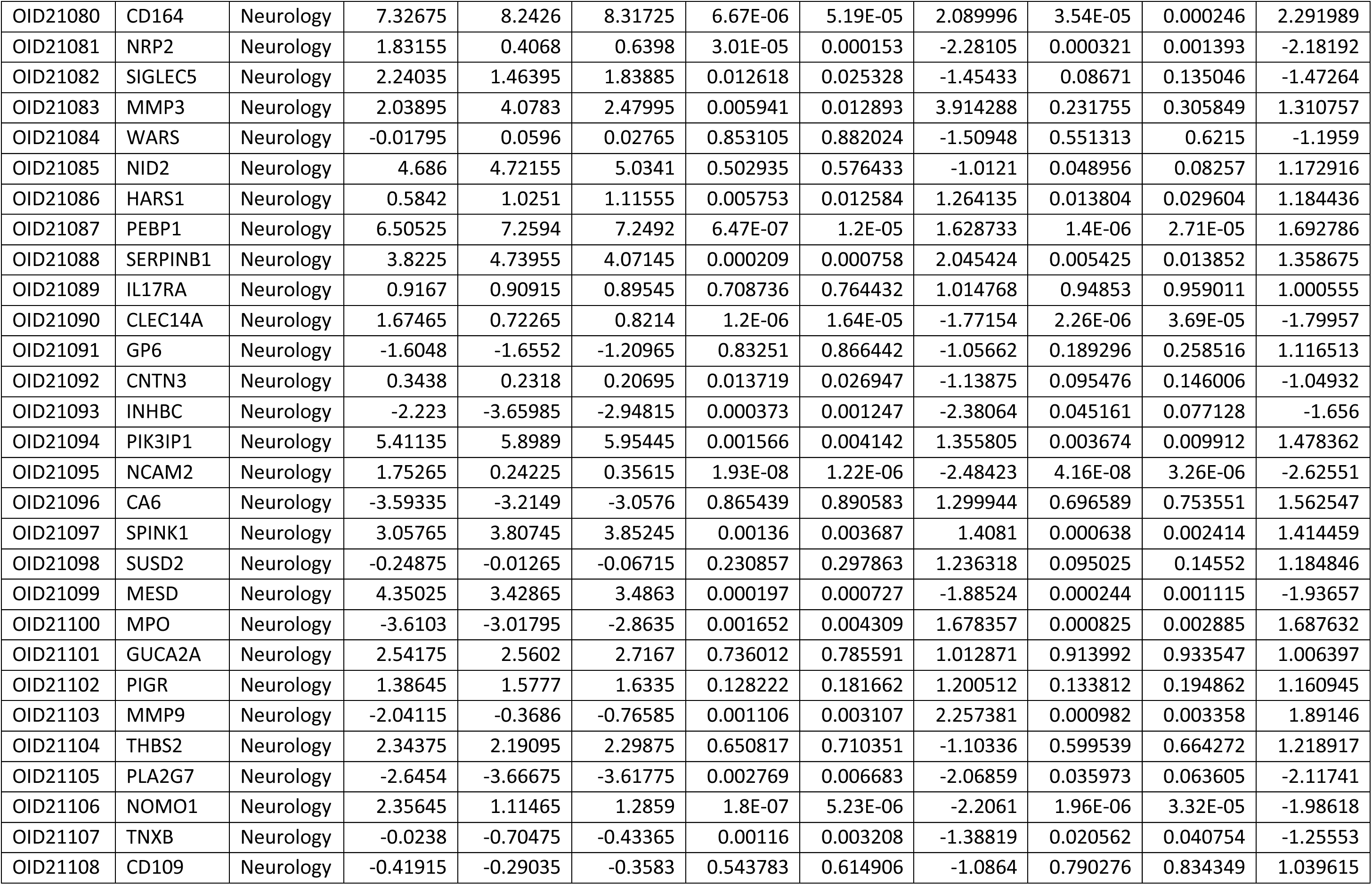
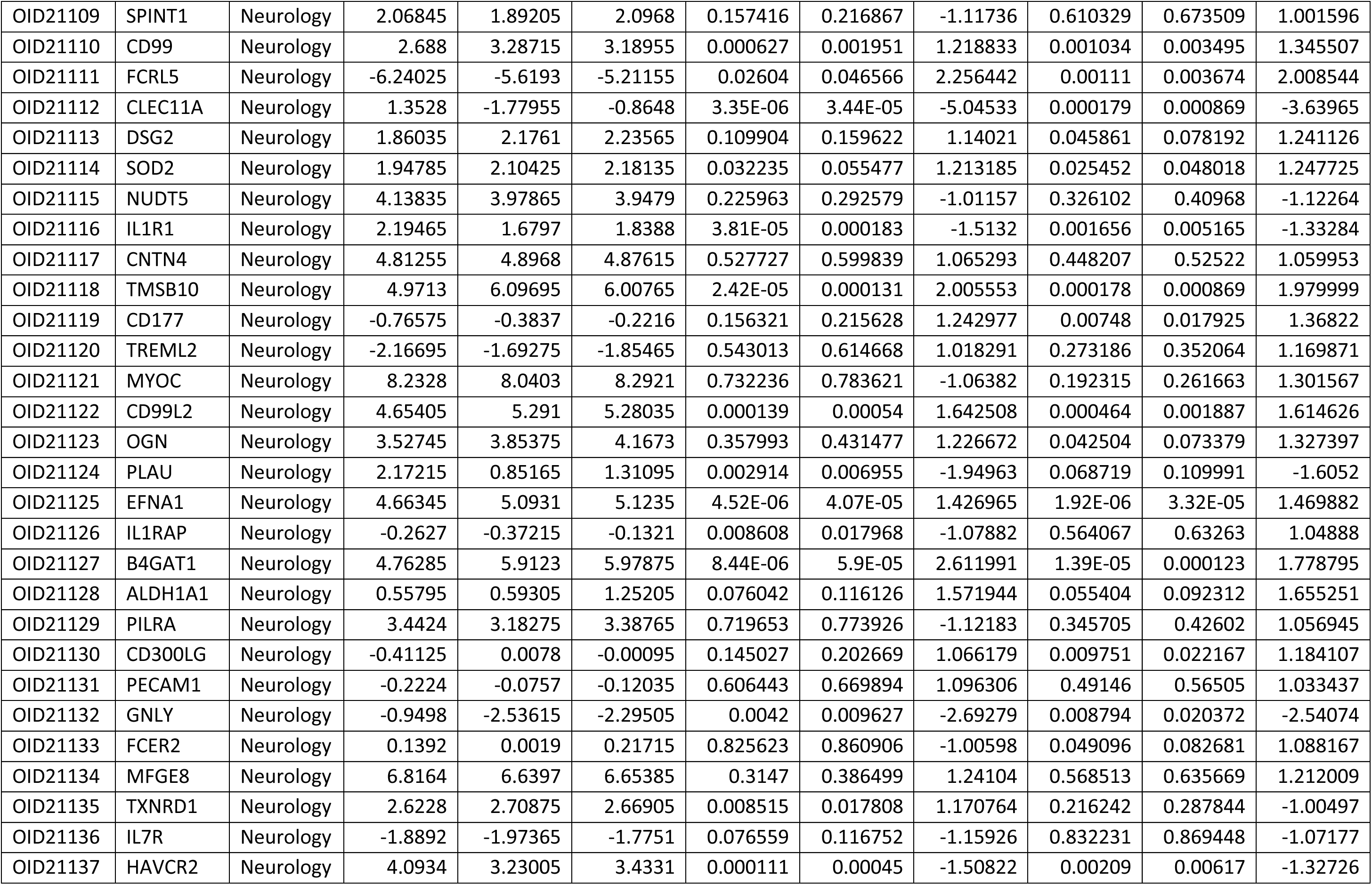
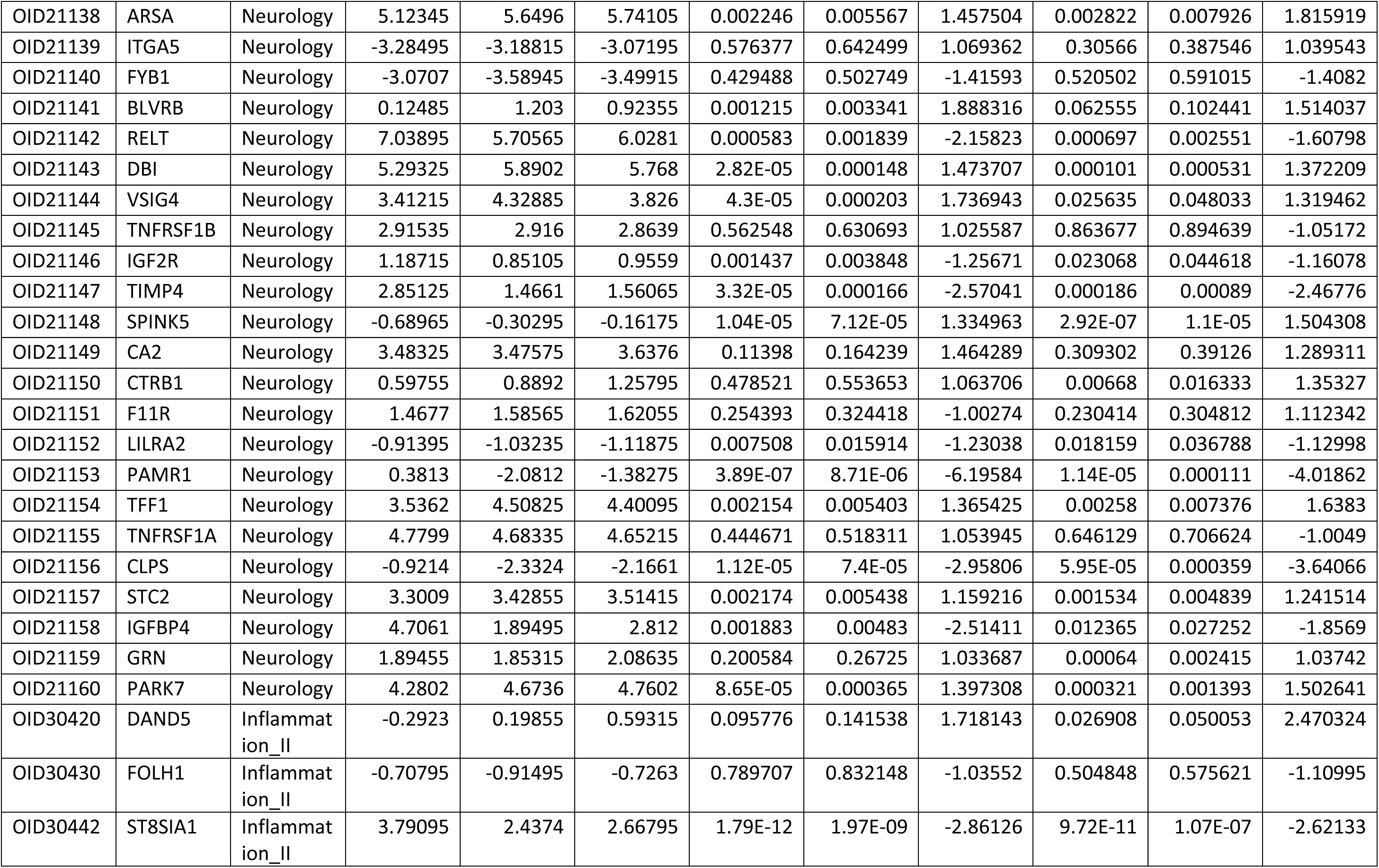
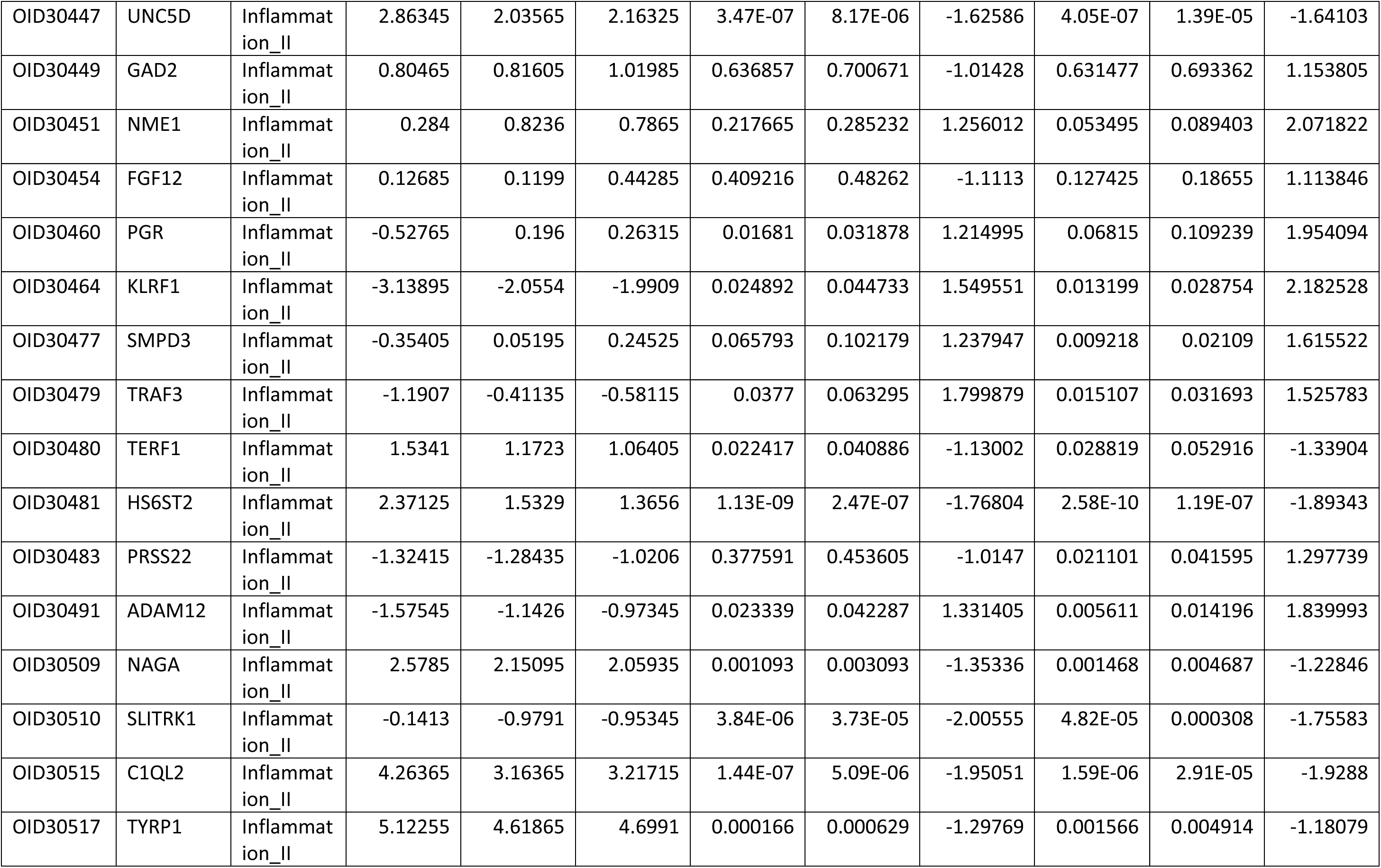
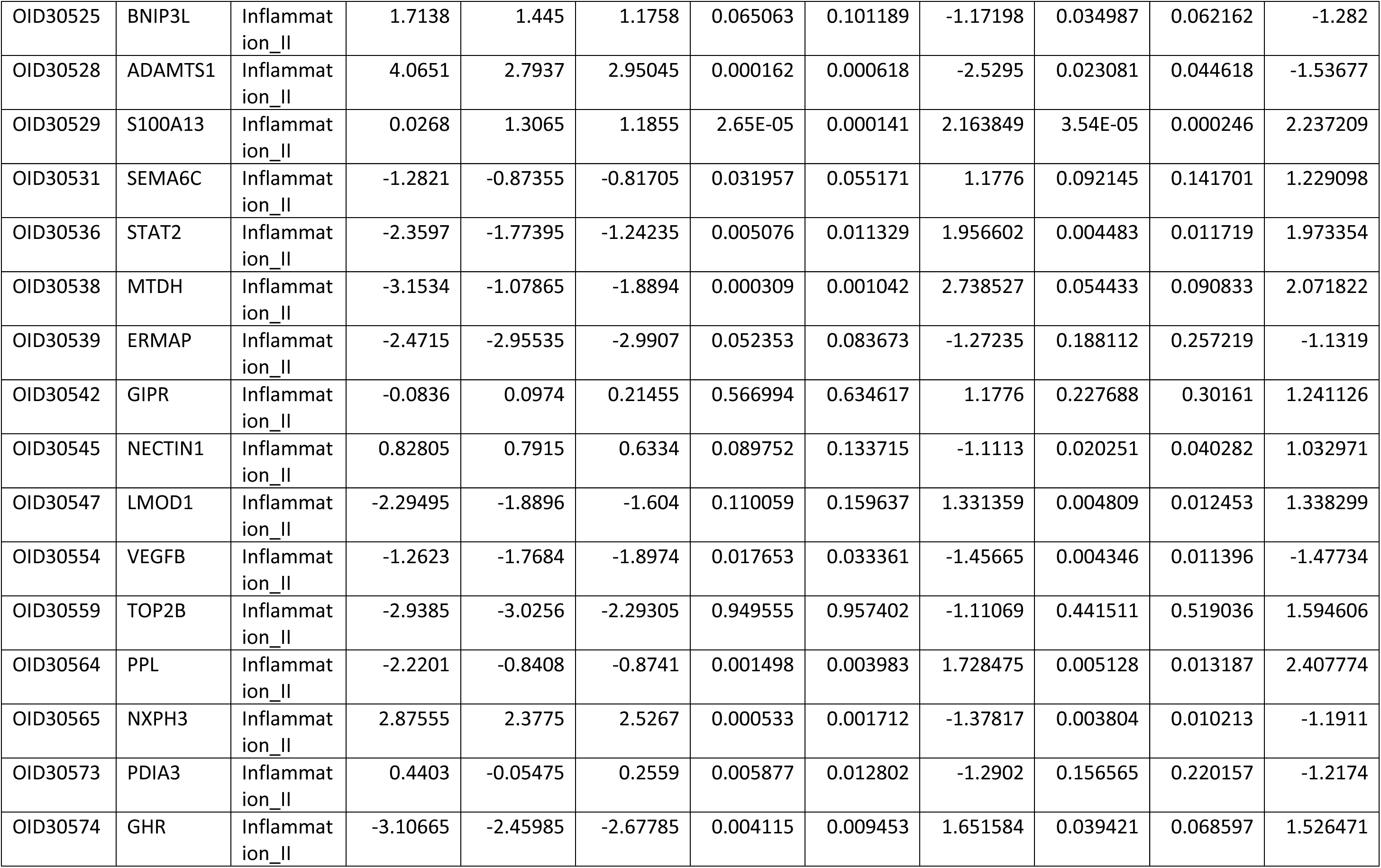
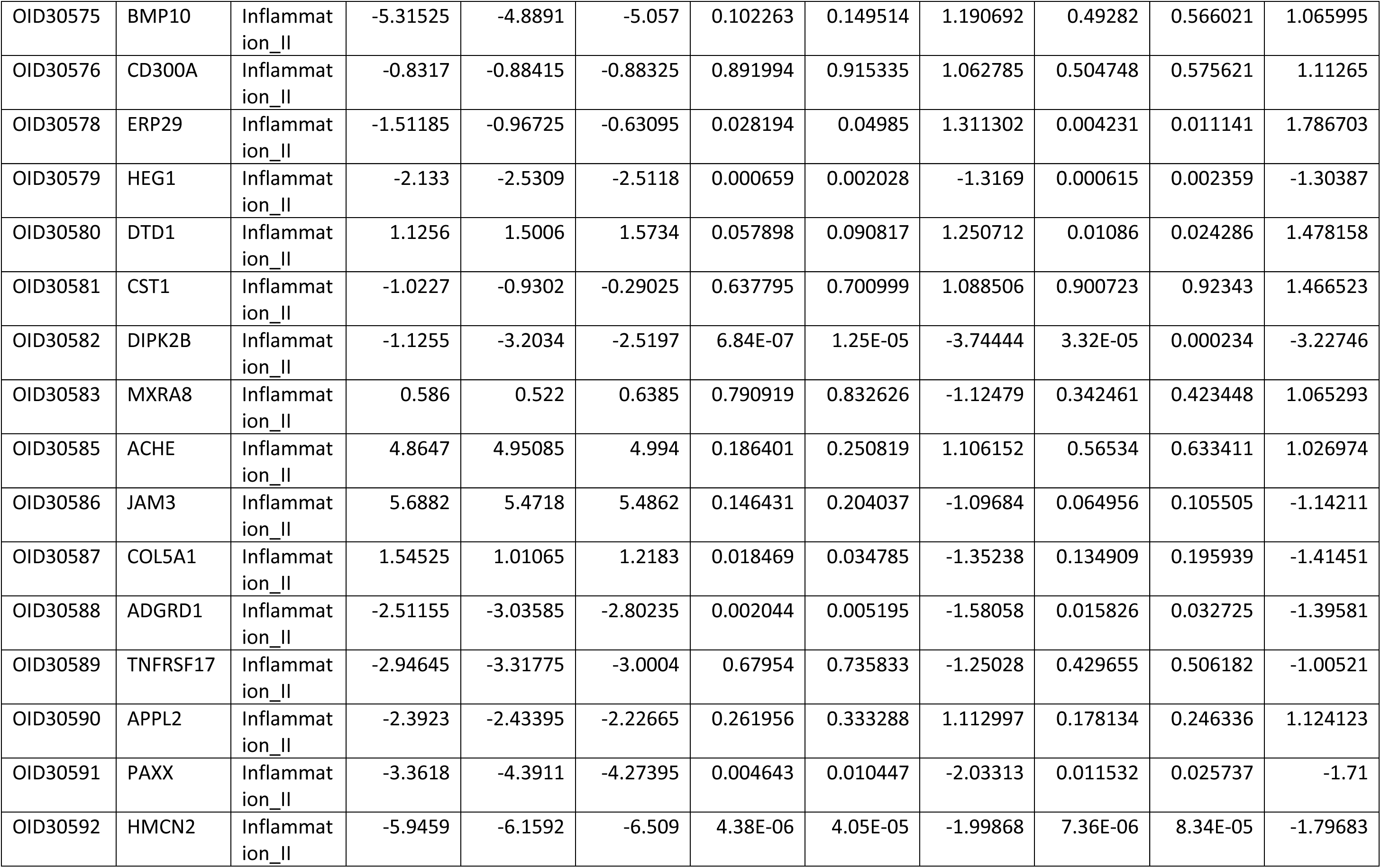
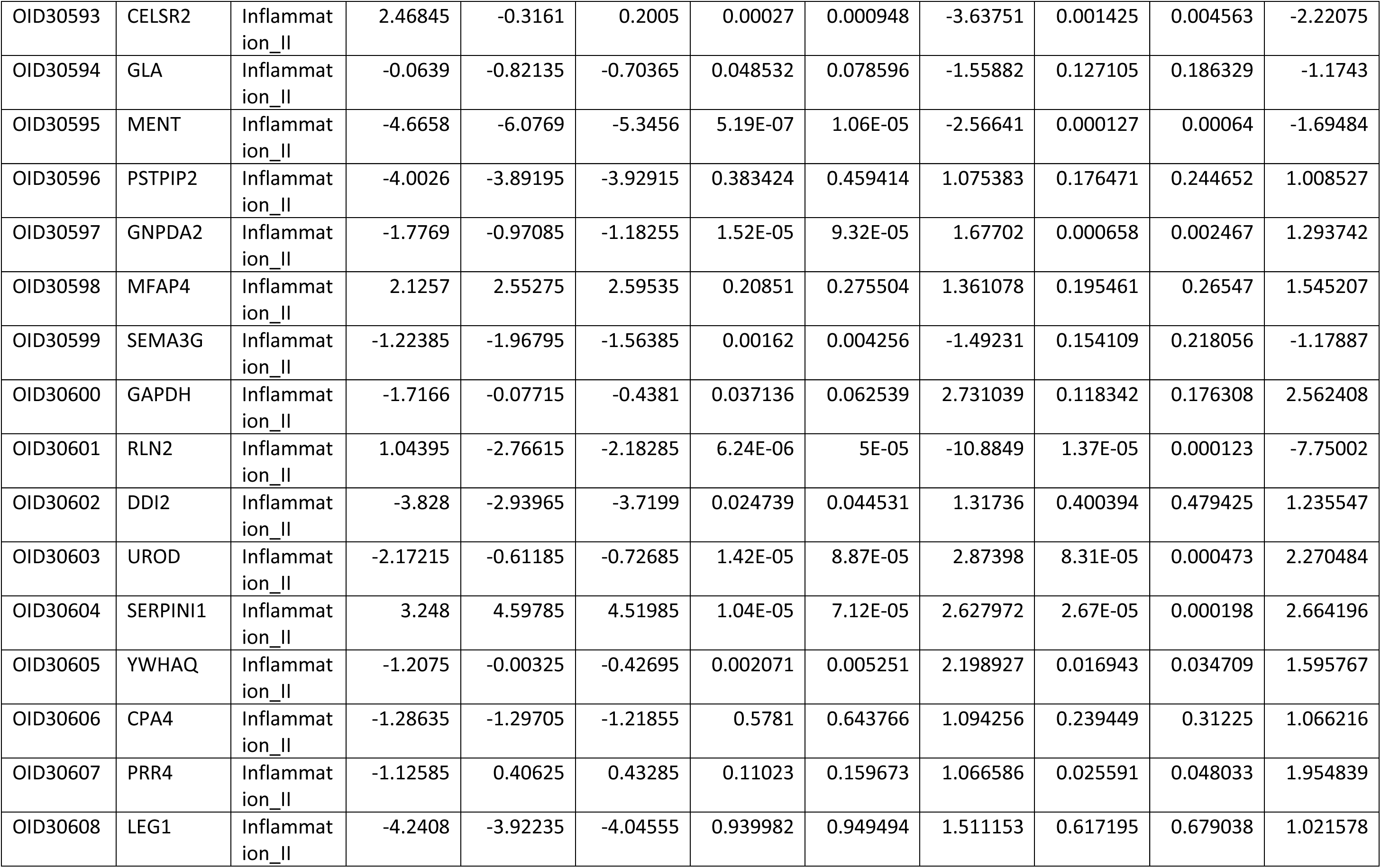
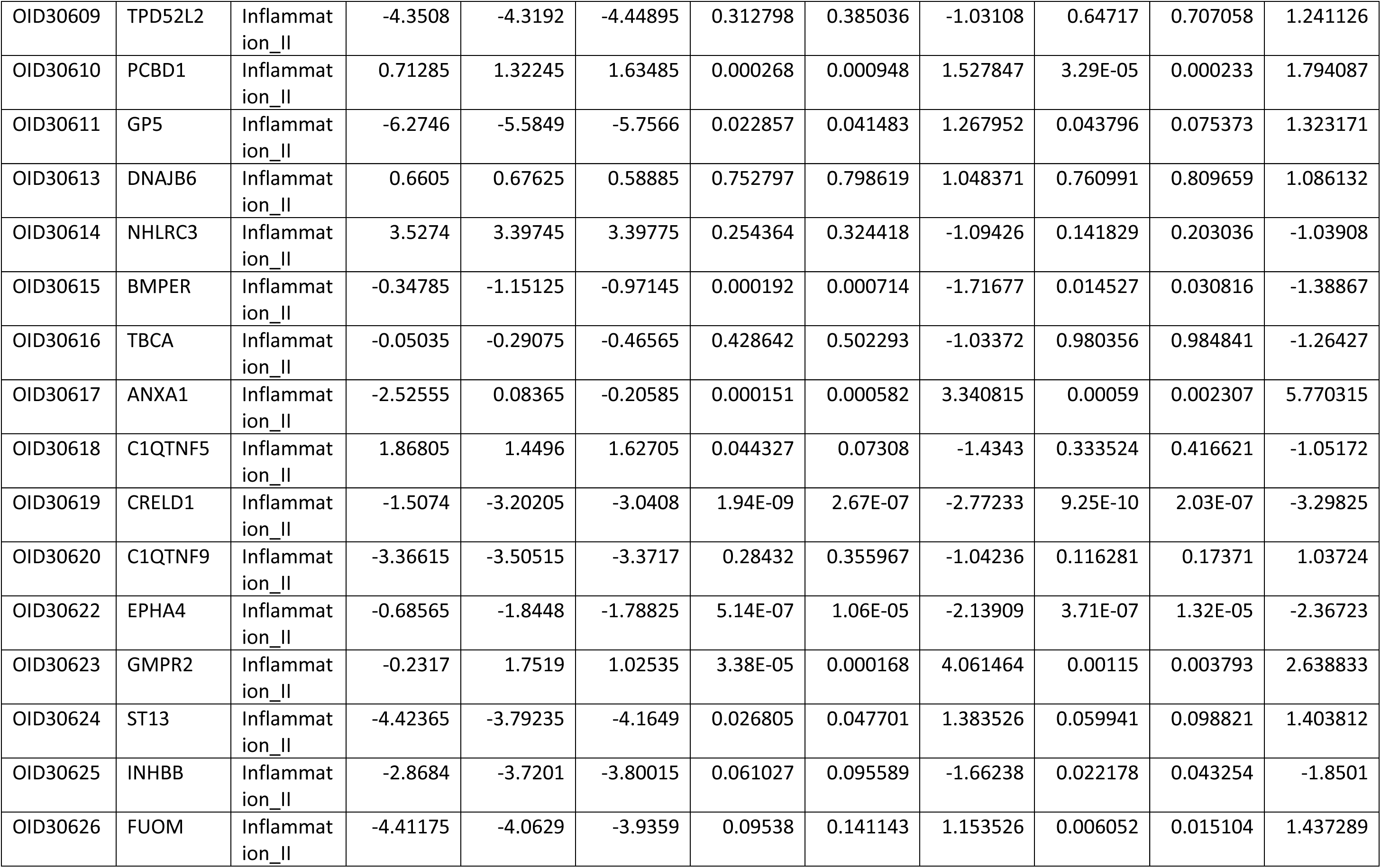
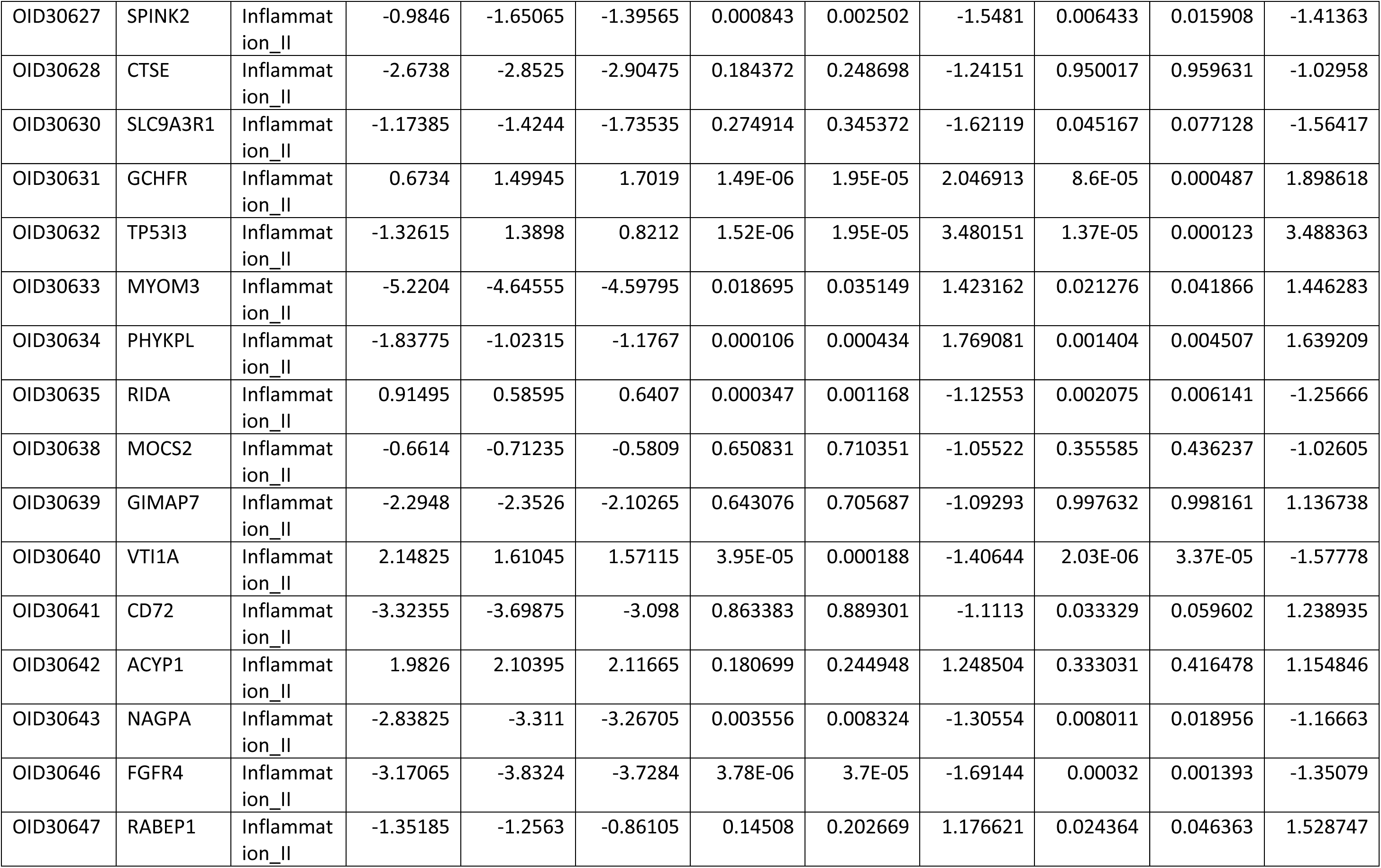
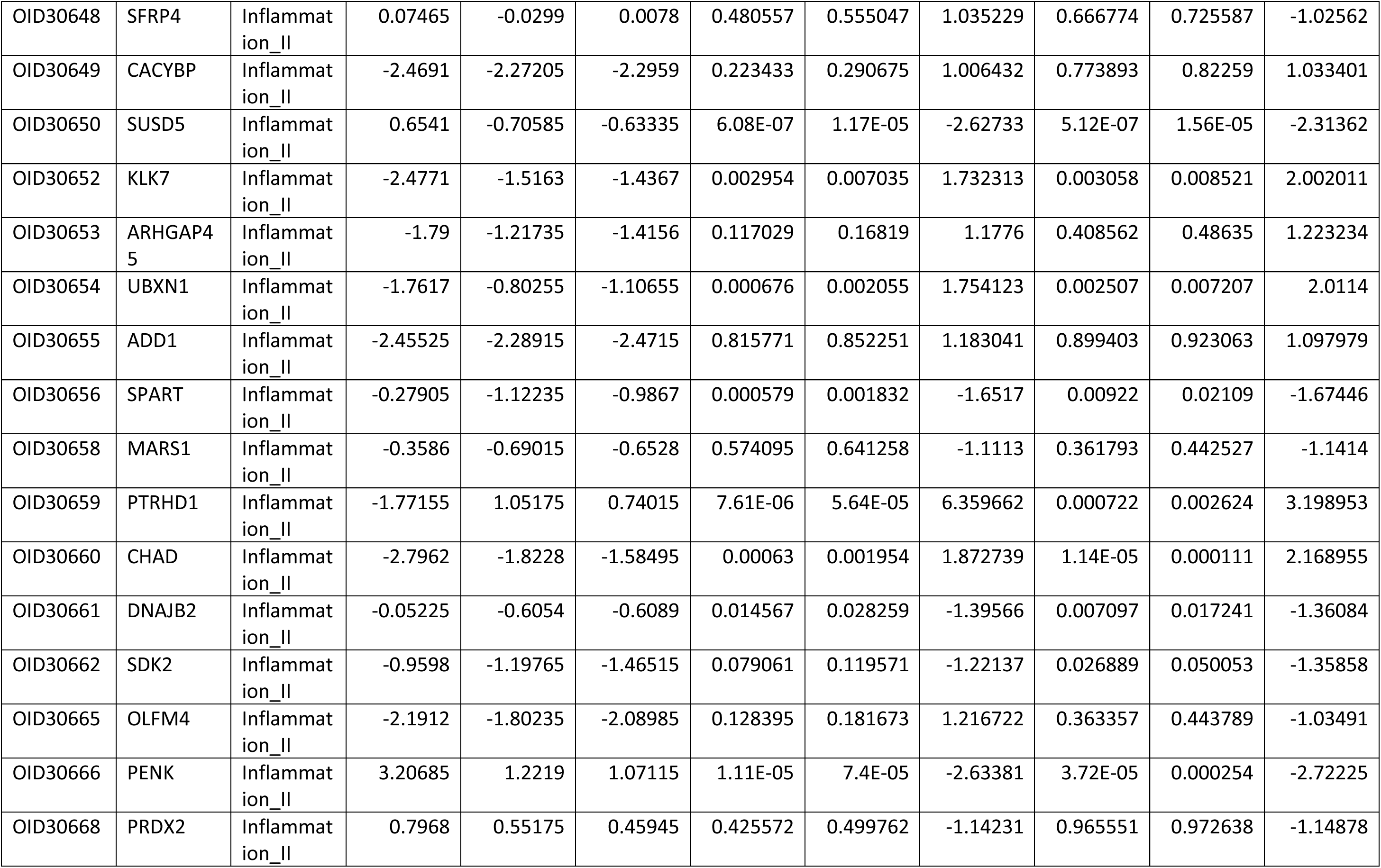
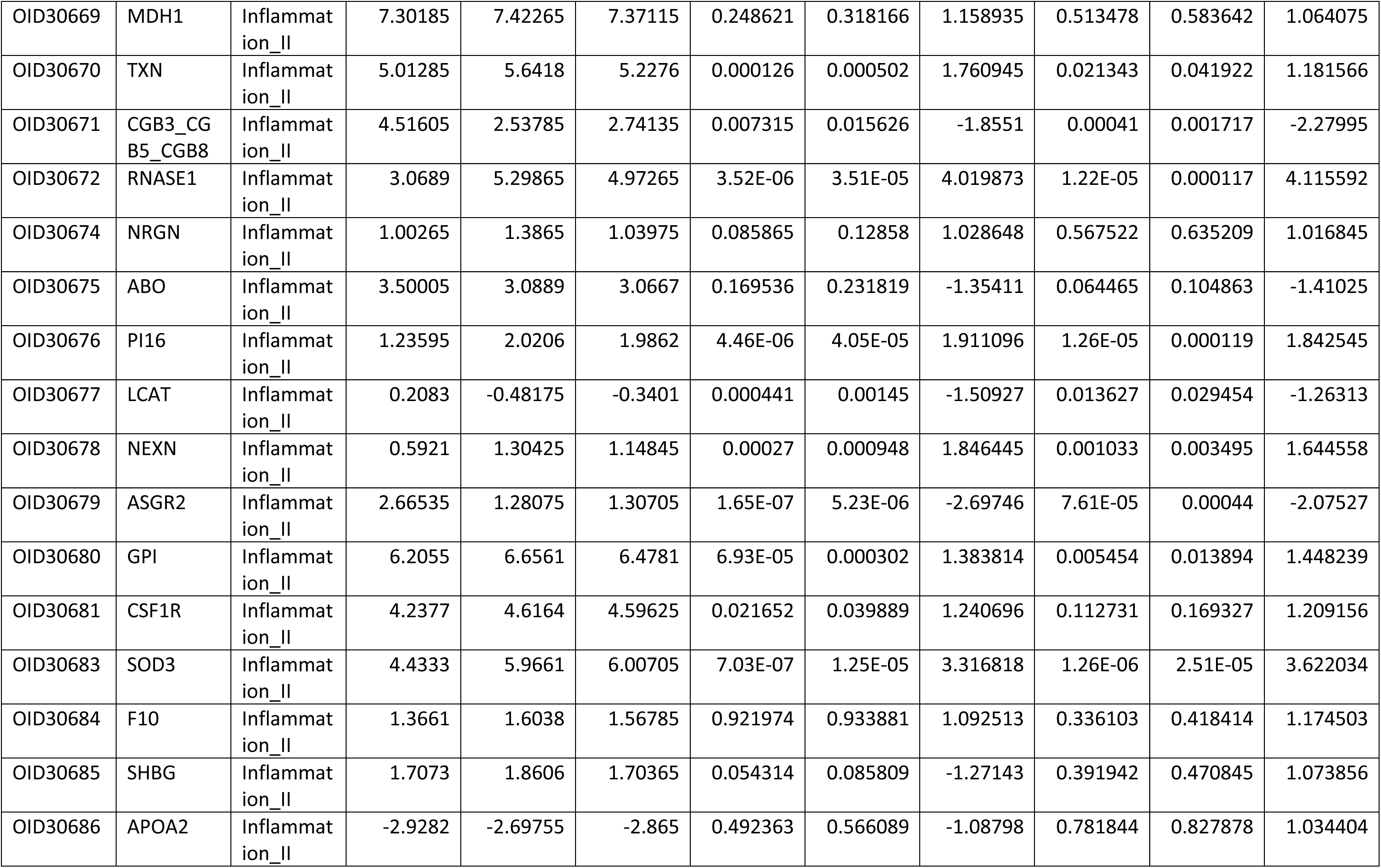
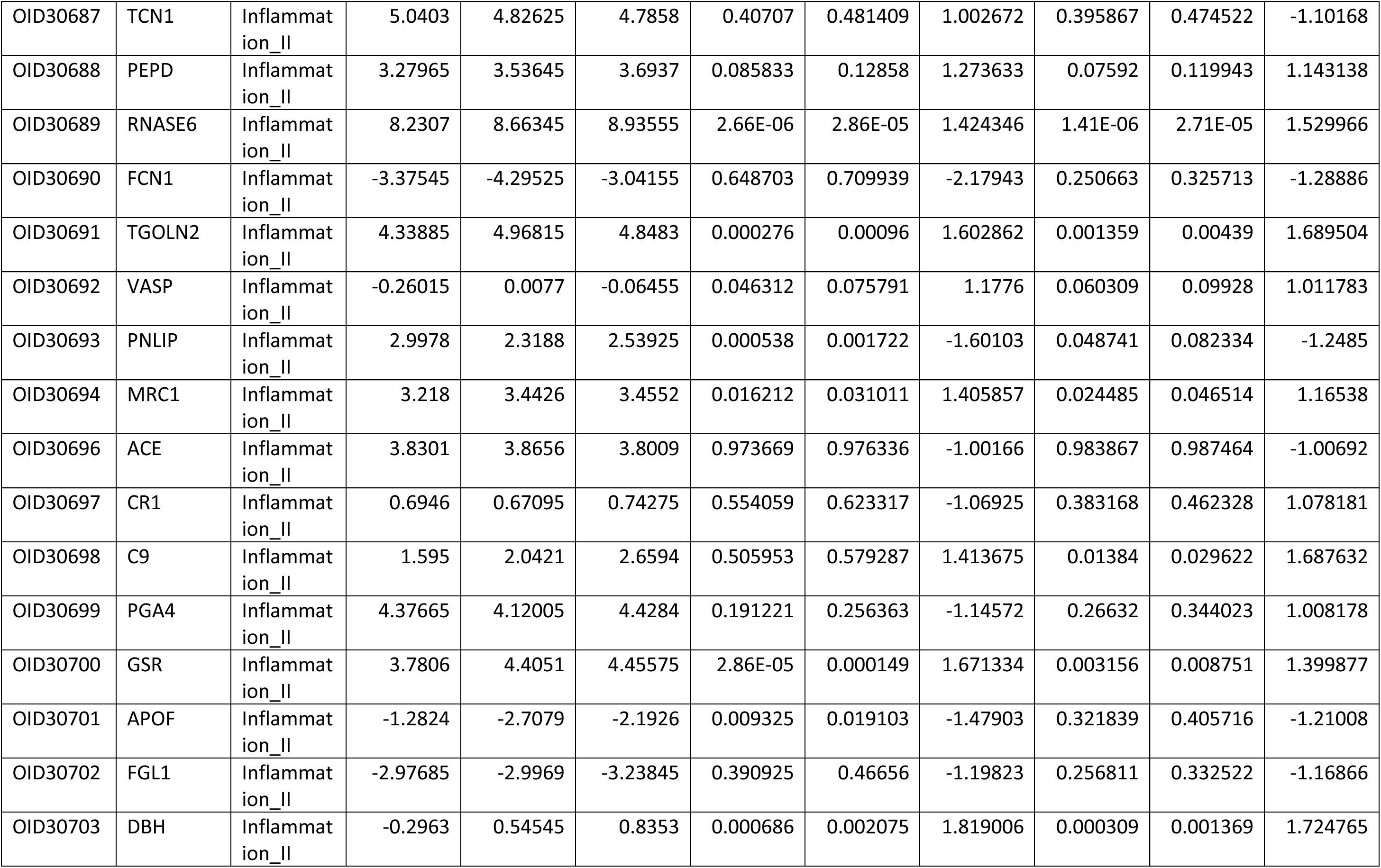
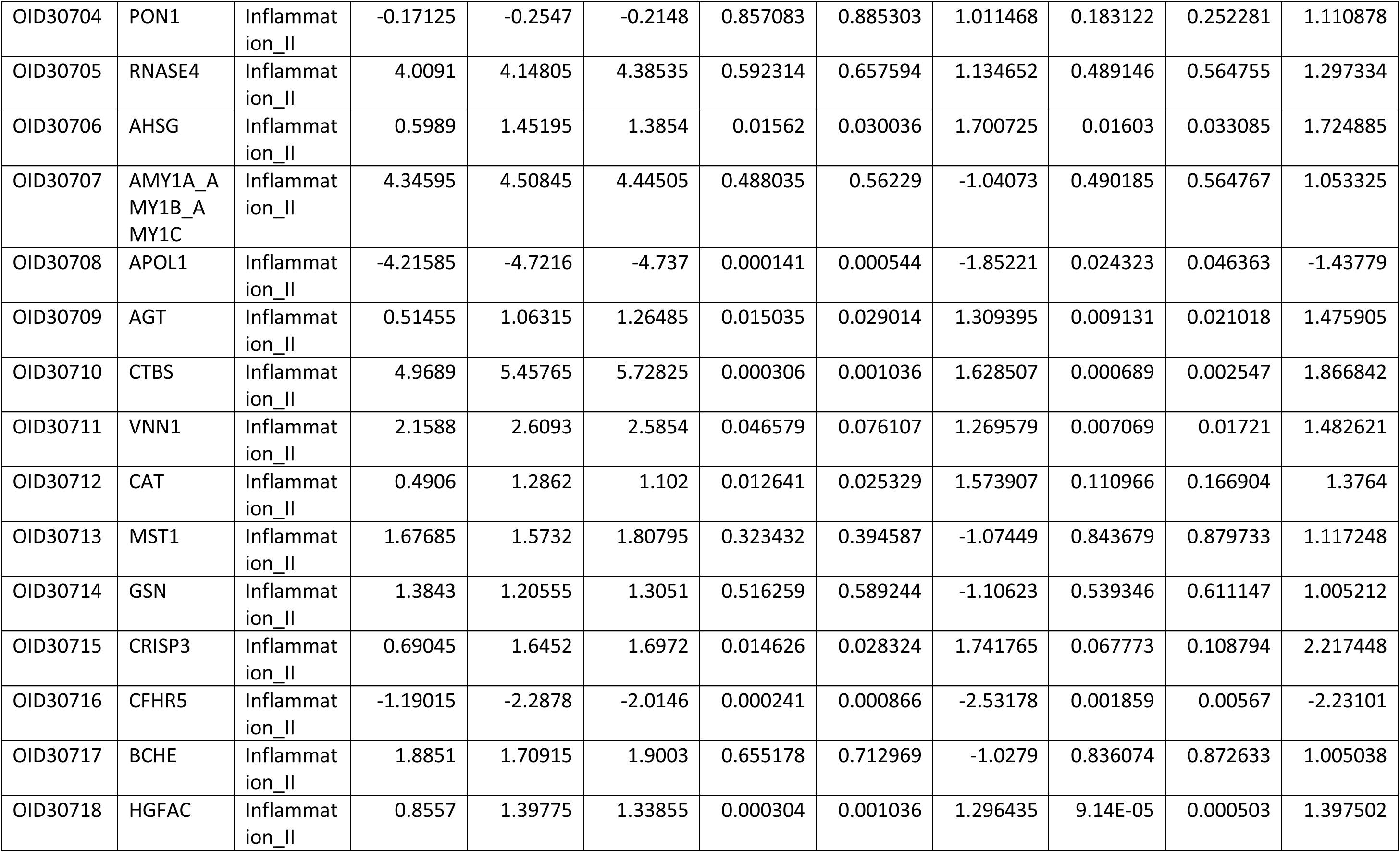
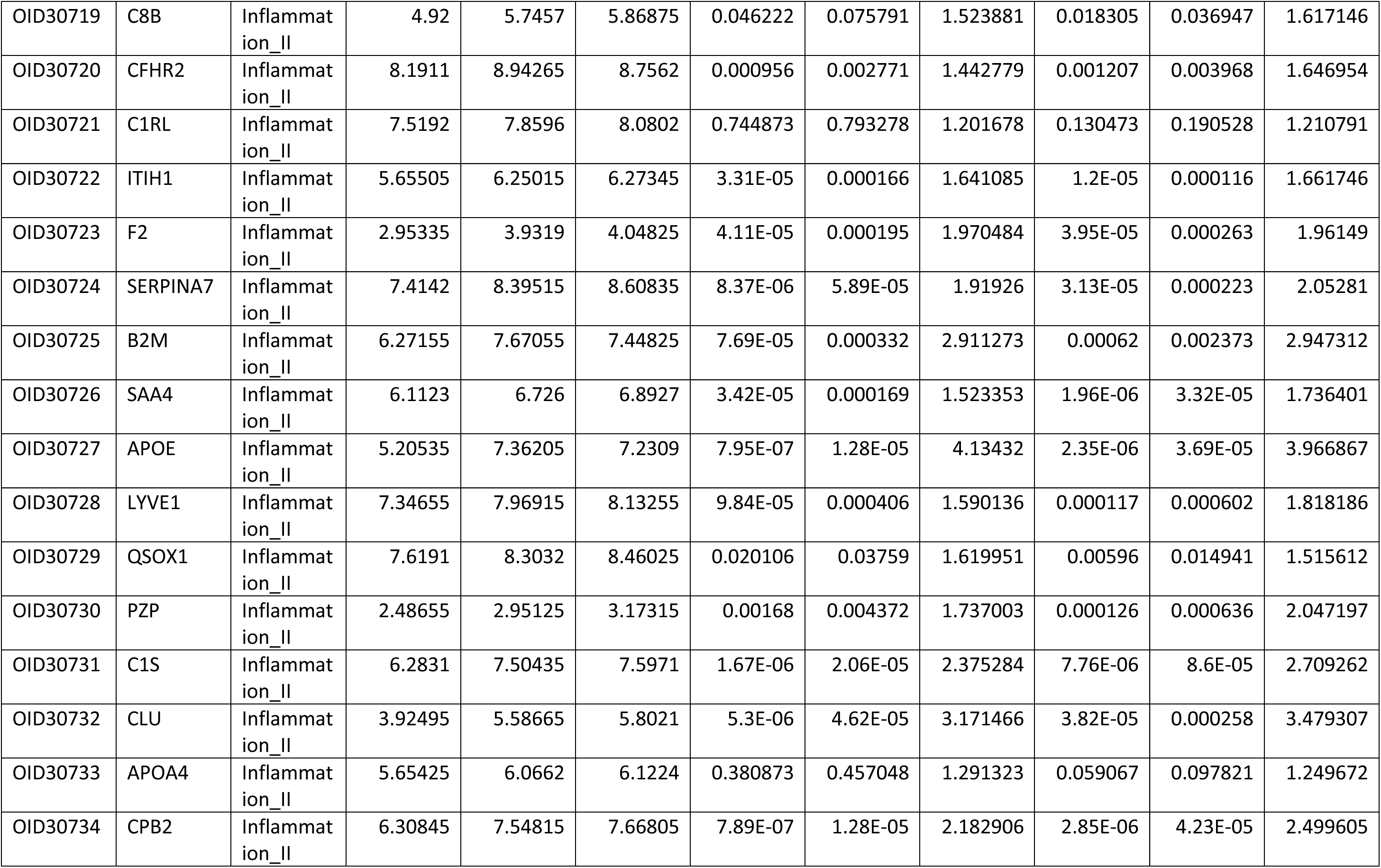
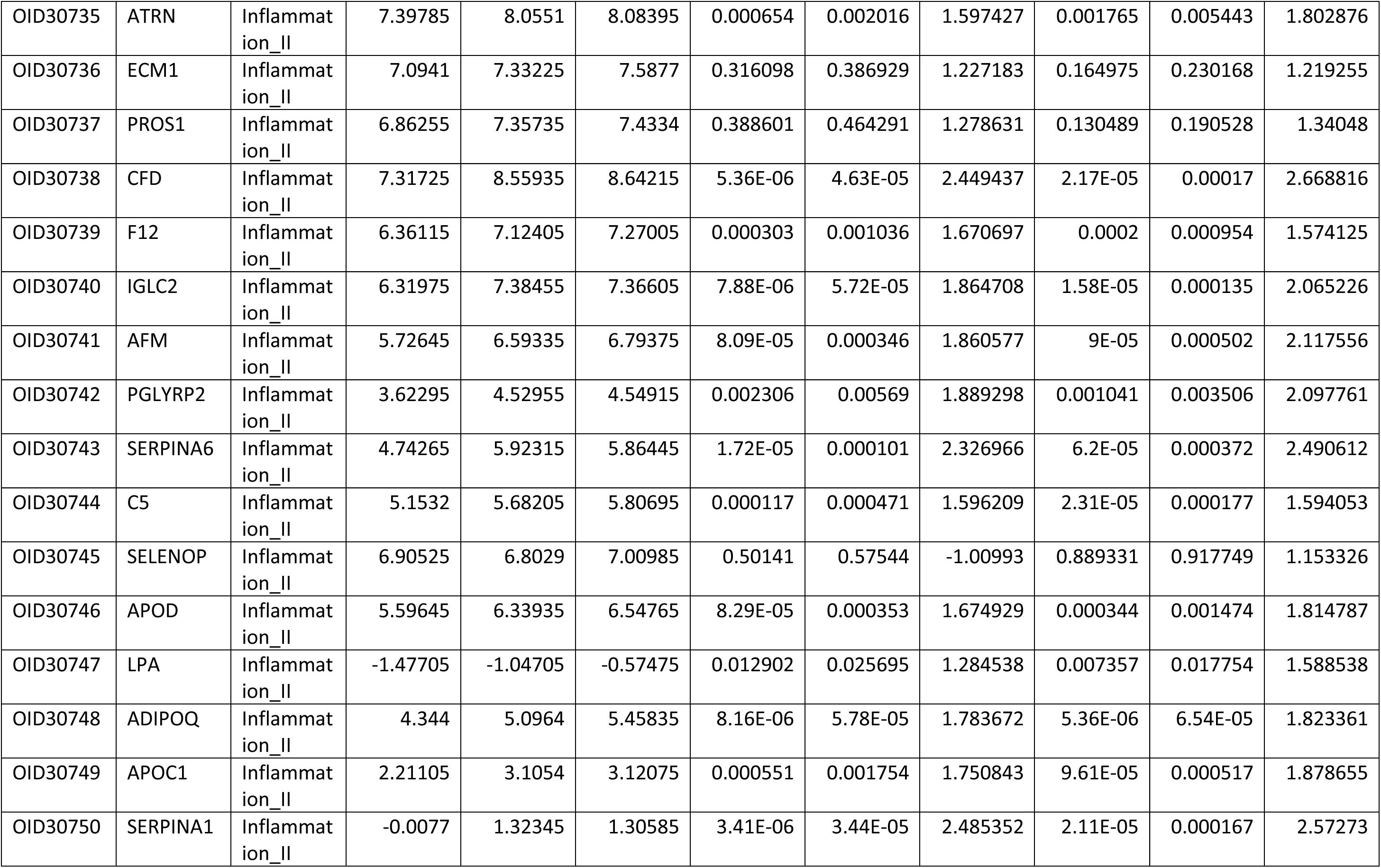
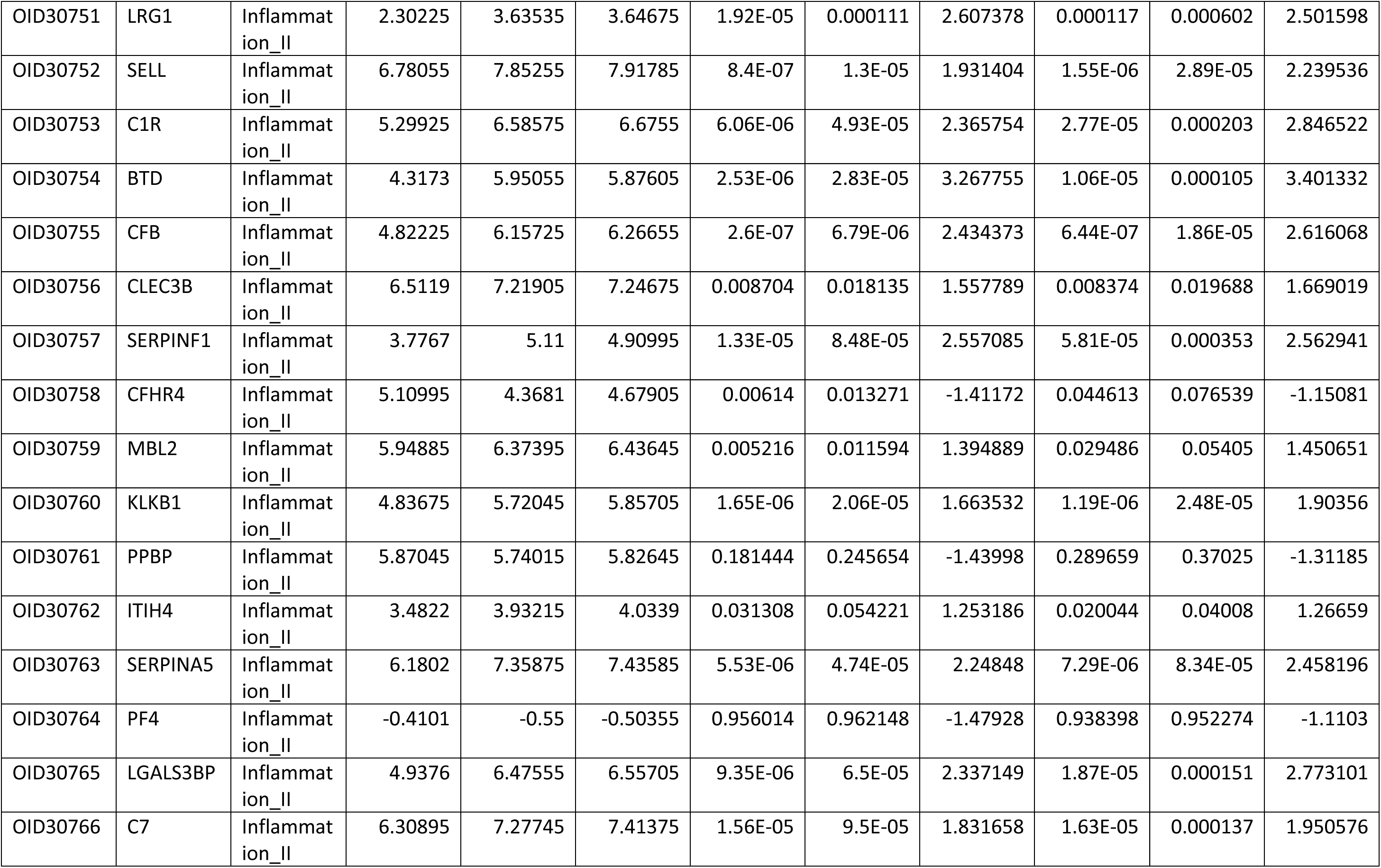
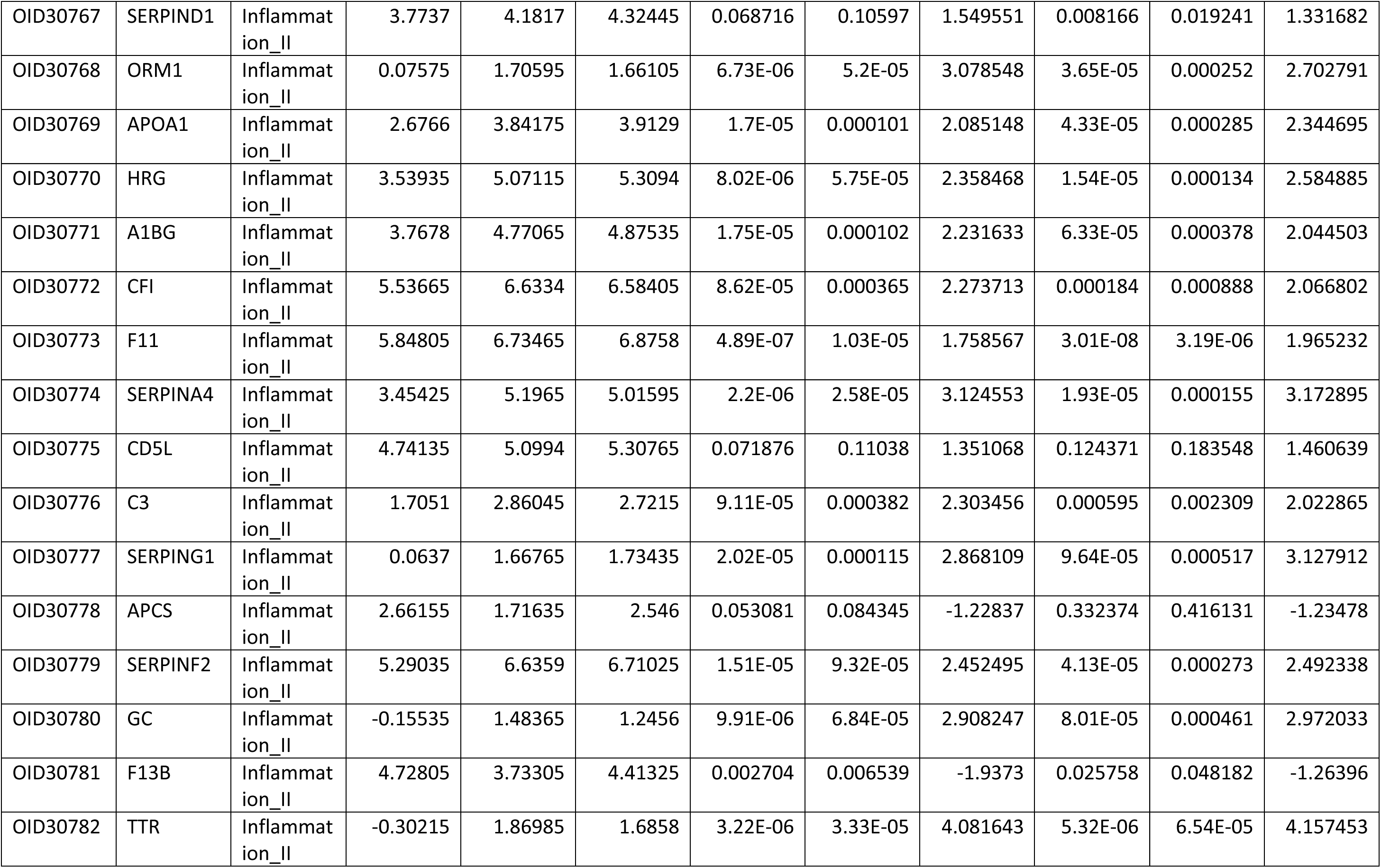
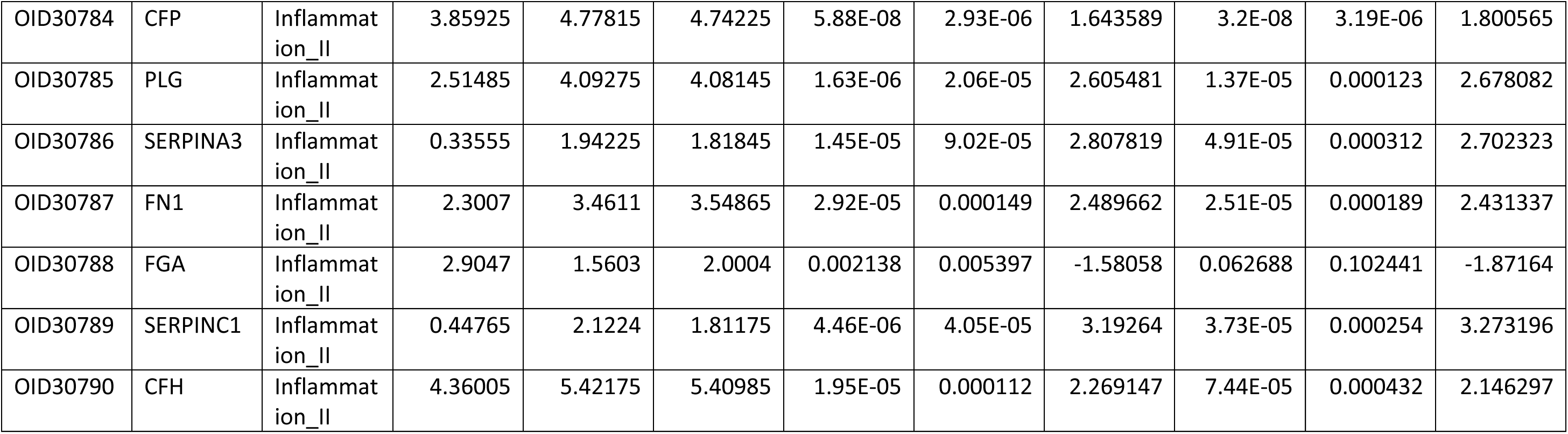
Temporal change in protein levels as evaluated using the Olink platform. Spreadsheet listing Olink (https://olink.com/) protein identifiers (IDs) that passed quality control alongside metrics for differential expression in vitreous humor comparing baseline to post-treatment at week 12 and week 36 timepoints. fdr = false discovery rate; NPX = normalised protein expression.

## References

1. Wong WL, Su X, Li X, et al. Global prevalence of age-related macular degeneration and disease burden projection for 2020 and 2040: A systematic review and meta-analysis. Lancet Glob Health. 2014;2(2). doi:10.1016/S2214-109X(13)70145-1

2. Ambati J, Fowler BJ. Mechanisms of age-related macular degeneration. Neuron. 2012;75(1):26–39. doi:10.1016/j.neuron.2012.06.018

3. Patnaik JL, Lynch AM, Pecen PE, et al. The impact of advanced age-related macular degeneration on the National Eye Institute’s Visual Function Questionnaire-25. Acta Ophthalmol. 2021;99(7):750–755. doi:10.1111/aos.14731

4. Patel PJ, Ziemssen F, Ng E, et al. Burden of illness in geographic atrophy: A study of vision-related quality of life and health care resource use. Clinical Ophthalmology. 2020;14:15–28. doi:10.2147/OPTH.S226425

5. Heier JS, Lad EM, Holz FG, et al. Pegcetacoplan for the treatment of geographic atrophy secondary to age-related macular degeneration (OAKS and DERBY): two multicentre, randomised, double-masked, sham-controlled, phase 3 trials. The Lancet. 2023;402(10411):1434–1448. doi:10.1016/S0140-6736(23)01520-9

6. Patel SS, Lally DR, Hsu J, et al. Avacincaptad pegol for geographic atrophy secondary to age-related macular degeneration: 18-month findings from the GATHER1 trial. Eye (Basingstoke). 2023;37(17):3551–3557. doi:10.1038/s41433-023-02497-w

7. Tzoumas N, Riding G, Williams MA, Steel DHW. Complement inhibitors for age-related macular degeneration. Cochrane Database of Systematic Reviews. 2023;2023(6). doi:10.1002/14651858.CD009300.pub3

8. Dreismann AK, McClements ME, Barnard AR, et al. Functional expression of complement factor I following AAV-mediated gene delivery in the retina of mice and human cells. Gene Ther. 2021;28(5):265–276. doi:10.1038/s41434-021-00239-9

9. Dreismann AK, Hallam TM, Tam LCS, et al. Gene targeting as a therapeutic avenue in diseases mediated by the complement alternative pathway. Immunol Rev. 2023;313(1):402–419. doi:10.1111/imr.13149

10. Nilsson B, Nilsson Ekdahl K. The tick-over theory revisited: Is C3 a contact-activated protein? Immunobiology. 2012;217(11):1106–1110. 10.1016/j.imbio.2012.07.008

11. Hallam TM, Sharp SJ, Andreadi A, Kavanagh D. Complement factor I: Regulatory nexus, driver of immunopathology, and therapeutic. Immunobiology. 2023;228(5). doi:10.1016/j.imbio.2023.152410

12. Pangburn MK, Schreiber RD, Muller-Eberhard HJ. HUMAN COMPLEMENT C3b INACTIVATOR: ISOLATION, CHARACTERIZATION, AND DEMONSTRATION OF AN ABSOLUTE REQUIREMENT FOR THE SERUM PROTEIN fllH FOR CLEAVAGE OF C3b AND C4b IN SOLUTION*. J Exp Med. 1977;146:257–270.

13. Medof ME, Iida K, Mold C, Nussenzweig V. UNIQUE ROLE OF THE COMPLEMENT RECEPTOR CR1 IN THE DEGRADATION OF C3b ASSOCIATED WITH IMMUNE COMPLEXES*. J Exp Med. 1982;156:1739–1754.

14. Whaley K, Ruddy S. Modulation of C3b Hemolytic Activity by a Plasma Protein Distinct from C3b Inactivator. Science (1979). 1976;193(4257):1011–1013. doi:10.1126/science.948757

15. Shiraishi S, Stroud RM. Cleavage products of C4b produced by enzymes in human serum. Immunochemistry. 1975;12(12):935–939. 10.1016/0019-2791(75)90256-6

16. Liszewski MK, Post TW, Atkinson JP. Membrane Cofactor Protein (MCP or CD46): Newest Member of the Regulators of Complement Activation Gene Cluster. Annu Rev Immunol. 1991;9(1):431–455. doi:10.1146/annurev.iy.09.040191.002243

17. Nagasawa S, Stroud RM. Mechanism of action of the C3b inactivator: Requiremen for a high molecular weight cofactor (C3b-C4bINA cofactor) and production of a new C3b derivative (C3b′). Immunochemistry. 1977;14(11):749–756. 10.1016/0019-2791(77)90345-7

18. Lachmann PJ, Halbwachs L. THE INFLUENCE OF C3b INACTIVATOR (KAF) CONCENTRATION ON THE ABILITY OF SERUM TO SUPPORT COMPLEMENT ACTIVATION. Vol 21.; 1975.

19. Kavanagh D, Yu Y, Schramm EC, et al. Rare genetic variants in the CFI gene are associated with advanced age-related macular degeneration and commonly result in reduced serum factor I levels. Hum Mol Genet. 2015;24(13):3861–3870. doi:10.1093/hmg/ddv091

20. Hallam TM, Marchbank KJ, Harris CL, et al. Rare genetic variants in complement factor i lead to low FI plasma levels resulting in increased risk of age-related macular degeneration. Invest Ophthalmol Vis Sci. 2020;61(6). doi:10.1167/IOVS.61.6.18

21. Jones A V., MacGregor S, Han X, et al. Evaluating a Causal Relationship between Complement Factor I Protein Level and Advanced Age-Related Macular Degeneration Using Mendelian Randomization. Ophthalmology Science. 2022;2(2). doi:10.1016/j.xops.2022.100146

22. Jones A V, Curtiss D, Harris C, et al. An assessment of prevalence of Type 1 CFI rare variants in European AMD, and why lack of broader genetic data hinders development of new treatments and healthcare access. PLoS One. 2022;17(9):e0272260-. 10.1371/journal.pone.0272260

23. Seddon JM, Rosner B, De D, Huan T, Java A, Atkinson J. Rare Dysfunctional Complement Factor I Genetic Variants and Progression to Advanced Age-Related Macular Degeneration. Ophthalmology Science. 2023;3(2):100265. 10.1016/j.xops.2022.100265

24. Java A, Baciu P, Widjajahakim R, et al. Functional Analysis of Rare Genetic Variants in Complement Factor I (CFI) using a Serum-Based Assay in Advanced Age-related Macular Degeneration. Transl Vis Sci Technol. 2020;9(9):37. doi:10.1167/tvst.9.9.37

25. Hallam TM, Cox TE, Smith-Jackson K, et al. A novel method for real-time analysis of the complement C3b:FH:FI complex reveals dominant negative CFI variants in age-related macular degeneration. Front Immunol. 2022;13. doi:10.3389/fimmu.2022.1028760

26. Hallam TM, Andreadi A, Sharp SJ, et al. Comprehensive functional characterization of complement factor I rare variant genotypes identified in the SCOPE geographic atrophy cohort. Journal of Biological Chemistry. 2024;300(7). doi:10.1016/j.jbc.2024.107452

27. Lachmann PJ, Lay E, Seilly DJ, Buchberger A, Schwaeble W, Khadake J. Further studies of the down-regulation by Factor I of the C3b feedback cycle using endotoxin as a soluble activator and red cells as a source of CR1 on sera of different complotype. Clin Exp Immunol. 2016;183(1):150—156. doi:10.1111/cei.12714

28. Pruimboom-Brees IM, Gupta S, Chemuturi N, et al. International consortium for innovation and quality: An industry perspective on the nonclinical and early clinical development of intravitreal drugs. Clin Transl Sci. 2023;16(5):723–741. doi:10.1111/cts.13480

29. Edmonds R, Steffen V, Honigberg LA, Chang MC. Alternative Complement Pathway Inhibition by Lampalizumab: Analysis of Data From Chroma and Spectri Phase III Clinical Trials. Ophthalmology Science. 2023;3(3). doi:10.1016/j.xops.2023.100286

30. Nürnberger W, Bhakdi S. Plasma C3d/C3 quotient as a parameter for in vivo complement activation. J Immunol Methods. 1984;74(1):87–91. 10.1016/0022-1759(84)90370-3

31. Kim AHJ, Strand V, Sen DP, et al. Association of Blood Concentrations of Complement Split Product iC3b and Serum C3 With Systemic Lupus Erythematosus Disease Activity. Arthritis and Rheumatology. 2019;71(3):420–430. doi:10.1002/art.40747

32. Lynch NJ, Willis CL, Nolan CC, et al. Microglial activation and increased synthesis of complement component C1q precedes blood-brain barrier dysfunction in rats. Mol Immunol. 2004;40(10):709–716. doi:10.1016/j.molimm.2003.08.009

33. Machacek M, Deschatelets P, Kim R, Johnson P, Grossi F. Prediction of duration of C3 inhibition with APL-2 in human eyes using a PK/PD binding model. Invest Ophthalmol Vis Sci. 2017;58:1971. https://iovs.arvojournals.org/article.aspx?articleid=2637839&resultClick=1

34. Reid MJ. Ocular Distribution of Pegcetacoplan in Rabbits Following a Single Intravitreal Injection. Invest Ophthalmol Vis Sci. 2021;62. https://iovs.arvojournals.org/article.aspx?articleid=2775957&resultClick=1

35. Mandava N, Tirado-Gonzalez V, Geiger MD, et al. Complement activation in the vitreous of patients with proliferative diabetic retinopathy. Invest Ophthalmol Vis Sci. 2020;61(11). doi:10.1167/IOVS.61.11.39

36. Wilson S, Siebourg-Polster J, Titz B, et al. Correlation of Aqueous, Vitreous, and Serum Protein Levels in Patients With Retinal Diseases. Transl Vis Sci Technol. 2023;12(11). doi:10.1167/tvst.12.11.9

37. Vogt SD, Curcio CA, Wang L, et al. Retinal pigment epithelial expression of complement regulator CD46 is altered early in the course of geographic atrophy. Exp Eye Res. 2011;93(4):413–423. 10.1016/j.exer.2011.06.002

38. Ricklin D, Reis ES, Mastellos DC, Gros P, Lambris JD. Complement component C3 – The “Swiss Army Knife” of innate immunity and host defense. Immunol Rev. 2016;274(1):33–58. doi:10.1111/imr.12500

39. Nielsen J, MacLaren RE, Heier JS, et al. Preliminary Results from a First-in-human Phase I/II Gene Therapy Study (FOCUS) of Subretinally Delivered GT005, an Investigational AAV2 Vector, in Patients with Geographic Atrophy Secondary to Age-related Macular Degeneration. Invest Ophthalmol Vis Sci. 2022;63(7):1504.

40. Tzoumas N, Kavanagh D, Cordell HJ, Lotery AJ, Patel PJ, Steel DH. Rare complement factor i variants associated with reduced macular thickness and age-related macular degeneration in the UK Biobank. Hum Mol Genet. 2022;31(16):2678–2692. doi:10.1093/hmg/ddac060

## Supplementary References

1. Xue K, Groppe M, Salvetti AP, MacLaren RE. Technique of retinal gene therapy: delivery of viral vector into the subretinal space. Eye. 2017;31(9):1308–1316. doi:10.1038/eye.2017.158

2. MacLaren RE, Groppe M, Barnard AR, et al. Retinal gene therapy in patients with choroideremia: initial findings from a phase 1/2 clinical trial. The Lancet. 2014;383(9923):1129–1137. 10.1016/S0140-6736(13)62117-0

3. Chang S. LXII Edward Jackson Lecture: Open Angle Glaucoma After Vitrectomy. Am J Ophthalmol. 2006;141(6). doi:10.1016/j.ajo.2006.02.014

4. Santos M. Sensitive AAV Capsid Protein Impurity Analysis by CE Using Easy to Label Fluorescent Chromeo Dye P503.; 2019.

